# Altered function of peripheral organ systems in first-episode drug naïve mental illnesses: A systematic review and meta-analysis

**DOI:** 10.64898/2026.08.01.26359479

**Authors:** Jetro E. Ang, Yangqi Xu, Vanessa Cropley, Andrew Zalesky, Ye Ella Tian

**Affiliations:** Department of Psychiatry, Melbourne Medical School, The University of Melbourne, Melbourne, Australia; School of Psychological Science, The University of Western Australia, Perth, Australia; St Vincent’s Hospital Melbourne, Melbourne, Australia; Orygen, Parkville, Australia; Centre for Youth Mental Health, The University of Melbourne, Melbourne, Australia; Department of Biomedical Engineering, Faculty of Engineering and Information Technology, The University of Melbourne, Melbourne, Australia

**Author notes:** These authors contributed equally (Jetro E. Ang, Yangqi Xu).

## Abstract

**Background:** Although mental illness is primarily regarded as a disorder of the brain, body system dysfunction is increasingly recognized as a salient biomarker in psychiatry, often emerging before the onset of overt symptoms. Here, we systematically review studies on peripheral organ systems (cardiovascular, metabolic, immune, liver, kidneys, lungs and muskeloskeletal) in schizophrenia, major depressive disorder (MDD), bipolar disorder (BD) and generalized anxiety disorder (GAD), aiming to synthesize findings on the function of multiple body systems in the early stages of mental illness.

**Methods:** EMBASE, MEDLINE and PsycINFO were searched from inception until 19 November 2024, identifying case-control studies comparing physiological markers of peripheral organ systems (i.e., cardiovascular, metabolic, immune, liver, kidney, lung and muskeloskeletal) in adults with one of the four mental illnesses at first-episode and drug naive, with healthy controls. We followed the PRISMA 2020 guidelines (PROSPERO: CRD42023408594).

**Results:** Of 2,637 citations retrieved, 138 studies met inclusion criteria for review with 52 markers of immune (n=32), metabolic (n=11), cardiovascular (n=6), liver (n=2) and musculoskeletal (n=1) function identified. 105 studies were eligible for meta-analysis, including 80, 26, 4 and 0 studies on schizophrenia, MDD, BD and GAD respectively. Meta-analysis revealed increased HDL-cholesterol, waist-hip-circumference ratio, triglycerides, insulin, insulin resistance, 2-hr glucose, neutrophil, monocyte, white blood cell, IL-4 and systolic blood pressure, and reduced albumin in schizophrenia; increased TNF-α and IL-10 in MDD; and increased IL-6 and IFN-γ in both schizophrenia and MDD. Other results were narratively discussed.

**Conclusions:** Alterations in peripheral organ function across multiple systems characterizes the onset of psychiatric disorders. However, research on peripheral organ function in psychiatry is limited and primarily focusses on the immune and metabolic systems.

## Introduction

Mental illness is associated with disproportionally prevalent chronic physical illnesses including coronary heart disease, diabetes and stroke, compared to the general population ^1–3^. Poor physical health in psychiatry contributes to inequalities in health care access ^4^, underdiagnosis ^5^ and increased mortality rates ^1,6^. Despite growing awareness of its impact, there remains continued difficulty in identifying and managing comorbid physical and mental health conditions ^1^.

Poor physical health has been considered a consequence of mental illness for decades, associated with adverse side effects of psychotropic medications, long-term illness, lifestyle changes and socioeconomic factors ^7–10^. Yet, growing evidence suggests that alterations in peripheral organ function are already present at the onset of mental illness, prior to long-term medication treatment. This suggests that mental illness is a multisystem disorder that confers an inherent risk of peripheral organ dysfunction ^11^, prior to diagnosis and treatment, leading to increased risk of physical illnesses.

Meta-research to date has largely focused on physiological dysregulation in metabolic, immune and the hypothalamic-pituitary-adrenal (HPA) systems, possibly due to their direct relevance to increased disease risk of cardiometabolic conditions. However, poor physical health is also evident in other organ systems, e.g., liver and kidneys, in people with mental illnesses ^12^. Understanding how early physiological changes across body systems coincide with emerging mental illness is essential for advancing integrated models of disease risk that account for both mental and physical illnesses.

So far, evidence is most abundant for disturbances in glucose ^13–15^ and lipid ^13,16^ metabolism, immune responses ^15,17–19^ and stress hormones related to the HPA axis ^20,21^ in first-episode schizophrenia and related psychosis. In contrast, evidence is scanter for other common mental illnesses including major depressive disorder ^15,22,23^ and bipolar disorder ^18^, with little research occurring in generalized anxiety disorder, an anxiety disorder that also frequently co-occurs with major depressive disorder ^24,25^. Despite high comorbidity rates ^26^ and possibly shared neuropathology across mental illnesses ^27^, the extent to which peripheral organ dysfunction is a transdiagnositc characteristic across these mental illnesses remains unknown.

Here, we report on the largest and most comphrensive systematic review and meta-analysis to date investigating alterations in physiological function across seven peripheral organ systems (cardiovascular, metabolic, immune, liver, kidneys, lungs and muskeloskeletal) in common psychiatric disorders. Our analyses provide new insights into peripheral organ function across schizophrenia, major depressive disorder, bipolar disorder and generalized anxiety disorder. We focus on drug naïve patients who were first-episode or at early stage of illness. We sought to identify common and unique patterns of physical health manifestations across the four mental illnesses.

## Methods

### Search Strategy

We followed the PRISMA guidlines ^28^ for systematic review and meta-analysis. The PRISMA Checklist is described in Supplementary Table 1. We searched EMBASE, MEDLINE, and PsycINFO from inception until 19 November 2024. Our search terms included a comprehensive set of physiological phenotypes indicative of peripheral organ function that i) are commonly used in clinical settings to index organ health and function for a given organ system (e.g., systolic blood pressure for cardiovascular; cholesterol for metabolic; albumin for liver; creatinine for kidney); or ii) were reported in existing literature assessing physical health in people with mental illness. We focused on physiological markers indicative of organ function that are already widely assayed in primary care or readily accessible at minimal cost. We did not consider complex body imaging measures, particularly those that require an injection of tracer (e.g., positron emission tomography) or contrast agent (e.g., contrast-enhanced magnetic resonance imaging). See Supplementary Table 2 for search terms used in the database.

### Selection criteria

Searches were limited to peer-reviewed, human studies published in the English language. Studies were eligible for inclusion if they assessed at least one physiological marker in both adult (at least 18 years old) patients and healthy control groups. Patients included i) had a diagnosis of either schizophrenia (or schizoaffective disorder, schizophreniform disorder, schizophrenia spectrum or psychotic disorder not otherwise specified), major depressive disorder, bipolar disorder (or mania), or generalized anxiety disorder according to the Diagnostic and Statistical Manual, 4th edition or higher, or the International Classification of Diseases, 10th edition or higher; ii) were in the first episode of illness or have an illness duration less than five years, following the criteria used in previous meta-analyses ^13,29^ and the operational definition for first-episode psychosis proposed by Breitborde and colleagues ^30^; and iii) were psychotropic medication naïve or had minimal exposure, defined as less than two weeks ^13^.

Studies were excluded if patients i) had multiple episodes of mental illness; ii) were diagnosed with a substance or medication induced mental disorder; iii) had comorbid mental or physical health conditions that may affect the regulatory function of the relevant body systems studied; or iv) received chronic psychotropic medication exposure (greater than two weeks). Studies that did not provide phenotype values in a healthy control group were also excluded. Conference abstracts, dissertations, opinion papers, and review studies were excluded.

### Data extraction

Two investigators (JA, YX) screened the titles and abstracts of all articles against the eligibility criteria. Articles that appeared to meet the inclusion criteria were retrieved for full-text screening by both investigators (JA, YX) to determine the eligibility of articles for final inclusion. Screening conflicts between the reviewers were resolved through discussion or with a third investigator (YET).

The data collection process was performed by two investigators (JA, YX) independently and then cross-checked by each other for consistency. An electronic data extraction sheet was developed and refined by the investigators during the data extraction process. Key variables collected included authors, publication year, country, sample size, participant ethnicity, mean age, mental health diagnosis, age of illness onset, illness duration, comorbid physical or mental health conditions, medication use, and reported phenotype values (mean and standard deviation, SD) for both patient and healthy control groups.

### Risk of bias assessment

Included studies were evaluated for methodological quality and risk of bias using the Joanna Briggs Institute critical appraisal checklist for case control studies ^31^.

### Data analysis

Meta-analyses were conducted to compare reported values of organ health phenotypes between patients and healthy controls. Analyses were separately performed for each organ function marker and grouped by mental illness diagnosis. Meta-analyses were conducted using the R Metafor package ^32^ in jamovi with a random effect model, when at least three eligible studies reported the sample size, mean, and standard deviation of outcomes for patients and healthy controls. In the case of shared cohorts across multiple studies, the study that had the largest sample size, reported the most variety of phenotypes, or was the most recently published, was used in that order. Studies were considered to originate from the same cohort only when this was explicitly stated in the respective publications. Studies recruiting participants from the same hospital or institution, but without evidence of a shared cohort, were treated as independent. Selection of studies with shared cohorts were individually decided for meta-analysis of each marker.

Overlap in study inclusion between pairs of organ systems was assessed using the Jaccard coefficient, defined as 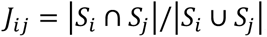, where *S_i_* is a list of cohorts unique to the organ/system with index *i*. We also considered a sample-size weighted variant of the Jaccard coefficient, defined as

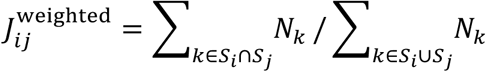

In this weighted formulation, *N_k_* denotes the sample size of patients in cohort *S_i_*, thus accounting for differences in sample sizes when computing cohort reuse. A narrative synthesis was performed for studies not eligible for meta-analysis.

The ‘restricted maximum-likelihood’ model was used to estimate heterogeneity between studies ^33,34^ and we report Q-test and *I*^2^ statistics for heterogeneity among studies ^35^. Standardized mean differences (SMD) were calculated for eligible studies with 95% confidence intervals. A two-sided p<0.05 was deemed significant. Multiple comparison correction was performed using the Benjamini-Hochberg method ^36^. The false discover rate (FDR) was controlled at 0.05 across the set of markers in each disorder group respectively. SMDs were utilized due to the expectation of high study heterogeneity (i.e., differences in the sensitivity of cytokine measurement methodology and reported units), which allowed for standardized analysis of results examining differences between groups within studies ^37^. Effect sizes were interpreted using Cohen’s d guidelines, such that SMD =0.2 small, SMD=0.5 medium, and SMD = 0.8 large ^38,39^.

Studentized residuals and Cook’s distances were used to assess outliers or overly influential studies. Specifically, studies with a studentized residual larger than the 100 x (1 - 0.05/(2 × *k*))th percentile of a standard normal distribution are considered potential outliers (i.e., using a Bonferroni correction with two-sided alpha = 0.05 for *k* studies included in the meta-analysis). Studies with a Cook’s distance larger than the median plus six times the interquartile range of the Cook’s distances are considered influential. Sensitivity analysis was performed by re-running meta-analysis excluding outlier and/or influential studies.

This study was registered with PROSPERO (CRD42023408594).

## Results

A total of 2,637 citations were retained for title and abstract screening after deduplication. Initial screening excluded 2,068 studies with 569 sought for full-text screening, and 545 ultimately retrieved. At the full-text screening stage, 407 studies were excluded, and 138 studies (including 25 reports) were included for review (Supplementary Table 3). Reports were identified as papers including samples duplicated in another study already included for review. Data from reports were combined where possible, alternatively the data was selected based on i) outcomes examined which were not included in the other reports and ii) largest sample size in the case where phenotypes were examined in multiple reports. Figure 1 shows the process of study selection. Among the 138 included studies, 43 studies examined a shared patient population with one or more other studies. A given cohort is never shared across more than 5 studies (Supplementary Table 4). The remaining 95 studies represent independent cohorts to the best of our knowledge. A list of studies excluded at the full-text screening stage and their reasons for exclusion is provided in Supplementary Table 5.

**Figure 1.**
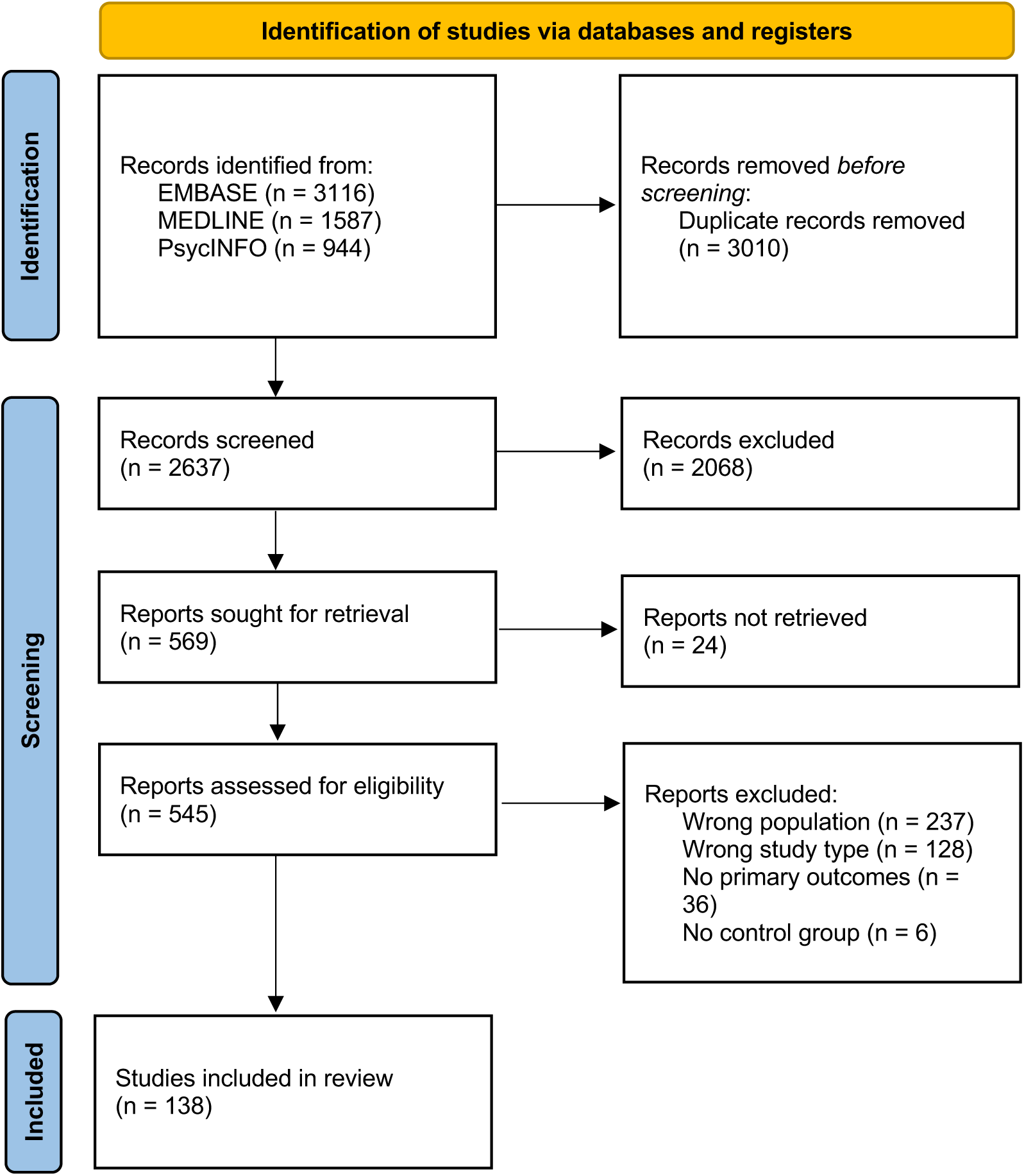
Overview of study selection. Schematic of literature review, inclusion and exclusion of studies. All 138 included studies were deemed to have low risk of bias (Supplementary Table 6). Studies were not excluded or weighted for analysis based on methodological quality or explicit inclusion and exclusion criteria for healthy control individuals, so long as a healthy control group was defined. Most studies defined healthy control individuals as no psychiatric (90.6%) nor major physical (76.8%) illness, while a small proportion of studies (9.4%) did not report their criteria for healthy controls. Amongst the included studies, 105 were eligible for meta-analysis. Of these, 94 studies included first episode patients, and 11 studies did not specify the number of episodes. Papers were excluded when there were insufficient studies to examine a particular phenotype (minimum of 3 studies required) or when data was not possible to synthesize (e.g., no mean or standard deviation provided).

For the meta-analysis, we identified 80 eligible studies with a total of 6,013 patients with schizophrenia (mean age (SD, years): 28.70 (7.63); 3,278 males) and 5,352 HC (mean age (SD): 28.90 (6.77); 2879 males); 26 studies with 1,694 patients with major depressive disorder (mean age (SD): 32.08 (8.94); 706 males) and 1,278 HC (mean age (SD): 32.63 (8.17) ; 552 males); and 4 studies on bipolar disorder with 475 patients (mean age (SD): 25.13 (7.93); 186 males) and 382 HC (mean age (SD): 24.65 (7.23); 144 males). We did not identify sufficient studies to perform meta-analysis for generalized anxiety disorder. For the narrative synthesis, 44 studies were discussed, among which 20 studies examined schizophrenia, 17 on major depressive disorder, 9 on bipolar disorder, and 2 on generalized anxiety disorder, noting that some studies discussed multiple disorders.

In terms of the measures of peripheral organ systems, a total of 52 physiological markers were identified, including measures indicating peripheral inflammation (n=32), metabolic (n=11), cardiovascular (n=6), liver (n=2) and musculoskeletal function (n=1). Studies of kidney and lung function were not identified. Meta-analysis was performed for 36 markers in schizophrenia, 8 in major depressive disorder, and 1 in bipolar disorder. The overlap in study inclusion between organ systems is modest for all system pairs (Jaccard average: 4%; weighted Jaccard average: 4%), except for the metabolic-immune system pair in the case of MDD (Jaccard: 39%; weighted Jaccard: 34%, Supplementary Figure 1). Consequently, differences between these two systems in MDD cannot be considered fully independent, and findings for this pair should be interpreted with caution. Narrative synthesis was performed for 15, 33, 18 and 9 markers in schizophrenia, major depressive disorder, bipolar disorder and generalized anxiety disorder respectively (Figure 2).

**Figure 2.**
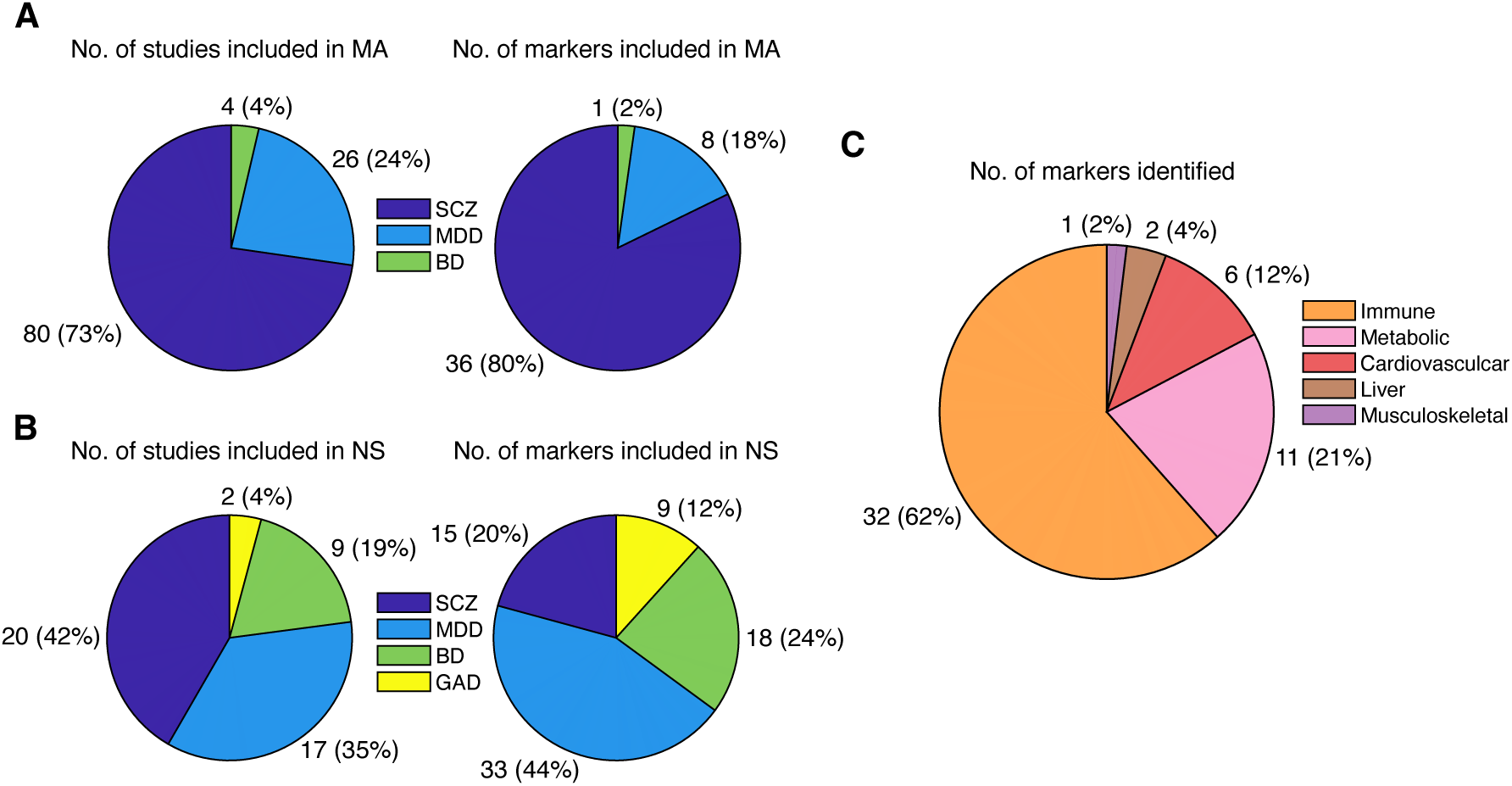
Distribution of eligible studies and markers in the literature survey across disorder groups and across organ systems. **A**) Proportion of studies and physiological markers eligible for meta-analysis (MA). **B**) Proportion of studies and physiological markers eligible for narrative sythesis (NS). **C**) Proportion of physiological markers eligible for MA across organ systems. SCZ, schizophrenia; MDD, major depressive disorder; BD, bipolar disorder; GAD, generalized anxiety disorder; No., number.

In the following sections, we present our findings for each organ system and compare the results across schizophrenia, major depressive disorder, bipolar disorder and generalized anxiety disorder, where possible. Results of the meta-analysis are provided in Table 1 and Figure 3. Raw measurements used for the meta-analysis extracted from eligible studies are provided in Supplementary Tables 7-20. Results of the meta-analysis excluding studies that included patients with ambiguous number of episodes are provided in Supplemenatry Table 21.

**Figure 3.**
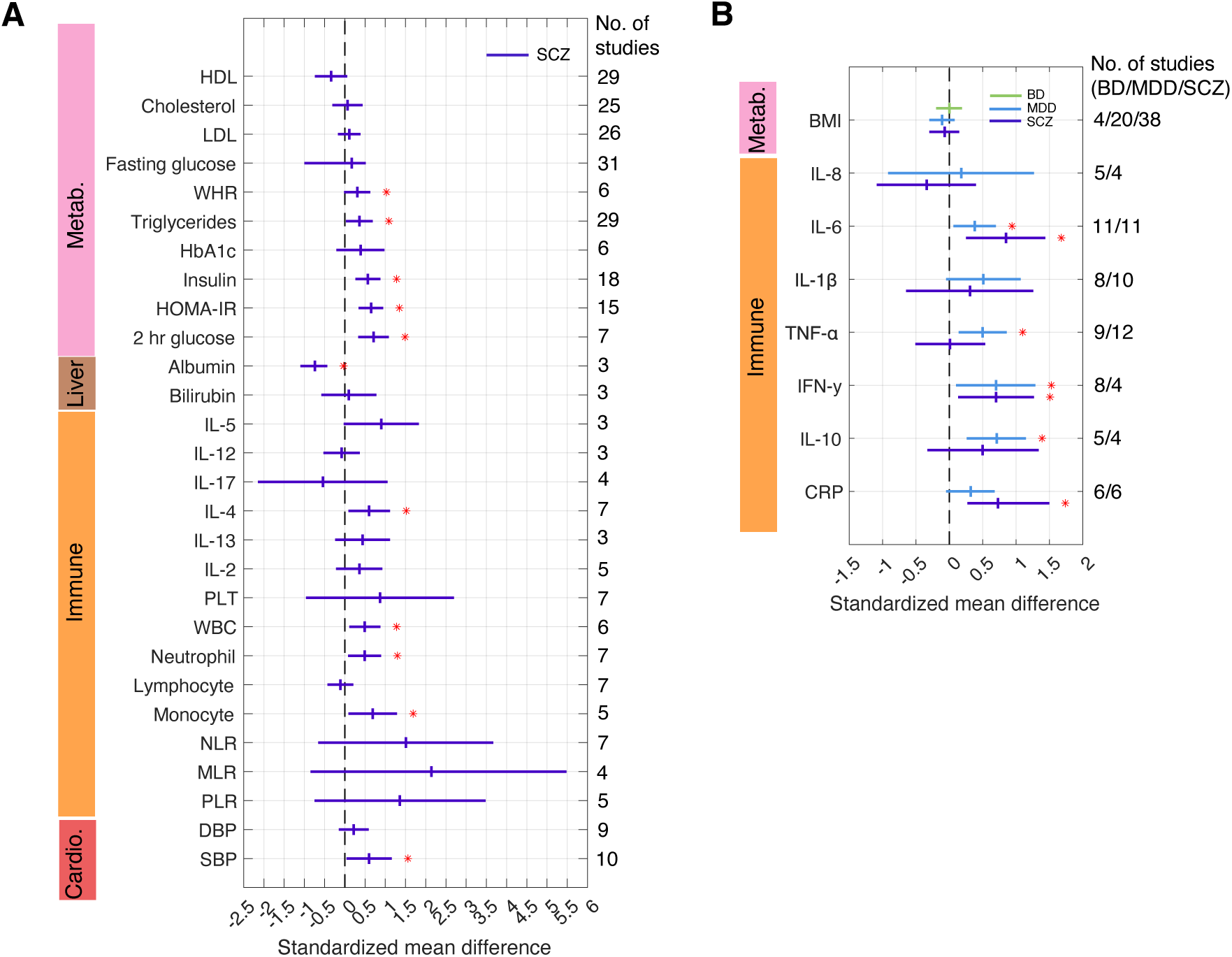
Forest plots for standarded mean differences of peripheral physiological markers in patients compared to healthy controls. **A**) Summary stardardized mean difference (SMD) between schizophrenia and healthy controls (HC) for physioloigcal markers eligible for meta-analysis. **B**) Summary SMD between schizophrenia (SCZ), major depressive disorder (MDD), bipolar disorder (BD) and HC respectively. A positive/negative SMD indicates higher/lower mean differences in patients compared to HC. The vertical marker in the middle of each horizontal line indicates the summary SMD and the width of the horizontal line indicates 95% confidence intervals. Markers that show significant between-group differences (p<0.05, FDR corrected, two-sided) are indicated with an asterisk. Cardio., cardiovascular; HDL, high-density lipoprotein cholesterol; LDL, low-density lipoprotein cholesterol; WHR, waist-hip circumferance ratio; HbA1c, glycated hemoglobin; HOMR-IR, insulin resistance; IL, interlukin; WBC, white blood cell count; PLT, platelet; PLR, platelet-lymphocyte ratio; NLR, neutrophil-lymphocyte ratio; MLR, monocyte-lymphocyte ratio; DBP, diastolic blood pressure; SBP, systolic blood pressure; BMI, body mass index; CRP, c-reactive protein; TNF, tumor necrosis factor; IFN, interferon.

**Table 1.** Summary of meta-analysis results.

| Marker | Category | No. of Studies | Patients (n) | Patients Age (Mean (SD)) | Patients Sex (Male%) | Controls (n) | Controls Age (Mean (SD)) | Controls Sex (Male %) | Statistic (SMD) | P-value | Effect Size | Heterogeneity (I<sup>2</sup>) |
| --- | --- | --- | --- | --- | --- | --- | --- | --- | --- | --- | --- | --- |
| **SCZ** |  |  |  |  |  |  |  |  |  |  |  |  |
| **IFN-γ\*** | Immune | 8 | 392 | 28.74 (6.81) | 54.0% | 368 | 27.34 (5.33) | 56.5% | 0.70 (95% CI: 0.19 to 1.21) | 0.007 | Medium | High (90.79%) |
| TNF-α | Immune | 9 | 485 | 27.17 (6.23) | 60.8% | 528 | 27.47 (5.68) | 54.7% | 0.01 (95% CI: -0.45 to 0.48) | 0.952 | Very small | High (91.95%) |
| IL-1β | Immune | 8 | 347 | 25.09 (5.78) | 60.2% | 356 | 26.65 (4.77) | 52.8% | 0.31 (95% CI: -0.59 to 1.20) | 0.504 | Small | High (96.70%) |
| IL-5 | Immune | 3 | 88 | 25.76 (5.57) | 64.7% | 87 | 26.56 (3.41) | 59.8% | 0.90 (95% CI: 0.07 to 1.73) | 0.033 | Large | High (84.86%) |
| **IL-6\*** | Immune | 11 | 565 | 28.12 (7.11) | 53.3% | 555 | 27.28 (5.67) | 51.4% | 0.85 (95% CI: 0.31 to 1.38) | 0.002 | Large | High (94.3%) |
| IL-8 | Immune | 5 | 160 | 25.66 (5.42) | 61.9% | 170 | 26.70 (4.93) | 51.8% | -0.34 (95% CI: -1.03 to 0.34) | 0.325 | Small | High (88.81%) |
| IL-12 | Immune | 4 | 128 | 27.01 (6.83) | 64.8% | 127 | 27.75 (5.04) | 59.8% | -0.08 (95% CI: -0.43 to 0.27) | 0.649 | Very small | Low (47.91%) |
| IL-17 | Immune | 4 | 345 | 30.31 (8.27) | 65.8% | 256 | 28.41 (5.84) | 69.5% | -0.54 (95% CI: -2.05 to 0.96) | 0.479 | Medium | High (98.6%) |
| **IL-4\*** | Immune | 7 | 323 | 29.00 (6.61) | 54.1% | 308 | 27.44 (5.51) | 58.8% | 0.60 (95% CI: 0.19 to 1.02) | 0.005 | Medium | High (83.19%) |
| IL-10 | Immune | 5 | 217 | 27.20 (5.63) | 59.4% | 247 | 26.25 (4.59) | 53.8% | 0.50 (95% CI: -0.27 to 1.28) | 0.205 | Medium | High (93.58%) |
| IL-13 | Immune | 3 | 81 | 26.36 (6.05) | 50.8% | 89 | 26.67 (4.69) | 46.1% | 0.44 (95% CI: -0.14 to 1.02) | 0.134 | Small | Moderate (69.96%) |
| IL-2 | Immune | 5 | 217 | 27.20 (5.63) | 59.4% | 247 | 26.25 (4.59) | 53.8% | 0.36 (95% CI: -0.12 to 0.83) | 0.139 | Small | High (82.82%) |
| **CRP\*** | Immune | 6 | 296 | 29.44 (8.27) | 63.2% | 333 | 30.53 (8.87) | 61.0% | 0.73 (95% CI: 0.33 to 1.14) | < 0.001 | Medium | High (82.42%) |
| PLT | Immune | 7 | 563 | 27.43 (7.37) | 63.8% | 511 | 29.14 (6.65) | 51.3% | 0.87 (95% CI: -0.86 to 2.60) | 0.325 | Large | High (99.42%) |
| **WBC\*** | Immune | 6 | 320 | 27.81 (8.04) | 60.9% | 388 | 30.15 (7.77) | 57.2% | 0.49 (95% CI: 0.21 to 0.78) | < 0.001 | Small | Moderate (69.08%) |
| **Neutrophil\*** | Immune | 7 | 495 | 28.26 (8.87) | 66.1% | 440 | 30.58 (8.17) | 53.6% | 0.49 (95% CI: 0.18 to 0.80) | 0.002 | Small | High (79.68%) |
| Lymphocyte | Immune | 7 | 495 | 28.26 (8.87) | 66.1% | 440 | 30.58 (8.17) | 53.6% | -0.11 (95% CI: -0.33 to 0.11) | 0.322 | Very small | Moderate (60.82%) |
| **Monocyte\*** | Immune | 5 | 283 | 27.56<br>(7.50) | 61.8% | 345 | 30.25<br>(7.77) | 57.7% | 0.69 (95% CI:<br>0.19 to 1.19) | 0.007 | Medium | High (88.78%) |
| NLR | Immune | 7 | 538 | 27.45<br>(7.66) | 63.2% | 482 | 28.81<br>(6.51) | 48.8% | 1.51 (95% CI: -<br>0.56 to 3.57) | 0.153 | Large | High (99.47%) |
| MLR | Immune | 4 | 182 | 26.12<br>(5.37) | 56.9% | 218 | 28.35<br>(5.84) | 50.3% | 2.41 (95% CI: -<br>0.75 to 5.58) | 0.135 | Large | High (99.55%) |
| PLR | Immune | 5 | 325 | 26.53<br>(6.19) | 56.6% | 381 | 28.46<br>(6.07) | 50.7% | 1.36 (95% CI: -<br>0.65 to 3.38) | 0.185 | Large | High (99.19%) |
| BMI | Metabolic<br>function | 38 | 3702 | 28.83<br>(8.36) | 53.2% | 3458 | 29.12<br>(7.62) | 53.7% | -0.07 (95% CI:<br>-0.24 to 0.09) | 0.386 | Very<br>small | High (91.00%) |
| **WHR\*** | Metabolic<br>function | 6 | 535 | 27.28<br>(8.50) | 65.0% | 329 | 26.29<br>(4.60) | 63.8% | 0.31 (95% CI:<br>0.08 to 0.53) | 0.007 | Small | Moderate<br>(52.88%) |
| Fasting<br>glucose | Metabolic<br>function | 31 | 2906 | 27.40<br>(8.88) | 53.5% | 2233 | 31.19<br>(8.89) | 53.2% | 0.17 (95% CI: -<br>0.90 to 0.42) | 0.201 | Very<br>small | High (94.46%) |
| **2 hr glucose\*** | Metabolic<br>function | 7 | 726 | 24.28<br>(7.25) | 58.0% | 345 | 27.83<br>(5.64) | 58.8% | 0.71 (95% CI:<br>0.43 to 0.99) | < 0.001 | Medium | High (75.73%) |
| **Insulin\*** | Metabolic<br>function | 18 | 1696 | 28.71<br>(8.75) | 50.6% | 1312 | 28.99<br>(7.01) | 48.6% | 0.57 (95% CI:<br>0.36 to 0.78) | < 0.001 | Medium | High (84.15%) |
| **HOMA-IR\*** | Metabolic<br>function | 15 | 1585 | 27.56<br>(8.54) | 50.4% | 1130 | 26.52<br>(4.55) | 49.2% | 0.65 (95% CI:<br>0.44 to 0.85) | < 0.001 | Medium | High (80.36%) |
| HbA1c | Metabolic<br>function | 6 | 956 | 28.95<br>(9.17) | 54.6% | 808 | 29.53<br>(7.76) | 52.7% | 0.39 (95% CI: -<br>0.11 to 0.88) | 0.126 | Small | High (94.93%) |
| Cholesterol | Metabolic<br>function | 25 | 2610 | 29.38<br>(9.60) | 53.3% | 2038 | 30.04<br>(8.28) | 50.8% | 0.07 (95% CI: -<br>0.21 to 0.34) | 0.622 | Very<br>small | High (94.62%) |
| **Triglycerides\*** | Metabolic<br>function | 29 | 3004 | 31.31<br>(9.98) | 53.7% | 2260 | 32.10<br>(10.04) | 52.6% | 0.36 (95% CI:<br>0.13 to 0.59) | 0.002 | Small | High (93.35%) |
| HDL | Metabolic<br>function | 29 | 3120 | 29.26<br>(8.64) | 53.7% | 2416 | 29.47<br>(6.83) | 53.5% | -0.34 (95% CI:<br>-0.64 to -0.04) | 0.025 | Small | High (96.24%) |
| LDL | Metabolic<br>function | 26 | 2737 | 29.17<br>(8.57) | 52.5% | 2159 | 29.08<br>(6.86) | 49.1% | 0.11 (95% CI: -<br>0.07 to 0.29) | 0.223 | Very<br>small | High (88.24%) |
| **Albumin\*** | Liver function | 3 | 153 | 29.85<br>(8.48) | 62.7% | 226 | 29.11<br>(6.73) | 56.6% | -0.74 (95% CI:<br>-1.00 to -0.53) | < 0.001 | Medium | Low (0%) |
| Bilirubin | Liver function | 3 | 232 | 28.97<br>(8.43) | 45.7% | 196 | 29.06<br>(6.63) | 49.0% | 0.10 (95% CI: -<br>0.48 to 0.68) | 0.744 | Very<br>small | High (86.92%) |
| **SBP\*** | Cardiovascular<br>function | 10 | 1515 | 30.33<br>(9.03) | 53.9% | 1254 | 30.63<br>(8.71) | 57.1% | 0.60 (95% CI:<br>0.14 to 1.06) | 0.01 | Medium | High (96.97%) |
| DBP | Cardiovascular<br>function | 9 | 1365 | 30.16<br>(9.03) | 52.2% | 1134 | 30.28<br>(8.70) | 53.2% | 0.22 (95% CI: -<br>0.05 to 0.49) | 0.109 | Very<br>small | High (89.43%) |

| MDD |  |  |  |  |  |  |  |  |  |  |  |  |
| --- | --- | --- | --- | --- | --- | --- | --- | --- | --- | --- | --- | --- |
| IFN-γ* | Immune | 4 | 225 | 36.62<br>(12.08) | 47.6% | 165 | 37.83<br>(8.93) | 58.8% | 0.70 (95% CI:<br>0.16 to 1.23) | 0.01 | Medium | High (83.57%) |
| TNF-α* | Immune | 12 | 718 | 32.98<br>(9.97) | 40.4% | 597 | 33.60<br>(8.17) | 42.5% | 0.50 (95% CI:<br>0.20 to 0.80) | < 0.001 | Medium | High (85.22%) |
| IL-1β | Immune | 10 | 497 | 34.68<br>(10.81) | 39.0% | 442 | 35.20<br>(8.45) | 41.9% | 0.51 (95% CI:<br>0.01 to 1.01) | 0.044 | Medium | High (92.19%) |
| IL-6* | Immune | 11 | 658 | 32.34<br>(9.99) | 39.5% | 516 | 33.51<br>(8.62) | 40.7% | 0.38 (95% CI:<br>0.12 to 0.64) | 0.004 | Small | High (78.06%) |
| IL-8 | Immune | 4 | 225 | 34.20<br>(12.00) | 41.7% | 185 | 35.50<br>(9.14) | 44.3% | 0.18 (95% CI: -<br>0.86 to 1.21) | 0.736 | Very<br>small | High (95.46%) |
| IL-10* | Immune | 4 | 407 | 31.19<br>(10.29) | 42.3% | 281 | 32.70<br>(8.27) | 44.5% | 0.71 (95% CI:<br>0.32 to 1.09) | < 0.001 | Medium | High (81.53%) |
| CRP | Immune | 6 | 259 | 29.96<br>(9.55) | 31.3% | 239 | 29.52<br>(8.90) | 34.3% | 0.32 (95% CI:<br>0.01 to 0.62) | 0.043 | Small | Moderate<br>(61.56%) |
| BMI | Metabolic<br>function | 20 | 1412 | 30.94<br>(8.27) | 41.5% | 1050 | 31.34<br>(7.51) | 43.4% | -0.11 (95% CI:<br>-0.24 to 0.02) | 0.092 | Very<br>small | Moderate<br>(56.14%) |
| BD |  |  |  |  |  |  |  |  |  |  |  |  |
| BMI | Metabolic<br>function | 4 | 475 | 29.18<br>(9.21) | 45.5% | 382 | 29.80<br>(8.73) | 45.6% | 0.00 (95% CI: -<br>0.14 to 0.13) | 0.951 | Very<br>small | Low (0%) |

\* $p < 0.05$ FDR corrected. **Abbreviations:** BD, Bipolar Disorder; BMI, body mass index; CRP, C-reactive protein; DBP, diastolic blood pressure; HbA1c, glycated haemoglobin; HDL, high-density lipoprotein; HOMA-IR, Homeostatic Model Assessment for Insulin Resistance; IFN, interferon; IL, interleukin; LDL, low-density lipoprotein; MDD, Major Depressive Disorder; MLR, monocyte-lymphocyte ratio; NLR, neutrophil-lymphocyte ratio; PLR, platelet-lymphocyte ratio; SCZ, Schizophrenia; SBP, systolic blood pressure; TNF, tumor necrosis factor; WBC, white blood cell; WHR, waist-hip circumference ratio.

### Immune system

#### Cytokines

We identified 12 pro-inflammatory cytokines including interferon (IFN)-γ, interleukin (IL)-1α, IL-1β, IL-5, IL-6, IL-8, IL-9, IL-12, IL-17, IL-23, tumor necrosis factor (TNF-α) and TNF-β; 3 anti-inflammatory cytokines including IL-4, IL-10 and IL-13; and 8 adaptive-inflammatory cytokines including IL-2, IL-3, IL-7, IL-15, IL-27, transforming growth factor (TGF)-β, TGF-β1 and TGF-β2. Meta-analyses were performed for IL-1β, IL-6, IL-8, IFN-γ,TNF-α, and IL-10 in schizophrenia and major depressive disorder, and for IL-5, IL-8, IL-12, IL-17, IL-4, IL-13 and IL-2 in schizophrenia. Overall, we found significantly elevated pro- and anti-inflammatory cytokines in both schizophrenia and major depressive disorder. We did not find significant alterations in adaptive inflammatory markers. Other findings were narratively summarized where possbile. Detailed results are presented below.

Among the 12 pro-inflammatory cytokins, we found significantly elevated IL-6 and IFN-γ in both schizophrenia and major depressive disorder compared to HC. While elevated IFN-γ were comparable across schizophrenia (SMD=0.7, p=0.007) and major depressive disorder (SMD=0.7, p=0.01), IL-6 elevation was larger in schizophrenia (SMD=0.85, p=0.002) comapred to major depressive disorder (SMD=0.38, p=0.004). TNF-α (SMD=0.50, p=0.001) was significantly elevated in major depressive disorder but not in schizophrenia (p>0.05). Meta-analysis also found no significant difference for IL-5, IL-8, IL-12 nor IL-17 (p>0.05) in schizophrenia. Study heterogeneity was high (*I*^2^>83%) for all cytokines except IL-12 (I^2^=47.9%). The significance of the results remained unchanged after excluding outlier/influential studies for IL-6 (n=1), IL-8 (n=1), IL-17 (n=1) and TNF-α (n=2) in schizophrenia.

In studies not eligilbe for meta-analysis, we observed higher IL-5 ^40^, IL-12 ^41^ and IL-17 ^42,40^ in major depressive disorder. Findings of IL-8 in major depressive disorder were mixed across studies, with one study reporting lower IL-8 in major depressive disorder compared to HC ^43^, and another reporting no significant differences ^40^. We also observed higher IL-6, IFN-γ, IL-8, IL-12 in generalized anxiety disorder compared to HC, but no difference for IL-15 as reported in one study ^44^.

The remaining pro-inflammatory cytokines IL-1α, IL-9, IL-23, and TNF-β have only been examined in several individual studies and were not eligible for meta-analysis. An elevated IL-1α was reported in one schizophrenia ^45^ and one generalized anxiety disorder ^44^ study respectively, but did not differ for major depressive disorder ^46^. An elevated IL-23 was reported in schizophrenia compared to HC in one study ^47^, but no significant difference was reported in major depressive disorder ^40^. Neither IL-9 ^48^ nor TNF-β ^49^ showed significantly different levels in schizophrenia compared to HC. We did not identify any studies reporting pro-inflammatory cytokines in bipolar disorder.

Among the 3 anti-inflammatory cytokines, meta-analysis was performed for IL-10 in schizophrenia and major depressive disorder, and for IL-4 and IL-13 in schizophrenia. We found significantly elevated IL-10 in major depressive disorder (SMD=0.71, p<0.001) but not in schizophrenia (p>0.05). IL-4 was significantly elevated in schizophrenia (SMD=0.6, p=0.005), but there was no significant between-group difference for IL-13 (p>0.05). Study heterogeneity was moderate to high (*I*^2^=69.9% - 93.6%). Sensitivity analyses were performed for IL-10 in schizophrenia and major depressive disorder after excluding one influential study respectively and the significance of the results remained the same. Further investigation in major depressive disorder in narrative synthesis did not identify altered IL-4 nor IL-13 ^40^. No reports were available for bipolar disorder or generalized anxiety disorder.

Among the 8 adaptive-inflammatory cytokines, meta-analysis was only available for IL-2 and only in schizophrenia, which did not show significant alteration (p>0.05). Study heterogeneity was high (*I^2^*=82.8%). Narrative synthesis of IL-2 in two major depressive disorder ^40,41^ and one generalized anxiety disorder study ^44^ found significantly higher IL-2 in patients. Other adaptive-inflammatory cytokines have only been reported in individual studies. A decreased IL-3 in schizophrenia was reported in two studies ^50,51^. Findings for IL-7 were mixed, with one study showing elevated concentration in schizophrenia ^52^ and another study reporting no differences between schizophrenia and HC ^48^; while no differences were found between major depressive disorder and HC ^40^. IL-15 and IL-27 did not differ between schizophrenia and HC, according to Frydecka et al (2018) ^48^ and Borovcanin et al (2012) ^47^, respectively. An elevated TGF-β ^53^ and TGF-β1 ^54^ in schizophrenia has also been reported; however, no between-group differences were found for TGF-β2 ^55^. We identified one major depressive disorder study that assessed TGF-β1 ^43^, with no significant between-group differences reported. No reports were available for bipolar disorder or generalized anxiety disorder.

#### Other inflammatory markers

In addition to cytokines, several other inflammatory markers including c-reactive protein (CRP) and immune cells including white blood cell (WBC), neutrophil, monocyte, lymphocyte and platelet were assessed. Meta-analysis was performed for CRP in schizophrenia and major depressive disorder. Other markers were only meta-analysed in schizophrenia. Study heterogeneity was moderate to high (*I^2^*=60.8% - 99.6%). In the meta-analysis, we found significantly elevated CRP in schizophrenia (SMD=0.73, p<0.001) compared to HC, while elevated CRP in major depressive disorder was only nominally significant (SMD=0.32, p=0.043). Meta-analysis also revealed significantly elevated WBC (SMD=0.49, p<0.001), neutrophil (SMD=0.49, p=0.002) and monocyte count (SMD=0.69, p=0.007) in schizophrenia compared to HC, but no significant difference for other blood cell count measures (p>0.05) including platelet, lymphocyte, platelet-lymphocyte ratio (PLR), neutrophil-lymphocyte ratio (NLR) and monocyte-lymphocyte ratio (MLR). Sensitivity analyses were performed for monocyte, platelet, PLR, NLR and MLR in schizophrenia after excluding one influential study respectively and the results remained the same.

While not feasible for meta-analysis, elevated CRP has been reported in generalized anxiety disorder ^44^, and has mixed evidence in bipolar disorder studies, with two reporting elevated levels in patients ^56, 57^ but one finding no difference ^58^. We also observed elevated WBC ^59^, neutrophil count ^59,60^, platelet ^59^ and PLR ^59^ in major depressive disorder, but no difference for monocyte count ^59,60^ nor MLR ^60^. Findings for lymphocyte count and NLR were mixed across studies in major depressive disorder, with one study reporting significant alteration (i.e., decreased lymphocyte and increased NLR) ^59^ and another study reporting no differences ^60^. For bipolar disorder, we observed elevated monocyte count ^58^ and NLR ^61^ in patients compared to HC, but no differences for WBC ^58^, platelet ^58,61,62^ nor PLR ^61^. Findings for neutrophil and lymphocyte count were mixed across studies in bipolar disorder, with one study reporting significant alteration (i.e., increased neutrophil and descreased lymphocype count) in patients ^61^ and another reporting no between-group differences ^58^.

### Metabolic system

Body mass index (BMI), waist-hip circumference ratio and blood-based markers relevant to glucose (fasting glucose, 2-hour glucose, glycated hemoglobin (HbA1c), insulin, insulin resistance (homeostatic model assessment for insulin resistance, HOMA-IR)) and lipid (triglyceride, total cholesterol, high-density (HDL) and low-density lipoprotein (LDL) cholesterol) metabolism were identified in our literature survey. Meta-analysis was performed for BMI in schizophrenia, major depressive disorder and bipolar disorder and we found no significant difference in BMI in either of the three patient groups compared to HC (p>0.05). Study heterogeneity was high (*I^2^*>75.7%) across all metabolic markers except BMI, which showed low-to-moderate heterogenity in bipolar disorder and major depressive disorder (*I^2^*<56.1%). Senitivity analysis was performed for BMI in schizophrenia by excluding one influential study and the results remained unchanged.

Meta-analysis was performed for other metabolic markers in schizophrenia only. We found that schizophrenia was associated with significantly increased waist-hip circumference ratio (SMD=0.31, p=0.007), 2-hour glucose (SMD=0.71, p<0.001), insulin (SMD=0.57, p<0.001), insulin resistance (SMD=0.65, p<0.001) and triglycerides (SMD=0.36, p=0.002) and decreased HDL-cholesterol (SMD=-0.34, p=0.025). We did not find significant alteration of fasting glucose, HbA1c, total and LDL-cholesterol in schizophrenia (p>0.05). The significance of the results remained unchanged after excluding outlier/influential studies for insulin (n=1), insulin resistance (n=2), triglycerides (n=2), total chelsterol (n=1), HDL-cholesterol (n=2), LDL-cholesterol (n=1), fasting glucose (n=1) and HbA1c (n=1) in schizophrenia.

Studies of these metabolic markers in other disorder groups were sparse. In one study examining glucose metabolism ^20^, the authors found that both major depressive disorder and bipolar disorder were associated with elevated 2-hour glucose level, but not with fasting glucose or insulin levels which was consistent with another bipolar disorder study ^63^. In another two studies examining lipid metabolism, major depressive disorder was not associated with significant changes in total, HDL or LDL cholesterol or triglyceride levels ^64,65^ and neither was in bipolar disorder ^63^. Waist-hip circumference ratio was not found to differ between major depressive disorder and HC ^66^. We did not find any eligible studies examining metabolic function in generalized anxiety disorder.

### Cardiovascular system

Systolic and diastolic blood pressure were the two most commonly studied cardiovascular markers in the literature; however, meta-analysis was only available for schizophrenia, with estimated high study heterogeneity (*I^2^*>89%). In the meta-analysis, we found that systolic blood pressure was significantly higher in schizophrenia compared to HC (SMD=0.60, p=0.01), but no difference for diastolic blood pressure (p>0.05). We also identified one study ^67^ assessing systolic and diastolic blood pressure in major depressive disorder and they reported no significant between-group differences in the two markers. Sensitivity analysis was performed for systolic blood pressure in schizophrenia after excluding one influential study and the results remained the same.

Other cardiovascular markers including resting heart rate, pulse pressure, left ventricular ejection fraction and heart rate variability were also identified; however, with insufficient studies for meta-analysis. Overall, no significant between-group differences were found for these markers ^68–71^, except a higher pulse pressure in schizophrenia compared to HC that has been reported in one study ^68^, and a lower heart rate range during deep breathing cycles found by another study ^71^ . Amongst other disorder groups few studies examined heart rate variability in major depressive disorder ^72,73^, however, due to the discrepancy in which heart rate variablily was quantified, we were unable to provide a valid synthesis and comparison. Qualitatively, the studies reported mixed variances in HRV across disorders.

### Liver

We identified two blood-derived markers for liver function including albumin and bilirubin in the literature survey. Meta-analysis was only feasible for schizophrenia. Significantly lower albumin levels were found in schizophrenia compared to HC (SMD=-0.74, p<0.001), but no difference for bilirubin (p>0.05). Study heterogeneity was low for albumin (*I^2^*=0%) but high for bilirubin (*I^2^*=86.9%). Narrative review showed mixed findings in bipolar disorder with one study finding lower albumin ^58^ and another finding no differences ^74^, as was found in major depressive disorder ^74^. No between-group differences in bilirubin levels were reported in major depressive disorder ^64,74^ nor bipolar disorder ^74^.

### Musculoskeletal system

Meta-analysis was not feasible for musculoskeletal function related periperal markers. In the narrative synthesis, we identified three studies that investigated bone mineral density in schizophrenia, one study in major depressive disorder, and one in bipolar disorder; across which there was a general pattern of lower bone mineral density in patients. Two studies ^75,76^ found lower bone mineral density across multiple skeleton sites (e.g., lumber spine, left hip, radius, ulna) in schizophrenia, while another ^77^ did not find any significant between-group differences. In major depressive disorder, while Yazici et al (2003) ^78^ found significantly lower bone mineral density at the lumbar spine and all sites of the proximal femur in major depressive disorder compared to HC, this study only included female participants. Similarly, lower bone mineral density was found in bipolar disorder compared to HC ^76^.

## Discussion

We surveyed markers of peripheral organ system health in populations at earlier stages of mental illness, prior to long-term medication exposure. Our analysis indicates that current research is disproportionately focused on inflammatory markers in schizophrenia, followed by major depressive disorder, with relatively few studies examining other peripheral systems or addressing bipolar disorder or generalized anxiety disorder. Elevated peripheral inflammation appears common across schizophrenia and major depressive disorder at early illness stages, but we cannot draw conlcusions for the other two disorders due to the paucity of primary studies. Investigating the function and health of the pulmonary, musculoskeletal, renal and other body systems is a priority area for future research.

### Immune system

Of the 52 markers assessed, only four (IFN-γ, IL-6, neutrophil count, CRP) show alterations across more than one mental illness group. The presence of these four markers across diagnoses may indicate an early-stage immune response and highlight their interconnected roles in the inflammatory cascade. In particular, IL-6 is the central cytokine that drives acute-phase immune response, leading to the production of CRP in liver cells. Once released, CRP facilitates and promotes the recruitment and activation of neutrophils, which is the primary innate immune cells involved in early inflammation, to act on the inflamed tissues. The activated neutrophils will release cytokines (e.g., IL-1β, TNF-α) that further stimulate IL-6 production. This feedback loop is tightly regulated, where excessive IL-6 production will lead to sustained neutrophil activation and CRP elevation and contribute to ongoing tissue damage and chronic inflammation ^79,80^.

While IFN-γ does not directly drive CRP production, it is a central regulator of inflammation that interacts with other cytokines. It also promotes chronic inflammation, accompanied by high CRP and neutrophil recruitment ^81^. Indeed, elevated CRP and neutrophil count are often associated with IL-6 and IFN-γ and thus form a key axis in immune response and resolution ^82^. Our findings thus suggest shared imbalances in this axis across the spectrum of common mental illnesses, which may be modulated by common stress-related proliferation of the immune cell network ^17,18^. While we are unable to synthesize related results in generalized anxiety disorder due to insufficient number of studies, it is speculated that generalized anxiety disorder may share similar pattern of inflammatory dysregulation as reported by Tang and colleagues ^44^. Further research is needed to test this hypothesis.

Our findings also suggest disorder specific inflammatory dysregulation. This is most exemplified by elevated IL-10 in major depressive disorder and IL-4 in schizophrenia. While the exact role and impact of IL-10 and IL-4 in major depressive disorder and schizophrenia is an ongoing research area, the two anti-inflammatory cytokines may be differentially involved in the complex interplay with increased pro-inflammatory cytokines at the onset of major depressive disorder and schizophrenia. Further research into the potential compensatory role of the two cytokines at the early illness stage and how they evolve over the course of illness will be important to enable a better understanding of potential disorder-specific implications. Our meta-analysis also reveals elevated TNF-α in major depressive disorder but no changes in schizophrenia, which is consistent with previous meta-analysis investigating cytokine alterations in first-episode drug-naïve psychosis ^17^ and mood disorders ^23^. However, an earlier meta-analysis ^15^ found elevated TNF-α in both schizophrenia and depression patients. It is worth noting that while our study focuses on adult-onset mental illness, the meta-analysis by Çakici and colleagues ^15^ included a mixed sample of adolescent and adult participants. The extent to which TNF-α dysregulation is a signature of early-onset psychosis remains to be addressed. Other potential disorder-specific alterations in cytokines including elevated IL-1α, IL-23 and TGF-β in schizophrenia, and elevated IL2 and IL-17 in major depressive disorder remain inconclusive due to limited number of studies.

### Metabolic system

Of the 11 metabolic markers assessed, BMI was the only marker available for meta-analysis in more than one disorder groups. While we did not find significantly elevated BMI in patients with schizophrenia, major depressive disorder or bipolar disorder, this result may be influenced by the selection biases inherent to many studies in psychiatic disorders as they often exclude people with high BMI or recruit partipicants with matching BMI. Given that BMI is a proxy for metabolic health and that elevated BMI is an established risk factor for type 2 diabetes ^83^, future research including a more naturalistic sample would be needed to assess the relationship between BMI and metabolic disease risk in people with mental illness. Nevertheless, assessment of other metabolic markers reveals elevated waist-hip circumference ratio, fasting glucose, 2-hour plasma glucose, insulin, insulin resistance and triglycerides, and decreased HDL cholesterol in schizophrenia with small to moderate effect sizes. Our findings are consistent with recent literature investigating metabolic dysregulation in schizophrenia, with this pattern of abnormalities suggesting that patients with schizophrenia may be already at increased risk of type 2 diabetes and metabolic syndrome ^13–16,29^ at the onset of illness without exposure to medication side effects. We also find some evidence that suggests abnormal glucose metabolism in bipolar disorder and major depressive disorder as indicated by elevated 2-hour glucose level, but not other metabolic markers ^20,63–66^. However, due to limited number of studies, further research is needed to assess metabolic function in these disorders.

### Other body systems

For other body systems including cardiovascular, liver and musculoskeletal systems, evidence is overall limited for comparisons across disorder groups. For the cardiovascular system, we find significantly elevated systolic blood pressure in patients with schizophrenia, which is thought to be associated with increased risk of autonomic dysregulation and cardiovascular disease ^84,85^. Review of studies on liver function are inconclusive, with mixed findings reported across studies and across disorder groups, except for significantly decreased albumin levels in patients with schizophrenia. Decreased albumin may be explained by poor diet commonly found in people with psychiatric disorders ^86^. However, previous studies also suggest the presence of hypoalbuminemia independent of malnutrition in these patients ^87^. Indeed, albumin is an antioxidant which is implicated in a protective role for inflammation. It is thus possible that oxidative dysfunction reflected by decreased albumin in patients with schizophrenia may be also associated with increased vulnerability to inflammation and psychiatric symptoms ^88,89^. Regarding musculoskeletal health, we find some evidence that suggests decreased bone mineral density in schizophrenia, bipolar disorder and major depressive disorder compared to healthy individuals ^75,76,78^. While a previous meta-analysis suggests that prolactin-raising antipsychotics may induce bone mineral density loss in chronic schizophrenia ^90^, our findings provide some preliminary evidence that decreased bone mineral density may be already present at illness onset before antipsychotic treatment, and thus suggests an inherent risk of osteoporosis and fracture associated with psychiatric disorders. We did not consider hormonal markers such as cortisol, thyroid or sex hormones due to their regulatory role over organ systems and their direct interactions with the brain through the hypothalamus-pituitary axis. It will be important for future meta-analyses to consider hormonal markers in the early stages of illness onset, given that altered hormonal activity during development or in adulthood may be associated with psychiatric symptoms ^21,91,92^.

### Limitations: Heterogeneity, biases and sparsity of studies

Overall, we find that most studies focussed on schizophrenia, followed by depression. Studies on bipolar disorder and generalized anxiety disorder are limited in the literature despite growing evidence suggesting that similar patterns of physical health manifestation may be common across disorders ^1,12^. Amongst physiological markers examined, inflammatory markers are the most commonly and widely studied category, followed by a substantial reduction in studies of metabolic, cardiovascular, liver and musculoskeletal function. We did not identify any studies of lung and renal function. Previous literature in the field of biological psychiatry suggests that inflammatory markers may provide the best transdiagnostic indication of physical health dysfunction across common psychiatric disorders ^93,94^. However, this may in part be attributed to the lack of studies investigating the role of other peripheral systems across illnesses contributing to ultimately inconclusive findings, as marked in prior reviews ^93^.

It is important to highlight that our review reveal significant heterogeneity across studies, which is known to be a common source of bias within the literature ^93,95,96^. We highlight the significant heterogeneity in study protocols, methodology and reporting of results. For example, individual studies utilized different sample collection procedures and assay kits from varying manufacturers. Participant selection is not entirely standardized in controlling for potential confounding factors such as BMI and lifestyle factors including smoking status, diet and physical activity, and age of illness onset or illness duration, the latter of which resulted in a large proportion of studies excluded from review. Hence, when interpreting these findings, it is important to bear in mind the potential effects of these and other unmeasured potential confounds, including lifestyle, diet and demographic factors. This also underscores the importance of improved reporting of these potential confounding factors, which are often unavailable. Given significant heterogeneity of both between-study methodology and findings in the literature, our results should be interpreted with caution and demonstrate further need for improved standardization in the study of peripheral biomarkers, guidelines for which have recently been proposed to improve study validity ^97^. Moreover, due to the lack of symptom severity measures in some studies and the variation in symptom scales used across studies, we were unable to examine the extent to which heterogeneity in findings of physiological markers may be explained by variation in symptom severity across cohorts. In addition, although a minimal of 3 studies is sufficient to run a meta-analysis, the robustness of the estimates can be limited when there is only a small number of studies. Further caution is therefore needed in interpreting the results.

### Conclusions

In conclusion, our review provides a comprehensive characterization of the state of the literature examining peripheral organ function in four common mental illnesses. Our findings suggest that psychiatric illness, at least schizophrenia, is likely is associated with altered peripheral organ function across multiple systems at illness onset. Yet, further evidence is needed to draw firm conclusion on the extent to which this pattern of body system dysregulation may be shared across the four common psychiatric disorders studied. We highlight the significant sparsity in research of peripheral organ function in drug naïve first-episode psychiatric patients other than immune and metabolic function in schizophrenia and related psychosis. Further research of other peripheral organ systems across the spectrum of mental disorders is needed to enable identification of individuals at risk of physical comorbidity at the very start of mental health care.

## Data Availability

This is a meta-analysis. All data produced in the present study are available from respective studies and are contained in the manuscript.

## Acknowledgement

This study was supported by a National Health and Medical Research Council Investigator Grant awarded to Y.E.T (APP2016413). The funders had no role in study design, data collection, data analysis, data interpretation, or writing of the report. The corresponding author had full access to all the data in the study and had final responsibility for the decision to submit for publication.

