## Supplementary Information for "Altered function of peripheral organ systems in first-episode drug naïve mental illnesses: A systematic review and meta-analysis"

[+]These authors contributed equally

### Table of Content

Supplementary Figure 1. Overlap in study inclusion between organ systems.

Supplementary Table 1. PRISMA 2020 checklist.

Supplementary Table 2. Search strategy (EMBASE, MEDLINE, PsycINFO).

Supplementary Table 3. Included studies.

Supplementary Table 4. Overlap in study cohorts.

Supplementary Table 5. Excluded studies at full text screening.

Supplementary Table 6. JBI risk of bias critical appraisal checklist for case control studies.

Supplementary Table 7. Meta-analysis raw data of pro-inflammatory biomarkers in schizophrenia.

Supplementary Table 8 Meta-analysis raw data of anti-inflammatory biomarkers in schizophrenia.

Supplementary Table 9. Meta-analysis raw data of adaptive-inflammatory biomarkers in schizophrenia.

Supplementary Table 10. Meta-analysis raw data of other inflammatory biomarkers in schizophrenia.

Supplementary Table 11. Meta-analysis raw data of BMI in schizophrenia.

Supplementary Table 12. Meta-analysis raw data of glucose metabolism in schizophrenia.

Supplementary Table 13. Meta-analysis raw data of lipid metabolism in schizophrenia.

Supplementary Table 14. Meta-analysis raw data of liver function in schizophrenia.

Supplementary Table 15. Meta-analysis raw data of cardiovascular function in schizophrenia.

Supplementary Table 16. Meta-analysis raw data of pro-inflammatory biomarkers in major depressive disorder.

Supplementary Table 17. Meta-analysis raw data of anti-inflammatory biomarkers in major depressive disorder.

Supplementary Table 18. Meta-analysis raw data of other inflammatory biomarkers in major depressive disorder.

Supplementary Table 19. Meta-analysis raw data of BMI in major depressive disorder.

Supplementary Table 20. Meta-analysis raw data of BMI in bipolar disorder.

Supplementary Table 21. Meta-analysis excluding studies that included patients with ambiguous number of episodes.

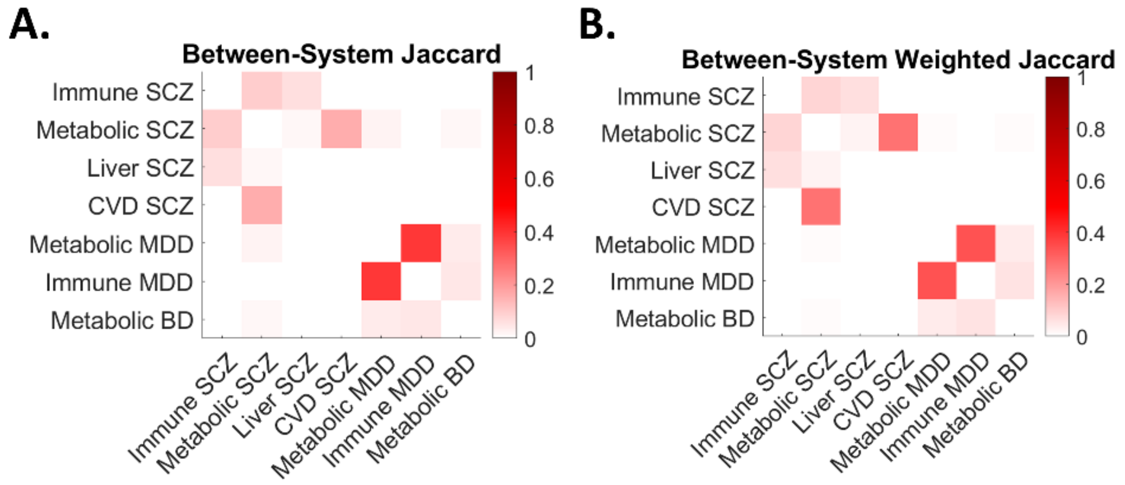


**Supplementary Figure 1. Overlap in study inclusion between organ systems. A**. Heatmaps show between-system Jaccard coefficient for all pairs of organs/systems across all disease groups. **B**. Sample size weighted Jaccard coefficient. SCZ, schizophrenia; MDD, major depressive disorder; BD, bipolar disorder; CVD, cardiovascular.

**Supplementary Table 1. PRISMA 2020 checklist.**

| Section and Topic | Item # | Checklist item | Location where item is reported |
|---|---|---|---|
| **TITLE** | | | |
| Title | 1 | Identify the report as a systematic review. | 1 |
| **ABSTRACT** | | | |
| Abstract | 2 | See the PRISMA 2020 for Abstracts checklist. | 2 |
| **INTRODUCTION** | | | |
| Rationale | 3 | Describe the rationale for the review in the context of existing knowledge. | 3 |
| Objectives | 4 | Provide an explicit statement of the objective(s) or question(s) the review addresses. | 3 |
| **METHODS** | | | |
| Eligibility criteria | 5 | Specify the inclusion and exclusion criteria for the review and how studies were grouped for the syntheses. | 3,4 |
| Information sources | 6 | Specify all databases, registers, websites, organisations, reference lists and other sources searched or consulted to identify studies. Specify the date when each source was last searched or consulted. | 3 |
| Search strategy | 7 | Present the full search strategies for all databases, registers and websites, including any filters and limits used. | 4 |
| Selection process | 8 | Specify the methods used to decide whether a study met the inclusion criteria of the review, including how many reviewers screened each record and each report retrieved, whether they worked independently, and if applicable, details of automation tools used in the process. | 4 |
| Data collection process | 9 | Specify the methods used to collect data from reports, including how many reviewers collected data from each report, whether they worked independently, any processes for obtaining or confirming data from study investigators, and if applicable, details of automation tools used in the process. | 4 |
| Data items | 10a | List and define all outcomes for which data were sought. Specify whether all results that were compatible with each outcome domain in each study were sought (e.g. for all measures, time points, analyses), and if not, the methods used to decide which results to collect. | 4 |
| | 10b | List and define all other variables for which data were sought (e.g. participant and intervention characteristics, | 4 |

| Section and Topic | Item # | Checklist item | Location where item is reported |
| --- | --- | --- | --- |
| | | funding sources). Describe any assumptions made about any missing or unclear information. | |
| Study risk of bias assessment | 11 | Specify the methods used to assess risk of bias in the included studies, including details of the tool(s) used, how many reviewers assessed each study and whether they worked independently, and if applicable, details of automation tools used in the process. | 4 |
| Effect measures | 12 | Specify for each outcome the effect measure(s) (e.g. risk ratio, mean difference) used in the synthesis or presentation of results. | 4 |
| Synthesis methods | 13a | Describe the processes used to decide which studies were eligible for each synthesis (e.g. tabulating the study intervention characteristics and comparing against the planned groups for each synthesis (item #5)). | 4 |
| | 13b | Describe any methods required to prepare the data for presentation or synthesis, such as handling of missing summary statistics, or data conversions. | 4 |
| | 13c | Describe any methods used to tabulate or visually display results of individual studies and syntheses. | 4,5 |
| | 13d | Describe any methods used to synthesize results and provide a rationale for the choice(s). If meta-analysis was performed, describe the model(s), method(s) to identify the presence and extent of statistical heterogeneity, and software package(s) used. | 4,5 |
| | 13e | Describe any methods used to explore possible causes of heterogeneity among study results (e.g. subgroup analysis, meta-regression). | 5 |
| | 13f | Describe any sensitivity analyses conducted to assess robustness of the synthesized results. | 5 |
| Reporting bias assessment | 14 | Describe any methods used to assess risk of bias due to missing results in a synthesis (arising from reporting biases). | 4,5 |
| Certainty assessment | 15 | Describe any methods used to assess certainty (or confidence) in the body of evidence for an outcome. | 5 |
| **RESULTS** | | | |
| Study selection | 16a | Describe the results of the search and selection process, from the number of records identified in the search to the number of studies included in the review, ideally using a flow diagram. | 5 |
| | 16b | Cite studies that might appear to meet the inclusion criteria, but which were excluded, and explain why they were | 5 |

| Section and Topic | Item # | Checklist item | Location where item is reported |
|---|---|---|---|
| | | excluded. | |
| Study characteristics | 17 | Cite each included study and present its characteristics. | 5 |
| Risk of bias in studies | 18 | Present assessments of risk of bias for each included study. | 5 |
| Results of individual studies | 19 | For all outcomes, present, for each study: (a) summary statistics for each group (where appropriate) and (b) an effect estimate and its precision (e.g. confidence/credible interval), ideally using structured tables or plots. | 5,6 |
| Results of syntheses | 20a | For each synthesis, briefly summarise the characteristics and risk of bias among contributing studies. | 5 |
| | 20b | Present results of all statistical syntheses conducted. If meta-analysis was done, present for each the summary estimate and its precision (e.g. confidence/credible interval) and measures of statistical heterogeneity. If comparing groups, describe the direction of the effect. | 6-9 |
| | 20c | Present results of all investigations of possible causes of heterogeneity among study results. | 6-9 |
| | 20d | Present results of all sensitivity analyses conducted to assess the robustness of the synthesized results. | 6-9 |
| Reporting biases | 21 | Present assessments of risk of bias due to missing results (arising from reporting biases) for each synthesis assessed. | 5 |
| Certainty of evidence | 22 | Present assessments of certainty (or confidence) in the body of evidence for each outcome assessed. | 6-9 |
| **DISCUSSION** | | | |
| Discussion | 23a | Provide a general interpretation of the results in the context of other evidence. | 9-11 |
| | 23b | Discuss any limitations of the evidence included in the review. | 10,11 |
| | 23c | Discuss any limitations of the review processes used. | 10,11 |
| | 23d | Discuss implications of the results for practice, policy, and future research. | 9-11 |
| **OTHER INFORMATION** | | | |
| Registration | 24a | Provide registration information for the review, including register name and registration number, or state that the | 5 |

| Section and Topic | Item # | Checklist item | Location where item is reported |
|---|---|---|---|
| and protocol | | review was not registered. | |
| | 24b | Indicate where the review protocol can be accessed, or state that a protocol was not prepared. | 5 |
| | 24c | Describe and explain any amendments to information provided at registration or in the protocol. | 5 |
| Support | 25 | Describe sources of financial or non-financial support for the review, and the role of the funders or sponsors in the review. | 12 |
| Competing interests | 26 | Declare any competing interests of review authors. | 12 |
| Availability of data, code and other materials | 27 | Report which of the following are publicly available and where they can be found: template data collection forms; data extracted from included studies; data used for all analyses; analytic code; any other materials used in the review. | 6 |

*From:* Page MJ, McKenzie JE, Bossuyt PM, Boutron I, Hoffmann TC, Mulrow CD, et al. The PRISMA 2020 statement: an updated guideline for reporting systematic reviews. BMJ 2021;372:n71. doi: 10.1136/bmj.n71. This work is licensed under CC BY 4.0. To view a copy of this license, visit https://creativecommons.org/licenses/by/4.0/

**Supplementary Table 2. Search strategy (EMBASE, MEDLINE, PsycINFO).**

| Search | Query |
|---|---|
| 1. | Psychotic Disorders/ or Schizophrenia/ or (schizophren* or schizoaffective).ti,ab. |
| 2. | Bipolar Disorder/ or (bipolar or manic or mania).ti,ab. |
| 3. | Depressive Disorder, Major/ or (major depressive disorder* or major depression disorder* or clinical depression).ti,ab. |
| 4. | Anxiety Disorders/ or (generalized anxiety disorder* or generalised anxiety disorder* or anxiety disorder*).ti,ab. |
| 5. | (drug naive or drug-naive or antipsychotic naive or antipsychotic-naive or antipsychotic-free or antipsychotic free or early-onset or early onset or first episode or first-episode or first presentation* or first manifestation or early stage*).mp. |
| 6 | (HbA1c or Hba1c or HbA1C or HOMA-IR or TyG index or plasma insulin or plasma glucose or OGTT or oral glucose tolerance).mp. or Glucose Tolerance Test/ |
| 7. | (LDL or HDL or triglyceride* or cholesterol* or BMI or waist circumference* or skinfold thickness).mp. |
| 8. | ((systolic adj2 blood) or (diastolic adj2 blood) or rate variability or HRV or CMR or cardiac index or cardiac output or stroke volume or systolic volume or diastolic volume or ejection fraction or blood pressure or echocardiog* or TOE or TTE).mp. |
| 9. | (creatinine or urea or GFR or eGFR or glomerular filtration).mp. |
| 10. | (bilirubin* or GGT or AST or ALT or ALP or albumin* or phosphatase* or transferase* or transaminase*).mp. |
| 11. | (absorptiometry or DEXA or DEX or bone densit* or BUA or ultrasound attenuation or stiff index or speed sound).mp. |
| 12. | (musculoskeletal or FGC or walking capacit* or 6MWT or broad jump or muscular fitness).mp. |
| 13. | (spiromet* or FVC or FEV or tidal volume* or lung function test*).mp. |
| 14. | (cytokine* or leyukotrine* or leukotriene* or IL-1* or IL-2* or IL-6* or interleukin* or CRP or ESR or TGF or TNF or interferon* or C-reactive or necrosis factor or growth factor or reactive-c or hsCRP or neutrophil* or leukocyte*).mp. |
| 15. | 1 or 2 or 3 or 4 |
| 16. | 6 or 7 or 8 or 9 or 10 or 11 or 12 or 13 or 14 |
| 17. | 5 and 15 and 16 |
| Limits | English Language, Humans |

**Supplementary Table 3. Included studies.**

| Study | Country | Diagnosis | Patients (n) | Patients Sex *M/F* | Patients Age *Mean (SD)* | Controls (n) | Controls Sex *M/F* | Controls Age *Mean (SD)* | Outcomes |
|---|---|---|---|---|---|---|---|---|---|
| Almis and Egilmez, 2021 [1]<br>Kapici et al, 2023 [2] | Turkey | Schizophrenia | 72 | 44/28 | 30.35 (9.09) | 64 | 29/35 | 31.23 (10.62) | CRP, Fasting glucose, HDL, LDL, Lymphocyte, Monocyte, Neutrophil, NLR, Platelet, PLR, RHR, Total cholesterol, Triglycerides, WBC |
| | | Bipolar | 63 | 31/32 | 27.92 (8.33) | | | | Platelet |
| Amoli et al, 2019 [3] | Iran | Schizophrenia | 15 | 8/7 | 34.24 (7.86) | 15 | 8/7 | 33.93 (4.78) | BMI |
| Bai et al, 2021 [4] | China | Depression | 53 | 20/33 | 32.34 (14.43) | 86 | 34/52 | 37.35 (13.94) | BMI |
| Bolu et al, 2019 [5] | Turkey | Schizophrenia | 48 | 48/0 | 23.5 (3.5) | 54 | 54/0 | 36.3 (12.5) | BMI, CRP |
| Borovcanin et al, 2012; 2013; 2015 [6-8] | Serbia | Schizophrenia | 88 | 36/52 | 33.64 (8.84) | 36 | 36/0 | 35.61 (10.44) | IFN-γ, IL-4, IL-6, IL-17, IL-23, IL-27, TGF-b |
| Cabello-Rangel et al, 2023 [9] | Mexico | Schizophrenia | 175 | 132/43 | 29.06 (10.1) | 52 | 14/38 | 33.85 (10.9) | Lymphocyte, Neutrophil, NLR, Platelet |
| Chang et al, 2017 [10] | Taiwan | Depression | 72 | 21/51 | 40.72 (12.45) | 96 | 41/55 | 33.30 (12.11) | BMI, CRP |
| | | Bipolar | 88 | 44/44 | 31.74 (11.78) | | | | |

| | | | | | | | | | |
|---|---|---|---|---|---|---|---|---|---|
| Chen et al, 2016 [11] | China | Schizophrenia | 172 | 83/89 | 28.7 (9.9) | 31 | 14/17 | 26.9 (5.2) | Fasting glucose, 2-hour fasting glucose, HDL, Insulin, Insulin resistance, LDL, Total cholesterol, Triglycerides |
| Chen et al, 2018 [12] | China | Schizophrenia | 100 | 53/47 | 25.51 (8.27) | 118 | 61/57 | 24.61 (3.10) | BMI, Fasting glucose, 2-hour fasting glucose, HDL, Insulin, Insulin resistance, LDL, Total cholesterol, Triglycerides, WHR |
| Chen et al, 2021a [13] | China | Schizophrenia | 113 | 113/0 | 29.2 (9.29) | 58 | 58/0 | 30.22 (6.4) | IL-17, WHR |
| Chen et al, 2021b [14] | China | Schizophrenia | 24 | 13/11 | 26.46 (6.487) | 25 | 10/15 | 27 (5.132) | IFN-γ, IL-1β, IL-2, IL-4, IL-5, IL-6, IL-7, IL-8, IL-10, IL-12, IL-13, IL-15, TNF-α |
| Chen et al, 2022a [15] | China | Depression | 29 | 5/24 | 34.38 (14.83) | 25 | 2/23 | 35 (13.91) | CRP, IL-6, TNF-α |
| Chen et al, 2022b [16] | China | Bipolar | 191 | 96/95 | 32.24 (11.68) | 112 | 56/56 | 33.04 (11.64) | BMI, CRP |

| | | | | | | | | |
|---|---|---|---|---|---|---|---|---|
| Chen et al, 2024 [17] | China | Schizophrenia | 52 | 27/25 | Median 25 | 52 | 28/24 | Median 25.1 | HDL, LDL, Lymphocyte, Monocyte, Neutrophil, Platelet, Total cholesterol, Triglycerides, WBC |
| Cropley et al, 2023 [18] | Germany | Schizophrenia | 24 | 20/4 | 30.3 (10.1) | 25 | 12/13 | 28.9 (9.8) | Albumin, BMI, Fasting glucose |
| Dai et al, 2020 [19] | China | Schizophrenia | 46 | 25/21 | 23.2 (6.9) | 60 | 32/28 | 24.6 (3.8) | BMI, IL-1β, IL-6 |
| Dasgupta et al, 2010 [20] | India | Schizophrenia | 30 | 14/16 | 32.53 (10.53) | 25 | 12/13 | 35.68 (9.57) | BMI, Fasting glucose, HDL, Insulin resistance, LDL, Total cholesterol, Triglycerides |
| De Berardis et al, 2013 [21] | Italy | Schizophrenia | 30 | 13/17 | 25.9 (6) | 30 | NR | NR | CRP |
| de Menezes Galvao et al, 2021 [22] | Brazil | Depression | 30 | 14/16 | 24.2 (3.84) | 32 | 15/17 | 27.06 (6.42) | BMI |
| Ding et al, 2014 [23] | China | Schizophrenia | 69 | 37/32 | 27.48 (7.75) | 60 | 27/33 | 26.82 (4.38) | IFN-γ, IL-6, IL-17 |
| Dongxia et al, 2024 [24] | China | Schizophrenia | 123 | 65/58 | 36.72 (8.18) | 38 | 22/16 | 37.35 (5.53) | HDL, LDL, Total cholesterol, Triglycerides |
| Fernandez-Egea et al, 2009a; 2009b [25,26] Garcia-Rizo et al, 2016 [27] Kirkpatrick et al, | Italy/Spain | Schizophrenia | 50 | 35/15 | 29.4 (8.8) | 50 | 35/15 | 28.8 (7.7) | CRP, DBP, Fasting glucose, 2-hour fasting glucose, HbA1c, HDL, IL-6, Insulin, Insulin resistance, LDL, |

| | | | | | | | | | |
|---|---|---|---|---|---|---|---|---|---|
| 2009 [28] | | | | | | | | | Pulse pressure, RHR, SBP, Total cholesterol, Triglycerides |
| Kirkpatrick et al, 2010 [29] | | | | | | | | | Fasting glucose, 2-hour fasting glucose, Insulin |
| | | Depression | 12 | 7/5 | 26.5 (5.6) | 98 | 61/37 | 26.66 (4.3) | Fasting glucose, 2-hour fasting glucose, Insulin |
| | | Bipolar | 6 | 5/1 | 29.7 (4.9) | | | | |
| Frydecka et al, 2018 [30] | Poland | Schizophrenia | 39 | 19/20 | 25.5 (5.1) | 39 | 19/20 | 26 (2.6) | IFN-γ, IL-1β, IL-2, IL-4, IL-5, IL-6, IL-7, IL-8, IL-9, IL-10, IL-12, IL-13, TNF-α |
| Gao et al, 2024 [31] | China | Depression | 23 | 15/8 | 39.87 (15.80) | 23 | 15/8 | 38.91 (10.38) | BMI, IFN-γ, IL-1β, IL-2, IL-6, IL-8, IL-12, TNF-α |
| Garcia-Rizo et al, 2019 [32] | Spain | Schizophrenia | 38 | 24/14 | 27.94 (6.7) | 49 | 28/21 | 26.17 (5) | Lymphocyte, Monocyte, Neutrophil, NLR, WBC |
| Garrido-Torres et al, 2022 [33] | Spain | Schizophrenia | 244 | 117/96 | 32 (10) | 166 | 151/15 | 29.6 (7.9) | BMI, DBP, Fasting glucose, HDL, SBP, Triglycerides |
| Goyal et al, 2020 [34] | India | Bipolar | 40 | 20/20 | 25.2 (5.04) | 40 | 20/20 | 26.7 (4.19) | Lymphocyte, Neutrophil, NLR, Platelet, PLR |

| | | | | | | | | | |
|---|---|---|---|---|---|---|---|---|---|
| Haring et al, 2015 [35]<br>Balotsev et al, 2019 [36]<br>Parksepp et al, 2022 [37]<br>Korhonen et al, 2023 [38]<br>Kuuskmae et al, 2023 [39] | Estonia | Schizophrenia | 38 | 21/17 | 25.4 (0.89) | 37 | 16/21 | 24.8 (0.86) | BMI, IFN-γ, IL-1α, IL-1β, IL-2, IL-4, IL-6, IL-8, IL-10, Insulin, TNF-α |
| Herran et al, 2000 [40] | Spain | Depression | 19 | 0/19 | 44.7 (12.1) | 19 | 0/19 | 43.5 (8.4) | BMI |
| Huang et al, 2022 [41] | China | Schizophrenia | 53 | 27/26 | 22.04 (8.7) | 59 | 30/29 | 35.56 (10.33) | Albumin, Bilirubin, Lymphocyte, MLR, Monocyte, Neutrophil, NLR, Platelet, PLR, WBC |
| Joaquim et al, 2018 [42] | Brazil | Schizophrenia | 28 | 17/11 | 26.0 (7.4) | 30 | 16/14 | 26.2 (3.9) | IL-1β |
| Jordan et al, 2018 [43]<br>Kapogiannis et al, 2019 [44]<br>Savukoski et al, 2024 [45]<br>Steiner et al, 2020 [46] | Germany | Schizophrenia | 22 | 13/9 | 30.00 (25.00, 35.50) *Median (IQR)* | 43 | 26/17 | 35.00 (26.00, 45.00) *Median (IQR)* | BMI, CRP, Fasting glucose, Insulin, Insulin resistance, Lymphocyte, MLR, Monocyte, Neutrophil, NLR |
| Kakeda et al, 2018; 2020 [47,48] | Japan | Depression | 40 | 20/20 | 46.6 (14.4) | 47 | 34/13 | 40.7 (11.4) | IFN-γ, IL-1β, IL-6, TNF-α |
| Kalmady et al, 2018 [49] | India | Schizophrenia | 75 | 41/34 | 30.69 (6.52) | 102 | 57/45 | 25.78 (4.75) | BMI, IFN-γ, IL-2, IL-4, IL-6, IL-10, IL-17, TNF-α |

| | | | | | | | | | |
|---|---|---|---|---|---|---|---|---|---|
| Keri et al, 2014 [50] | Hungary | Depression | 50 | 19/31 | 22.6 (6.0) | 30 | 9/21 | 24.3 (8.3) | BMI, CRP, IL-6, WHR |
| Keri et al, 2017 [51] | Hungary | Schizophrenia | 42 | 29/13 | 26.1 (6.8) | 42 | 29/13 | 26.2 (5.9) | BMI, WHR |
| Khan, 2022 [52] | US | Schizophrenia | 22 | 22/0 | 23.54 (4.65) | 14 | 14/0 | 25 (7.6) | BMI |
| Kim et al, 2021 [53] | Korea | Depression | 50 | 23/27 | 24.18 (3.74) | 50 | 24/26 | 24.8 (3.22) | BMI, CRP, IL-1β, IL-6, IL-17, TNF-α |
| Kolenic et al, 2018 [54] | Czech Republic | Schizophrenia | 120 | 74/46 | 27.00 (4.94) | 114 | 53/61 | 25.70 (4.01) | BMI, HDL, LDL, Triglycerides |
| Kuwano et al, 2018 [55] | Japan | Depression | 15 | 9/6 | 30.1 (7.6) | 19 | 10/9 | 30.2 (6.8) | Bilirubin, CRP, HDL, LDL, Total cholesterol |
| Lan et al, 2021; 2022 [56,57] | China | Depression | 54 | 25/29 | 30.74 (10.96) | 60 | 35/25 | 31.57 (10.74) | BMI, IFN-γ, IL-1β, IL-2, IL-4, IL-5, IL-6, IL-7, IL-8, IL-10, IL-13, IL-17, IL-23, TNF-α |
| Lang et al, 2021 [58] Li et al, 2021 [59] | China | Schizophrenia | 430 | 208/222 | 32.72 (11.26) | 453 | 197/256 | 33.24 (11.49) | BMI, DBP, Fasting glucose, HbA1c, HDL, Insulin, Insulin resistance, LDL, SBP, Total cholesterol, Triglycerides |
| Leo et al, 2006 [60] | Italy | Depression | 46 | 20/26 | 34.85 (5.88) | 46 | 19/27 | 34.11 (5.22) | BMI, IL-1β, IL-6, LDL, TNF-α, Total cholesterol, Triglycerides |
| Li et al, 2013 [61] | China | Depression | 64 | 11/53 | 32.1 (6.8) | 64 | 14/50 | 31.6 (5.9) | BMI, TNF-α |

| | | | | | | | | | |
|---|---|---|---|---|---|---|---|---|---|
| Li et al, 2022a [62] | China | Schizophrenia | 83 | 83/0 | 26.9 (9.12) | 14 | 14/0 | 28.86 (5.96) | BMI, Fasting glucose, 2-hour fasting glucose, Total cholesterol, Triglycerides, HDL, LDL |
| Li et al, 2024a [63] | China | Schizophrenia | 56 | 21/35 | 24.31 (5.10) | 108 | 47/61 | 23.34 (2.72) | BMD, BMI |
| | | Bipolar | 130 | 46/84 | 22.96 (3.83) | | | | |
| Li et al, 2024b [64]<br>Su et al, 2023 [65]<br>Sun et al, 2024 [66]<br>Wang et al, 2025 [67] | China | Schizophrenia | 137 | 50/87 | 36.47 (10.46) | 67 | 30/37 | 35.85 (8.59) | BMI, IFN-γ |
| | | Depression | 237 | 68/169 | 37.26 (12.94) | 89 | 45/44 | 27.09 (8.46) | BMI, IL-6 |
| Li et al, 2025 [68] | China | Schizophrenia | 89 | 0/89 | 30.76 (10.40) | 17 | 0/17 | 25.35 (4.03) | BMI, Fasting glucose, 2-hour fasting glucose, HDL, Insulin, Insulin resistance, LDL, Total cholesterol, Triglycerides, WHR |
| | | | 83 | 83/0 | 26.90 (9.12) | 14 | 14/0 | 28.86 (5.96) | |
| Liang et al, 2015 [69] | Taiwan | Depression | 156 | 156/0 | 24.2 (3) | 49 | 49/0 | 24.7 (1.5) | BMI, RHR |

| | | | | | | | | | |
|---|---|---|---|---|---|---|---|---|---|
| Liang et al, 2019 [70] | China | Schizophrenia | 49 | 21/28 | 24.8 (4.9) | 71 | 31/40 | 25.9 (4.7) | BMD, BMI, Fasting glucose, HDL, Insulin, LDL, Total cholesterol, Triglycerides |
| Liang et al, 2022 [71] | China | Schizophrenia | 92 | 40/52 | 27.09 (12.44) | 59 | 25/34 | 27.11 (7.76) | BMI, Fasting glucose, HDL, LDL, Total cholesterol, Triglycerides |
| Lin et al, 2021 [72] | China | Schizophrenia | 51 | 31/19 | 27.51 (8.71) | 114 | 21/93 | 44.31 (14.40) | TNF-α |
| Liu et al, 2015 [73] | China | Schizophrenia | 35 | 14/21 | 32.49 (14.12) | 35 | 18/17 | 36.54 (5.98) | BMI |
| | | Depression | 35 | 17/18 | 36.40 (10.65) | | | | |
| Liu et al, 2022 [74] | China | Depression | 66 | 27/39 | 24.20 (9.60) | 43 | 20/23 | 23.67 (3.19) | BMI, CRP, IL-1β, IL-6, IL-10, TNF-α |
| Liu et al, 2023 [75] | China | Bipolar | 66 | NR | 23 (19, 27) | 66 | NR | 24.5 (22, 26) | BMI, Fasting glucose, HDL, Insulin, Insulin resistance, LDL, Total cholesterol, Triglycerides |
| Milleit et al, 2019 [76] | Germany | Schizophrenia | 18 | 12/13 | 28.1 (7.5) | 25 | 12/13 | 27.4 (7.5) | IFN-γ, IL-4, IL-6, IL-8, IL-13, TNF-α |
| Misiak et al, 2014 [77] | Poland | Schizophrenia | 35 | 11/24 | 29.22 (4.50) | 53 | 26/27 | 25.68 (2.89) | Fasting glucose, HDL, LDL, Total cholesterol, Triglycerides |
| Misiak et al, 2016 [78] | Poland | Schizophrenia | 135 | 75/60 | 27.2 (6.5) | 146 | 67/79 | 27.6 (5.5) | BMI, DBP, Fasting glucose, HDL, LDL, SBP, Total |

| | | | | | | | | | |
|---|---|---|---|---|---|---|---|---|---|
| | | | | | | | | | cholesterol, Triglycerides |
| Ntouros et al, 2018 [79] | Greece | Schizophrenia | 25 | 25/0 | 25.48 (5.41) | 23 | 25/0 | 27.04 (2.91) | IFN-γ, IL-1β, IL-2, IL-4, IL-5, IL-8, IL-10, IL-12, TNF-α, TNF-b |
| Onur et al, 2021 [80] | Turkey | Schizophrenia | 60 | 38/22 | 28.3 (NR) | 32 | 17/15 | 29.2 (NR) | Lymphocyte, MLR, Monocyte, Neutrophil, NLR, Platelet, PLR, WBC |
| Pan et al, 2022 [81] | China | Schizophrenia | 75 | 34/41 | 28.61 (6.90) | 44 | 24/40 | 30.07 (7.49) | BMI, TGF-b1 |
| Petrikis et al, 2015a; 2015b; 2016 [82-84] | Greece | Schizophrenia | 40 | 27/13 | 32.45 (9.81) | 40 | 25/15 | 31.90 (8.29) | Fasting glucose, HbA1c, HDL, IL-2, IL-6, IL-10, IL-17, Insulin, Insulin resistance, TGF-b2, Total cholesterol, Triglycerides |
| Puangsri and Ninla-Aesong, 2021 [85] | Thailand | Depression | 137 | 41/96 | 20.0 (1.3) *Median (SD)* | 56 | 20/36 | 20.0 (1.1) *Median (SD)* | BMI, Lymphocyte, MLR, Monocyte, Neutrophil, NLR, Platelet, PLR, WBC |
| Radu et al, 2020 [86] | Romania | Schizophrenia | 50 | 23/27 | 29.7 (6.6) | 50 | NR | NR | DBP, Fasting glucose, HDL, LDL, LVEF, SBP, Triglycerides |
| Reddy et al, 2003 [87] | United States | Schizophrenia | 31 | 20/11 | 28.5 (7.8) | 40 | 29/11 | 27.9 (8) | Albumin, Bilirubin |
| | | Depression & Bipolar | 12 | 7/5 | 25.3 (7.5) | | | | |

| | | | | | | | | | |
|---|---|---|---|---|---|---|---|---|---|
| Ryan et al, 2003 [88] | Ireland | Schizophrenia | 26 | 15/11 | 33.6 (13.5) | 26 | 15/11 | 34.4 (1.9) | BMI, Fasting glucose, HDL, Insulin, Insulin resistance, LDL, Total cholesterol, Triglycerides |
| Saddichha et al, 2008a; 2008b; 2008c [89-91] | India | Schizophrenia | 99 | 52/47 | 26.06 (5.57) | 51 | 30/21 | 27.5 (5.9) | BMI, DBP, Fasting glucose, 2-hour fasting glucose, HDL, SBP, Triglycerides |
| Sahpolat et al, 2022 [92] | Turkey | Schizophrenia | 37 | 20/17 | 29.73 (11.23) | 43 | 23/20 | 29.32 (7.71) | Lymphocyte, Neutrophil, NLR, Platelet, PLR, WBC |
| Saloojee et al, 2018 [93] | South Africa | Schizophrenia | 67 | 48/19 | 22.8 (3.7) | 67 | 48/19 | 23.3 (2.6) | BMI, DBP, Fasting glucose, HDL, LDL, SBP, Total cholesterol, Triglycerides |
| Sayed et al, 2023 [94] | Saudi Arabia | Schizophrenia | 150 | 103/47 | 31.86 (9.06) | 120 | 85/35 | 33.79 (8.77) | BMI, Fasting glucose, HbA1c, HDL, LDL, SBP, Total cholesterol, Triglycerides |
| Sengupta et al, 2008 [95] | Canada | Schizophrenia | 38 | 33/5 | 25.4 (5.6) | 36 | 28/8 | 25.1 (5.3) | BMI, Fasting glucose, HbA1c, HDL, Insulin, Insulin resistance, LDL, Pulse pressure, Total cholesterol, Triglycerides |

| | | | | | | | | | |
|---|---|---|---|---|---|---|---|---|---|
| Shafiee-Kandjani et al, 2024 [96] | Iran | Schizophrenia | 40 | NR | 29.78 (9.60) | 40 | NR | 30.33 (8.58) | IL-6, IL-12 |
| Shinba, 2017 [97] | Japan | Depression | 14 | 7/7 | 38.5 (10) | 41 | 18/23 | 39.4 (11.9) | HRV, RHR |
| | | Anxiety | 11 | 5/6 | 35.3 (13.5) | | | | |
| Singh et al, 2022 [98] | Germany | Depression | 82 | 39/43 | 33.5 (25.0;46.3) | 129 | 52/77 | 38.0 (28.0;47.5) | BMI, CRP, Lymphocyte, MLR, Monocyte, Neutrophil, NLR |
| Song et al, 2014a; 2014b [99,100] Li et al, 2022b [101] Wang et al, 2023 [102] | China | Schizophrenia | 62 | 33/29 | 24.7 (5.5) | 60 | 33/27 | 26.2 (4.9) | Bilirubin, BMI, Fasting glucose, HDL, IL-1β, IL-6, LDL, TNF-α, Total cholesterol, Triglycerides |
| Sugimoto et al, 2018 [103] | Japan | Depression | 35 | 17/18 | 46.3 (14.3) | 35 | 13/22 | 44.0 (11.4) | IFN-γ, IL-1β, IL-6, TNF-α |
| Tai et al, 2020 [104] | Taiwan | Schizophrenia | 42 | 19/23 | 28.7 (7.2) | 42 | 19/23 | 28.8 (7.2) | DBP, HRV, RHR, SBP |
| Tang et al, 2018 [105] | China | Anxiety | 48 | 20/53 | 40.75 (12.21) | 48 | 22/26 | 39.56 (10.06) | CRP, IFN-γ, IL-1α, IL-2, IL-5, IL-6, IL-8, IL-12 |
| Tang et al, 2021 [106] | China | Depression | 139 | 66/73 | 31.08 (8.76) | 76 | 38/38 | 32.87 (9.64) | IL-6, IL-10, TNF-α |
| Tao et al, 2020 [107] | China | Schizophrenia | 90 | 44/46 | 21.5 (7.7) | 70 | 32/38 | 23.4 (5.4) | Fasting glucose, Insulin, Insulin resistance |
| Targitay Ozturk et al, 2023 [108] | Turkey | Depression | 58 | 12/46 | 43.62 (10.27) | 58 | 12/46 | 44.14 (10.29) | BMI, SBP, DBP |
| Van Nimwegen et al, 2008 [109] | Netherlands | Schizophrenia | 7 | 7/0 | 23.8 (2.2) | 7 | 7/0 | 23.0 (1.7) | Fasting glucose, Insulin, Insulin resistance |

| | | | | | | | | | |
|---|---|---|---|---|---|---|---|---|---|
| Venkatasubramanian et al, 2007 [110] | India | Schizophrenia | 44 | 23/21 | 33.0 (7.7) | 44 | 23/21 | 32.5 (7.6) | Fasting glucose, Insulin, Insulin resistance, Total cholesterol, Triglycerides |
| Verma et al, 2009 [111] | Singapore | Schizophrenia | 160 | 87/73 | 30 (6.5) | 200 | 100/100 | 30.2 (5.5) | BMI, HDL, LDL, Total cholesterol |
| Wang et al, 2014 [112] | China | Schizophrenia | 81 | 43/38 | 35.9 (11.1) | 90 | 47/43 | 34.2 (10.6) | BMD |
| Wani et al, 2015 [113] | India | Schizophrenia | 50 | 32/18 | 25.4 (4.85) | 50 | 35/15 | 26.6 (2.42) | Fasting glucose, 2-hour fasting glucose |
| Wu et al, 2013 [114] | China | Schizophrenia | 70 | 37/33 | 24.49 (6.98) | 44 | 20/24 | 26.20 (4.20) | BMI, Fasting glucose, HDL, Insulin, Insulin resistance, LDL, Total cholesterol, Triglycerides, WHR |
| Xiong et al, 2011 [115] | China | Schizophrenia | 30 | 17/13 | 22.83 (4.07) | 28 | 15/13 | 23.11 (3.21) | BMI |
| | | Depression | 30 | 13/17 | 25.63 (6.41) | | | | |
| Xiu et al, 2016 [116] | China | Schizophrenia | 256 | 150/106 | 26.4 (9.1) | 540 | 300/240 | 27.9 (11.8) | BMI |
| Xiu et al, 2018 [117] | China | Schizophrenia | 45 | 23/22 | 24.1 (6.3) | 40 | 16/24 | 45.1 (12.8) | BMI, IL-3 |
| Xiu et al, 2023 [118] | China | Schizophrenia | 43 | 19/24 | 28.3 (10.1) | 29 | 13/16 | 27.7 (8.0) | BMI, Fasting glucose, HDL, Insulin, Insulin resistance, LDL, Total cholesterol, Triglycerides |
| Yan et al, 2023 [119] | China | Schizophrenia | 40 | 28/12 | 29.5 (23, 46.5) | 36 | 22/14 | 26.5 (24, 31) | BMI, IL-1β, IL-4, IL-6, IL-10, TNF-α |

| | | | | | *Median (IQR)* | | | *Median (IQR)* | |
|---|---|---|---|---|---|---|---|---|---|
| Yang et al, 2016 [120] | China | Schizophrenia | 55 | 32/23 | 25.7 (7.8) | 43 | 20/23 | 46.5 (13.6) | IL-3 |
| Yang et al, 2018 [121] | China | Schizophrenia | 79 | 0/79 | 34.40 (8.88) | 35 | 0/35 | 37.03 (8.09) | BMI |
| Yang et al, 2021 [122] | Taiwan | Depression | 34 | 9/25 | 43.7 (11.4) | 34 | 9/25 | 43.3 (11.1) | BMI, IL-1α, IL-1β |
| Yazici et al, 2003 [123] | Turkey | Depression | 25 | 0/25 | 30.8 (8.4) | 15 | 0/15 | 31.2 (7.9) | BMD |
| Yesilkaya and Bisgin, 2024 [124] | Turkey | Bipolar | 56 | 26/30 | 32.37 (11.40) | 57 | 27/30 | 32.97 (8.83) | Albumin, CRP, Lymphocyte, Monocyte, Neutrophil, Platelet, WBC |
| Yesilkaya et al, 2024 [125] | Turkey | Schizophrenia | 69 | 43/26 | 31.02 (9.5) | 127 | 69/58 | 30.2 (8.5) | Albumin, CRP, Lymphocyte, MLR, Monocyte, Neutrophil, NLR, Platelet, PLR, WBC |
| Yu et al, 2020 [126] | China | Schizophrenia | 106 | 56/50 | 23.74 (1.01) | 120 | 54/66 | 22.63 (0.81) | MLR, NLR, Platelet, PLR |
| | | Depression | 82 | 36/46 | 21.06 (1.23) | | | | |
| Yuan et al, 2018 [127] | China | Schizophrenia | 41 | 23/18 | 23.1 (8.0) | 41 | 20/21 | 24.7 (6.7) | CRP, Fasting glucose, HDL, LDL, Triglycerides |
| Zhang et al, 2015; 2020 [128,129] | China | Schizophrenia | 120 | 61/59 | 26.5 (6.3) | 31 | 14/17 | 26.9 (5.2) | Fasting glucose, HDL, Insulin, Insulin resistance, LDL, Total cholesterol, Triglycerides |

| | | | | | | | | | |
|---|---|---|---|---|---|---|---|---|---|
| Zhang et al, 2016a [130] | China | Schizophrenia | 31 | 15/16 | 25.6 (5.1) | 71 | 32/39 | 30.2 (5.0) | BMI, Fasting glucose, HDL, Insulin, LDL, Total cholesterol, Triglycerides |
| Zhang et al, 2016b [131] | China | Depression | 50 | 17/33 | 29.2 (6.0) | 50 | 21/29 | 30.8 (6.1) | BMI |
| Zhao et al, 2022 [132] | China | Depression | 24 | 7/17 | 29.96 (8.554) | 26 | 8/18 | 31.31 (9.707) | IL-1β, IL-6, TNF-α |
| Zhou et al, 2021 [133] | China | Schizophrenia | 257 | 123/134 | M: 28.81 (8.41) F: 30.43 (9.48) | 118 | 63/55 | M: 28.70 (8.38) F: 30.60 (9.76) | BMI, DBP, Fasting glucose, HbA1c, HDL, Insulin, Insulin resistance, LDL, SBP, Total cholesterol, Triglycerides |
| Zhu et al, 2018 [134] | China | Schizophrenia | 69 | 46/23 | 25.8 (5.9) | 61 | 31/30 | 29.5 (6.7) | IL-1β, TNF-α |
| Zhu et al, 2019 [135] | China | Schizophrenia | 58 | 25/33 | 38.81 (11.64) | 31 | 12/19 | 36.77 (11.56) | CRP |
| Zhu et al, 2020; 2021 [136,137] | China | Schizophrenia | 119 | 76/43 | 29.07 (7.71) | 135 | 80/55 | 29.38 (7.21) | BMI, TNF-α |
| Zou et al, 2018 [138] | China | Depression | 75 | 24/51 | 39.2 (14.14) | 102 | 32/70 | 37.05 (11.30) | BMI, IL-1β, IL-6, IL-8, IL-10, TNF-α, TGF-b1 |

***Abbreviations:*** *BMI, body mass index; BMD, bone mineral density; CRP, C-reactive protein; DBP, diastolic blood pressure; HbA1c, glycated haemoglobin; HDL, high-density lipoprotein; HRV, heart rate variability; IFN, interferon; IL, interleukin; LDL, low-density lipoprotein; MLR, monocyte-lymphocyte ratio; NLR, neutrophil-lymphocyte ratio; NR, not reported; PLR, platelet-lymphocyte ratio; RHR, resting heart rate; SBP, systolic blood pressure; TGF, transforming growth factor; TNF, tumor necrosis factor; WBC, white blood cell; WHR, waist-hip circumference ratio*

**Supplementary Table 4. Overlap in study cohorts.**

| Study | Country | Cohort | Recruitment Period | Diagnosis | Outcomes |
|---|---|---|---|---|---|
| Almis and Egilmez, 2021 [1]<br>Kapici et al, 2023 [2] | Turkey | Adiyaman University Psychiatry Clinic | Aug 1, 2015 - Aug 1, 2020 | Schizophrenia | CRP, Fasting glucose, HDL, LDL, Lymphocyte, Monocyte, Neutrophil, NLR, Platelet, PLR, RHR, Total cholesterol, Triglycerides, WBC |
| | | | | Bipolar | Platelet |
| Borovcanin et al, 2012; 2013; 2015 [6-8] | Serbia | Clinical Centre Kragujevac and Special Hospital for Psychiatric Diseases "Dr Laza Lazarevic" | May 2010 - Mar 2011 | Schizophrenia | IFN-γ, IL-4, IL-6, IL-17, IL-23, IL-27, TGF-b |
| Fernandez-Egea et al, 2009a; 2009b [25,26]<br>Garcia-Rizo et al, 2016 [27]<br>Kirkpatrick et al, 2009 [28]<br>Kirkpatrick et al, 2010 [29] | Italy/Spain | Hospital Clinic of Barcelona | NR | Schizophrenia | CRP, DBP, Fasting glucose, 2-hour fasting glucose, HbA1c, HDL, IL-6, Insulin, Insulin resistance, LDL, Pulse pressure, RHR, SBP, Total cholesterol, Triglycerides<br>Fasting glucose, 2-hour fasting glucose, Insulin |
| | | | | Depression<br>Bipolar | Fasting glucose, 2-hour fasting glucose, Insulin |
| Haring et al, 2015 [35]<br>Balotsev et al, 2019 [36]<br>Parksepp et al, 2022 [37]<br>Korhonen et al, 2023 [38]<br>Kuuskmae et al, 2023 [39] | Estonia | Tartu University Hospital | NR | Schizophrenia | BMI, IFN-γ, IL-1α, IL-1β, IL-2, IL-4, IL-6, IL-8, IL-10, Insulin, TNF-α |

| | | | | | |
|---|---|---|---|---|---|
| Jordan et al, 2018 [43]<br>Kapogiannis et al, 2019 [44]<br>Savukoski et al, 2024 [45]<br>Steiner et al, 2020 [46] | Germany | Otto-vonGuericke-University Magdeburg, Germany (University of Magdeburg) | Feb 2008 - Jun 2018 | Schizophrenia | BMI, CRP, Fasting glucose, Insulin, Insulin resistance, Lymphocyte, MLR, Monocyte, Neutrophil, NLR |
| Kakeda et al, 2018; 2020 [47,48] | Japan | NR | NR | Depression | IFN-γ, IL-1β, IL-6, TNF-α |
| Lan et al, 2021; 2022 [56,57] | China | Affiliated Brain Hospital of Guangzhou Medical University | Dec 2016 - Dec 2019 | Depression | BMI, IFN-γ, IL-1β, IL-2, IL-4, IL-5, IL-6, IL-7, IL-8, IL-10, IL-13, IL-17, IL-23, TNF-α |
| Lang et al, 2021 [58]<br>Li et al, 2021 [59] | China | First Hospital of Shanxi Medical University and Beijing Huilongguan Hospital | NR | Schizophrenia | BMI, DBP, Fasting glucose, HbA1c, HDL, Insulin, Insulin resistance, LDL, SBP, Total cholesterol, Triglycerides |
| Li et al, 2024b [64]<br>Su et al, 2023 [65]<br>Sun et al, 2024 [66]<br>Wang et al, 2025 [67] | China | Tianjin Anding Hospital | NR | Schizophrenia | BMI, IFN-γ |
| | | | | Depression | BMI, IL-6 |
| Petrikis et al, 2015a; 2015b; 2016 [82-84] | Greece | Early Intervention in Psychosis Unit, University Hospital of Ioannina | 2012 - 2014 | Schizophrenia | Fasting glucose, HbA1c, HDL, IL-2, IL-6, IL-10, IL-17, Insulin, Insulin resistance, TGF-b2, Total cholesterol, Triglycerides |
| Saddichha et al, 2008a; 2008b; 2008c [89-91] | India | Central Institute of Psychiatry, Ranchi | Jun 2006 | Schizophrenia | BMI, DBP, Fasting glucose, 2-hour fasting glucose, HDL, SBP, Triglycerides |

| | | | | | |
|---|---|---|---|---|---|
| Song et al, 2014a; 2014b [99,100]<br>Li et al, 2022b [101]<br>Wang et al, 2023 [102] | China | First Affiliated Hospital of Zhengzhou University | NR | Schizophrenia | Bilirubin, BMI, Fasting glucose, HDL, IL-1β, IL-6, LDL, TNF-α, Total cholesterol, Triglycerides |
| Zhang et al, 2015; 2020 [128,129] | China | Beijing Huilongguan Hospital | NR | Schizophrenia | Fasting glucose, HDL, Insulin, Insulin resistance, LDL, Total cholesterol, Triglycerides |
| Zhu et al, 2020; 2021 [136,137] | China | First Hospital of Shanxi Medical University and Shanghai Mental Health Center | NR | Schizophrenia | BMI, TNF-α |

***Abbreviations:*** *BMI, body mass index; CRP, C-reactive protein; DBP, diastolic blood pressure; HbA1c, glycated haemoglobin; HDL, high-density lipoprotein; IFN, interferon; IL, interleukin; LDL, low-density lipoprotein; MLR, monocyte-lymphocyte ratio; NLR, neutrophil-lymphocyte ratio; NR, not reported; PLR, platelet-lymphocyte ratio; RHR, resting heart rate; SBP, systolic blood pressure; TGF, transforming growth factor; TNF, tumor necrosis factor; WBC, white blood cell*

**Supplementary Table 5. Excluded studies at full text screening.**

| Reference | Reason for exclusion |
|---|---|
| Aas et al, 2010 [139] | Wrong study type |
| Adamowicz et al, 2022 [140] | Wrong study type |
| Afshari et al, 2016 [141] | Wrong population |
| Afzaljavan and Talaei, 2021 [142] | No primary outcomes |
| Akcan et al, 2018 [143] | Wrong population |
| Al-Dujaili et al, 2019 [144] | Wrong population |
| Al-Hakeim et al, 2020 [145] | Wrong population |
| Almulla et al, 2023 [146] | Wrong study type |
| Almulla et al, 2024 [147] | No primary outcomes |
| Andres-Olivera et al, 2024 [148] | Wrong study type |
| Anjum et al, 2018 [149] | Wrong population |
| Anjum et al, 2019 [150] | No control group |
| Arabska et al, 2018 [151] | Wrong population |
| Armio et al, 2024a [152] | Wrong population |
| Armio et al, 2024b [153] | Wrong population |
| Arranz et al, 2004 [154] | Wrong population |
| Arzaghi et al, 2016 [155] | Wrong study type |
| Ascoli et al, 2019 [156] | Wrong population |
| Ayhan et al, 2017 [157] | Wrong population |
| Babushkina et al, 2012 [158] | Wrong population |
| Badamasi et al, 2022 [159] | No primary outcomes |
| Bahn, 2012 [160] | Wrong study type |
| Bahn, 2014 [161] | Wrong study type |
| Bai, 2014 [162] | Wrong population |
| Balaji et al, 2020 [163] | Wrong population |
| Balcioglu et al, 2021 [164] | Wrong study type |
| Balcioglu et al, 2023 [165] | Wrong population |
| Barcones et al, 2018 [166] | Wrong population |
| Baskak et al, 2008 [167] | Wrong population |
| Basoglu et al, 2010 [168] | Wrong population |
| Becklen et al, 2021 [169] | Wrong population |
| Bioque et al, 2022 [170] | Wrong population |
| Bioque et al, 2024 [171] | Wrong population |
| Blok-Husum et al, 2023 [172] | Wrong population |
| Bocchio-Chiavetto et al, 2018 [173] | Wrong population |
| Bond et al, 2009 [174] | Wrong study type |
| Bond et al, 2011a [175] | Wrong population |
| Bond et al, 2011b [176] | Wrong study type |
| Bond et al, 2014 [177] | Wrong population |
| Bond et al, 2015 [178] | Wrong study type |
| Bond et al, 2016a [179] | Wrong population |

| | |
|---|---|
| Bond et al, 2016b [180] | Wrong population |
| Bond et al, 2017a [181] | Wrong population |
| Bond et al, 2017b [182] | Wrong population |
| Bond et al, 2017c [183] | Wrong study type |
| Bond et al, 2020 [184] | Wrong population |
| Borovcanin et al, 2012 [185] | Wrong study type |
| Borovcanin et al, 2013 [186] | Wrong study type |
| Borovcanin et al, 2021 [187] | No primary outcomes |
| Boukouaci et al, 2018 [188] | Wrong population |
| Brinholi et al, 2015 [189] | Wrong population |
| Cacciotti-Saija et al, 2018 [190] | Wrong population |
| Cadenhead et al, 2015 [191] | Wrong study type |
| Cao et al, 2024 [192] | Wrong population |
| Cazzullo et al, 2001 [193] | Wrong population |
| Chakrabarty et al, 2019 [194] | Wrong population |
| Chang et al, 2013 [195] | Wrong population |
| Chang et al, 2019 [196] | Wrong study type |
| Chen et al, 2013 [197] | Wrong population |
| Chen et al, 2016 [198] | Wrong population |
| Chen et al, 2020 [199] | Wrong population |
| Chen et al, 2022 [200] | Wrong population |
| Chen et al, 2024 [201] | Wrong population |
| Cheng et al, 2023 [202] | Wrong population |
| Chenx and Tsai, 2020 [203] | Wrong population |
| Chiappelli et al, 2017 [204] | Wrong population |
| Chouinard et al, 2018 [205] | Wrong study type |
| Chouinard et al, 2019 [206] | Wrong population |
| Ciftci et al, 2024 [207] | Wrong population |
| Coello et al, 2019 [208] | Wrong population |
| Corsi-Zuelli et al, 2018 [209] | Wrong study type |
| Corsi-Zuelli et al, 2020a [210] | Wrong population |
| Corsi-Zuelli et al, 2020b [211] | Wrong study type |
| Corsi-Zuelli et al, 2022 [212] | Wrong population |
| Corsi-Zuelli et al, 2024 [213] | Wrong population |
| Costa et al, 2021 [214] | Wrong study type |
| Costa et al, 2023 [215] | No primary outcomes |
| Costa-Dookhan et al, 2020 [216] | Wrong study type |
| Coughlin et al, 2016 [217] | Wrong population |
| Crespo-Facorro et al, 2008 [218] | Wrong population |
| Cseh et al, 2013 [219] | Wrong study type |
| Cubala and Landowski, 2014 [220] | No primary outcomes |
| da Silva, 2012 [221] | Wrong study type |
| Dahl et al, 2023 [222] | No control group |
| Daria et al, 2020 [223] | Wrong population |

| | |
|---|---|
| De La Fuente-Sandoval, 2022 [224] | No primary outcomes |
| De Menezes Galvao et al, 2021 [225] | No primary outcomes |
| De Witte et al, 2014 [226] | No primary outcomes |
| De Mello et al, 2012 [227] | Wrong study type |
| Demir and Yildirim, 2022 [228] | No primary outcomes |
| Di Nicola et al, 2013 [229] | Wrong population |
| Diniz et al, 2010 [230] | Wrong population |
| Doolin and O'Keane, 2016 [231] | Wrong study type |
| Doolin et al, 2018 [232] | Wrong population |
| Du Plessis et al, 2023 [233] | Wrong population |
| Dudzinska et al, 2022 [234] | Wrong population |
| Dzikowski et al, 2020 [235] | Wrong population |
| Elghonemy, 2012 [236] | Wrong population |
| Eller et al, 2009 [237] | Wrong population |
| Ellouze et al, 2019 [238] | Wrong study type |
| Emekdar et al, 2023 [239] | Wrong population |
| Emsley et al, 2015a [240] | Wrong population |
| Emsley et al, 2015b [241] | Wrong study type |
| Emsley et al, 2023 [242] | Wrong population |
| Engberg, 2012 [243] | Wrong study type |
| Enrico et al, 2023 [244] | Wrong population |
| Eren et al, 2023 [245] | Wrong population |
| Ergun et al, 2018 [246] | Wrong population |
| Erhardt et al, 2017 [247] | Wrong study type |
| Erhardt et al, 2022 [248] | Wrong study type |
| Faisal et al, 2019 [249] | Wrong population |
| Fan et al, 2013 [250] | No primary outcomes |
| Fawzi et al, 2011 [251] | Wrong population |
| Foley et al, 2023 [252] | Wrong study type |
| Foteli et al, 2018 [253] | Wrong study type |
| Friebel et al, 2009 [254] | Wrong study type |
| Galiano Rus et al, 2022 [255] | Wrong population |
| Gallart-Palau et al, 2023 [256] | Wrong population |
| Ganguli et al, 1995 [257] | Wrong population |
| Gao et al, 2023 [258] | Wrong population |
| Garcia et al, 2016 [259] | Wrong study type |
| Garcia-Rizo et al, 2011 [260] | Wrong study type |
| Garcia-Rizo et al, 2013a [261] | No control group |
| Garcia-Rizo et al, 2013b [262] | Wrong study type |
| Garcia-Rizo et al, 2017 [263] | No primary outcomes |
| Garcia-Rizo et al, 2022 [264] | Wrong study type |
| Garcia-Rizo et al, 2024 [265] | No primary outcomes |
| Garrido-Torres et al, 2022 [266] | Wrong population |
| Garrido-Torres et al, 2023 [267] | Wrong population |

| | |
|---|---|
| Gazal et al, 2015 [268] | Wrong population |
| Gercek et al, 2023 [269] | Wrong population |
| Ghosh et al, 2022 [270] | Wrong study type |
| Gine-Serven et al, 2024 [271] | Wrong population |
| Giordano et al, 2015 [272] | Wrong study type |
| Giordano et al, 2018 [273] | Wrong study type |
| Gjerde et al, 2020 [274] | Wrong population |
| Goldstein, 1965 [275] | Wrong population |
| Gonzalez-Pinto et al, 2016 [276] | Wrong study type |
| Grande et al, 2013 [277] | Wrong study type |
| Guan et al, 2024 [278] | Wrong population |
| Guclu et al, 2024 [279] | Wrong study type |
| Guha et al, 2014 [280] | Wrong population |
| Hajek et al, 2018 [281] | Wrong study type |
| Hamilton et al, 2011 [282] | Wrong study type |
| Hatzimanolis et al, 2024 [283] | Wrong population |
| Hayes et al, 2014 [284] | Wrong population |
| He et al, 2020 [285] | Wrong population |
| Heald et al, 2023 [286] | No control group |
| Heald et al, 2024 [287] | Wrong population |
| Hepgul et al, 2012 [288] | Wrong population |
| Herberth et al, 2011 [289] | No primary outcomes |
| Hoang et al, 2022 [290] | Wrong population |
| Hou et al, 2019 [291] | No control group |
| Hu et al, 2017 [292] | Wrong population |
| Huang et al, 2022 [293] | Wrong population |
| Hughes et al, 2021 [294] | Wrong population |
| Hughes et al, 2022 [295] | Wrong population |
| Inoue et al, 2022 [296] | Wrong population |
| Ipcioglu et al, 2008 [297] | Wrong population |
| Jansen et al, 2016 [298] | Wrong study type |
| Jindal et al, 2009 [299] | Wrong population |
| Jing et al, 2024 [300] | No primary outcomes |
| Jmal et al, 2019 [301] | Wrong study type |
| Jorgensen et al, 2017 [302] | Wrong population |
| Juchnowicz et al, 2023 [303] | Wrong population |
| Juncal-Ruiz et al, 2018 [304] | Wrong population |
| Juncal-Ruiz et al, 2020 [305] | Wrong population |
| Kalmady et al, 2011 [306] | Wrong study type |
| Kalmady et al, 2014 [307] | Wrong population |
| Kampman et al, 2005 [308] | Wrong population |
| Kango et al, 2023 [309] | Wrong population |
| Karabulut et al, 2019 [310] | Wrong population |
| Karanikas et al, 2017 [311] | Wrong study type |

| | |
|---|---|
| Karpihski et al, 2017 [312] | Wrong study type |
| Kauer-Sant'Anna, 2011 [313] | Wrong population |
| Kavoor et al, 2017 [314] | Wrong population |
| Kavzoglu and Hariri, 2013 [315] | Wrong population |
| Keinanen et al, 2015 [316] | Wrong population |
| Keinanen et al, 2018 [317] | Wrong population |
| Kelsven et al, 2017 [318] | Wrong study type |
| Kelsven et al, 2020 [319] | Wrong population |
| Kendirlioglu et al, 2023 [320] | Wrong population |
| Kesebir and Turan, 2014 [321] | Wrong population |
| Kesebir et al, 2014 [322] | Wrong study type |
| Ketharanathan et al, 2020 [323] | Wrong study type |
| Kikuchi et al, 2009 [324] | Wrong population |
| Kirkpatrick et al, 2011 [325] | Wrong study type |
| Kirkpatrick et al, 2012 [326] | No primary outcomes |
| Kobayashi et al, 2017 [327] | Wrong population |
| Kondaveeti, 2022 [328] | Wrong study type |
| Kopczynska et al, 2019 [329] | Wrong population |
| Korkmaz et al, 2023 [330] | Wrong population |
| Kriisa et al, 2017 [331] | No primary outcomes |
| Kubistova et al, 2012 [332] | Wrong population |
| Kuswanto et al, 2014 [333] | Wrong population |
| Lakatos et al, 2014 [334] | Wrong study type |
| Lamichhane et al, 2021 [335] | Wrong population |
| Lang et al, 2021 [336] | Wrong population |
| Langbein et al, 2018 [337] | Wrong study type |
| Laskaris et al, 2021 [338] | Wrong population |
| Lavebratt et al, 2021 [339] | Wrong study type |
| Lee et al, 2022 [340] | Wrong study type |
| Leon-Ortiz et al, 2023 [341] | Wrong population |
| Lesh et al, 2018 [342] | Wrong population |
| Leung et al, 2023 [343] | Wrong population |
| Li et al, 2020 [344] | Wrong population |
| Li et al, 2021 [345] | Wrong population |
| Li et al, 2022a [346] | Wrong population |
| Li et al, 2022b [347] | Wrong population |
| Li et al, 2024 [348] | Wrong population |
| Liang et al, 2024 [349] | Wrong population |
| Lin et al, 2018 [350] | Wrong population |
| Linnaranta et al, 2021 [351] | Wrong population |
| Lis et al, 2020 [352] | Wrong population |
| Liu et al, 2019 [353] | Wrong population |
| Liu et al, 2020a [354] | No primary outcomes |
| Liu et al, 2020b [355] | Wrong population |

| | |
|---|---|
| Liu et al, 2022 [356] | Wrong population |
| Liu et al, 2024a [357] | No primary outcomes |
| Liu et al, 2024b [358] | Wrong population |
| Liu et al, 2024c [359] | Wrong population |
| Liu et al, 2024d [360] | Wrong population |
| Lizano et al, 2015 [361] | Wrong study type |
| Luckhoff et al, 2020 [362] | Wrong population |
| Luckhoff et al, 2022 [363] | Wrong population |
| Luckhoff et al, 2024 [364] | Wrong population |
| Luty et al, 2002 [365] | Wrong population |
| Ma et al, 2021 [366] | Wrong population |
| Maes et al, 2023a [367] | Wrong study type |
| Maes et al, 2023b [368] | Wrong study type |
| Maes et al, 2024a [369] | Wrong population |
| Maes et al, 2024b [370] | Wrong population |
| Maes et al, 2024c [371] | Wrong study type |
| Magalhaes et al, 2012 [372] | Wrong study type |
| Magdaleno Herrero et al, 2023 [373] | Wrong study type |
| Maj et al, 2020 [374] | Wrong population |
| Mantere et al, 2019 [375] | Wrong study type |
| Manzanares et al, 2014 [376] | Wrong population |
| Marcinko et al, 2007 [377] | Wrong population |
| Markovic et al, 2024 [378] | Wrong study type |
| Martins-De-Souza et al, 2010 [379] | Wrong population |
| Martorell et al, 2019 [380] | Wrong population |
| Masopust et al, 2012 [381] | Wrong study type |
| Masserini et al, 1990 [382] | Wrong population |
| Maurus et al, 2023 [383] | Wrong population |
| Mazza et al, 2017 [384] | Wrong study type |
| McWhinney et al, 2021 [385] | Wrong population |
| Mersich et al, 2016 [386] | Wrong study type |
| Michalczyk et al, 2022 [387] | Wrong population |
| Michalczyk et al, 2023a [388] | Wrong population |
| Michalczyk et al, 2023b [389] | Wrong study type |
| Milleit et al, 2009 [390] | Wrong study type |
| Miller et al, 2013 [391] | Wrong study type |
| Millett et al, 2020 [392] | Wrong population |
| Misiak et al, 2014 [393] | Wrong population |
| Misiak et al, 2015 [394] | Wrong study type |
| Mondelli et al, 2010 [395] | Wrong study type |
| Mondelli et al, 2011a [396] | Wrong population |
| Mondelli et al, 2011b [397] | Wrong study type |
| Mondelli, 2014 [398] | Wrong study type |
| Mondelli et al, 2015 [399] | Wrong population |

| | |
|---|---|
| Moody and Miller, 2018 [400] | Wrong population |
| Moreno et al, 2023 [401] | Wrong population |
| Moussiopoulou et al, 2023 [402] | Wrong population |
| Nadalin et al, 2023 [403] | No control group |
| Nadia et al, 2012 [404] | Wrong population |
| Naifar et al, 2023 [405] | Wrong study type |
| Nettis et al, 2019 [406] | Wrong population |
| Nishuty et al, 2019 [407] | Wrong population |
| North et al, 2021 [408] | Wrong population |
| Noto et al, 2015 [409] | Wrong population |
| Noto et al, 2016 [410] | Wrong population |
| Noto et al, 2019 [411] | Wrong population |
| Nunez et al, 2019 [412] | Wrong population |
| Nyboe et al, 2015 [413] | Wrong population |
| O'Donoghue et al, 2022 [414] | Wrong population |
| Omar et al, 2018 [415] | Wrong study type |
| Opgen-Rhein et al, 2009 [416] | Wrong study type |
| Orhan et al, 2018a [417] | Wrong population |
| Orhan et al, 2018b [418] | Wrong study type |
| Orhan, 2022 [419] | Wrong study type |
| Oriolo et al, 2017 [420] | Wrong study type |
| Ouyang et al, 2022 [421] | Wrong population |
| Oxenkrug et al, 2019a [422] | Wrong study type |
| Oxenkrug et al, 2019b [423] | Wrong study type |
| Ozel et al, 2022 [424] | Wrong study type |
| Pan et al, 2021 [425] | Wrong population |
| Panizzutti et al, 2015 [426] | Wrong population |
| Peng et al, 2022 [427] | No primary outcomes |
| Pesce et al, 2014 [428] | No primary outcomes |
| Petrikis et al, 2024 [429] | No primary outcomes |
| Phutane et al, 2011 [430] | Wrong population |
| Pradeep et al, 2012 [431] | Wrong population |
| Prasad et al, 2016 [432] | Wrong population |
| Pujol et al, 2022 [433] | Wrong population |
| Qian et al, 2020 [434] | Wrong population |
| Qiao et al, 2024 [435] | No primary outcomes |
| Qiu et al, 2022 [436] | Wrong population |
| Ramsey et al, 2013 [437] | Wrong study type |
| Reale et al, 2011 [438] | No primary outcomes |
| Reddy et al, 2013 [439] | Wrong study type |
| Refisch et al, 2023 [440] | Wrong population |
| Refisch et al, 2024 [441] | Wrong population |
| Rizzo et al, 2016 [442] | Wrong study type |
| Rog et al, 2022 [443] | Wrong study type |

| | |
|---|---|
| Ruljancic et al, 2024 [444] | Wrong study type |
| Saddichha et al, 2010 [445] | Wrong study type |
| Saddichha and Akhtar, 2011a [446] | Wrong study type |
| Saddichha and Akhtar, 2011b [447] | Wrong study type |
| Saleem et al, 2010 [448] | Wrong study type |
| Samoud et al, 2010a [449] | Wrong study type |
| Samoud et al, 2010b [450] | Wrong study type |
| Schwieler et al, 2017 [451] | Wrong study type |
| Seitz et al, 2019 [452] | Wrong population |
| Serpa et al, 2015 [453] | Wrong study type |
| Serpa et al, 2023 [454] | Wrong population |
| Severance et al, 2013 [455] | No primary outcomes |
| Shafiee-Kandjani et al, 2023 [456] | Wrong population |
| Shahraki et al, 2016 [457] | Wrong population |
| Shan et al, 2020 [458] | Wrong population |
| Shi et al, 2020 [459] | No primary outcomes |
| Shi et al, 2021 [460] | No primary outcomes |
| Shivakumar et al, 2020 [461] | Wrong population |
| Silva et al, 2017 [462] | Wrong study type |
| Singh et al, 2023 [463] | Wrong population |
| Skibinska et al, 2017 [464] | Wrong population |
| Skibinska et al, 2019 [465] | Wrong population |
| Skorobogatov et al, 2024 [466] | Wrong population |
| Song et al, 2009 [467] | Wrong population |
| Spelman et al, 2007 [468] | Wrong study type |
| Srihari et al, 2013 [469] | Wrong population |
| Steiner et al, 2010 [470] | Wrong study type |
| Steiner et al, 2017 [471] | Wrong study type |
| Steiner et al, 2019 [472] | Wrong population |
| Steiner et al, 2020 [473] | Wrong study type |
| Stelzhammer et al, 2014 [474] | No primary outcomes |
| Stojanovic et al, 2014 [475] | Wrong population |
| Stojanovic et al, 2024 [476] | Wrong study type |
| Su et al, 2022 [477] | Wrong population |
| Subbanna et al, 2018 [478] | Wrong population |
| Sun et al, 2016 [479] | Wrong population |
| Sun et al, 2024 [480] | Wrong population |
| Sutterland et al, 2016 [481] | Wrong study type |
| Suvisaari et al, 2018 [482] | Wrong study type |
| Suzuki et al, 2008 [483] | Wrong population |
| Szeszko et al, 2013 [484] | Wrong study type |
| Szymona et al, 2019 [485] | Wrong population |
| Taene et al, 2020 [486] | No primary outcomes |
| Tang et al, 2017 [487] | Wrong study type |

| | |
|---|---|
| Tatay-Manteiga et al, 2017 [488] | Wrong population |
| Teja et al, 2018 [489] | Wrong study type |
| Thakore et al, 2002 [490] | Wrong population |
| Theodoropoulou et al, 2001 [491] | Wrong population |
| Tian et al, 2021a [492] | Wrong population |
| Tian et al, 2021b [493] | Wrong population |
| Tofani et al, 2015 [494] | Wrong study type |
| Tosato et al, 2018 [495] | Wrong study type |
| Turan and Kesebir, 2013 [496] | Wrong study type |
| Turner et al, 2017 [497] | Wrong study type |
| Twayej et al, 2020 [498] | Wrong population |
| Udupa et al, 2007 [499] | Wrong population |
| Unknown, 2024 [500] | Wrong study type |
| Uzbekov et al, 2019 [501] | Wrong study type |
| Valkonen-Korhonen et al, 2003 [502] | Wrong population |
| Varden Gjerde et al, 2024 [503] | No primary outcomes |
| Varsak et al, 2015 [504] | Wrong study type |
| Vazquez-Bourgon et al, 2022 [505] | Wrong population |
| Veling et al, 2013 [506] | Wrong study type |
| Venkatasubramanian et al, 2010 [507] | No primary outcomes |
| Verma et al, 2010 [508] | Wrong study type |
| Verma, 2014 [509] | Wrong study type |
| Vianna-Sulzbach et al, 2014 [510] | Wrong study type |
| Vilella et al, 2022 [511] | Wrong study type |
| Vochoskova et al, 2023a [512] | No primary outcomes |
| Vochoskova et al, 2023b [513] | Wrong study type |
| Walsh et al, 2002 [514] | No primary outcomes |
| Wang et al, 2024a [515] | No primary outcomes |
| Wang et al, 2024b [516] | Wrong population |
| Wang et al, 2024c [517] | Wrong population |
| Wang et al, 2024d [518] | Wrong population |
| Wei et al, 2022a [519] | Wrong population |
| Wei et al, 2022b [520] | Wrong population |
| Whitson et al, 2021 [521] | Wrong population |
| Wu et al, 2022 [522] | Wrong population |
| Xiu et al, 2008 [523] | Wrong study type |
| Xiu et al, 2012 [524] | Wrong population |
| Xiu et al, 2014a [525] | Wrong population |
| Xiu et al, 2014b [526] | Wrong study type |
| Xu et al, 2023 [527] | Wrong population |
| Xu et al, 2024 [528] | Wrong population |
| Yan et al, 2022 [529] | Wrong population |
| Yan et al, 2024a [530] | Wrong population |
| Yan et al, 2024b [531] | Wrong population |

| | |
|---|---|
| Yao et al, 2017 [532] | Wrong study type |
| Yoshimura et al, 2010 [533] | Wrong study type |
| Zhai et al, 2021 [534] | Wrong population |
| Zhang et al, 2013 [535] | Wrong population |
| Zhang et al, 2023 [536] | Wrong population |
| Zhao et al, 2020 [537] | No primary outcomes |
| Zhao et al, 2024 [538] | Wrong population |
| Zheng et al, 2013 [539] | Wrong population |
| Zheng et al, 2023 [540] | Wrong population |
| Ziauddeen et al, 2016 [541] | Wrong population |
| Zorrilla et al, 1996 [542] | Wrong population |
| Zozulya et al, 2021 [543] | Wrong study type |

**Supplementary Table 5. JBI risk of bias critical appraisal checklist for case control studies.**

| Study | Q1 | Q2 | Q3 | Q4 | Q5 | Q6 | Q7 | Q8 | Q9 | Q10 | Overall |
|---|---|---|---|---|---|---|---|---|---|---|---|
| Almis and Egilmez, 2021 [1] | Y | Y | Y | Y | Y | Y | Y | Y | NA | Y | Low |
| Amoli et al, 2019 [3] | Y | Y | Y | Y | Y | Y | Y | Y | NA | Y | Low |
| Bai et al, 2021 [4] | Y | Y | Y | Y | Y | Y | Y | Y | NA | Y | Low |
| Balotsev et al, 2019 [36] | Y | Y | Y | Y | Y | N | N | Y | NA | Y | Low |
| Bolu et al, 2019 [5] | Y | Y | Y | Y | Y | Y | Y | Y | NA | Y | Low |
| Borovcanin et al, 2012 [6] | Y | Y | Y | Y | Y | Y | Y | Y | NA | Y | Low |
| Borovcanin et al, 2013 [7] | Y | Y | Y | Y | Y | Y | Y | Y | NA | Y | Low |
| Borovcanin et al, 2015 [8] | Y | Y | Y | Y | Y | Y | Y | Y | NA | Y | Low |
| Cabello-Rangel et al, 2023 [9] | Y | Y | Y | Y | Y | Y | Y | Y | NA | Y | Low |
| Chang et al, 2017 [10] | Y | Y | Y | Y | Y | Y | Y | Y | NA | Y | Low |
| Chen et al, 2016 [11] | Y | N | Y | Y | Y | Y | N | Y | NA | Y | Low |
| Chen et al, 2018 [12] | Y | Y | Y | Y | Y | Y | Y | Y | NA | Y | Low |
| Chen et al, 2021a [13] | Y | Y | Y | Y | Y | Y | Y | Y | NA | Y | Low |
| Chen et al, 2021b [14] | Y | Y | Y | Y | Y | Y | Y | Y | NA | Y | Low |
| Chen et al, 2022a [15] | Y | Y | Y | Y | Y | Y | Y | Y | NA | Y | Low |
| Chen et al, 2022b [16] | Y | Y | Y | Y | Y | Y | Y | Y | NA | Y | Low |
| Chen et al, 2024 [17] | Y | Y | Y | Y | Y | Y | Y | Y | NA | Y | Low |
| Cropley et al, 2023 [18] | Y | Y | Y | Y | Y | Y | Y | Y | NA | Y | Low |
| Dai et al, 2020 [19] | Y | Y | Y | Y | Y | Y | Y | Y | NA | Y | Low |
| Dasgupta et al, 2010 [20] | Y | Y | Y | Y | Y | Y | Y | Y | NA | Y | Low |
| De Berardis et al, 2013 [21] | Y | Y | Y | Y | Y | Y | N | Y | NA | Y | Low |
| de Menezes Galvao et al, 2021 [22] | Y | Y | Y | Y | Y | Y | Y | Y | NA | Y | Low |
| Ding et al, 2014 [23] | Y | Y | Y | Y | Y | Y | Y | Y | NA | Y | Low |
| Dongxia et al, 2024 [24] | Y | Y | Y | Y | Y | Y | Y | Y | NA | Y | Low |
| Fernandez-Egea et al, 2009a [25] | Y | N | Y | Y | Y | Y | Y | Y | NA | Y | Low |
| Fernandez-Egea et al, 2009b [26] | Y | Y | Y | Y | Y | Y | Y | Y | NA | Y | Low |
| Frydecka et al, 2018 [30] | Y | Y | Y | Y | Y | Y | Y | Y | NA | Y | Low |

| | | | | | | | | | | |
|---|---|---|---|---|---|---|---|---|---|---|
| Gao et al, 2024 [31] | Y | Y | Y | Y | Y | Y | Y | Y | NA | Y | Low |
| Garcia-Rizo et al, 2016 [27] | Y | Y | Y | Y | Y | Y | Y | Y | NA | Y | Low |
| Garcia-Rizo et al, 2019 [32] | Y | N | Y | Y | Y | Y | Y | Y | NA | Y | Low |
| Garrido-Torres et al, 2022 [33] | Y | Y | Y | Y | Y | Y | Y | Y | NA | Y | Low |
| Goyal et al, 2020 [34] | Y | Y | Y | Y | Y | Y | Y | Y | NA | Y | Low |
| Haring et al, 2015 [35] | Y | Y | Y | Y | Y | Y | Y | Y | NA | Y | Low |
| Herran et al, 2000 [40] | Y | Y | Y | Y | Y | Y | Y | Y | NA | Y | Low |
| Huang et al, 2022 [41] | Y | N | Y | Y | Y | Y | Y | Y | NA | Y | Low |
| Joaquim et al, 2018 [42] | Y | Y | Y | Y | Y | Y | Y | Y | NA | Y | Low |
| Jordan et al, 2018 [43] | Y | Y | Y | Y | Y | Y | Y | Y | NA | Y | Low |
| Kakeda et al, 2018 [47] | Y | Y | Y | Y | Y | Y | Y | Y | NA | Y | Low |
| Kakeda et al, 2020 [48] | Y | N | Y | Y | Y | Y | Y | Y | NA | Y | Low |
| Kalmady et al, 2018 [49] | Y | Y | Y | Y | Y | Y | Y | Y | NA | Y | Low |
| Kapici et al, 2023 [2] | Y | Y | Y | Y | Y | Y | Y | Y | NA | Y | Low |
| Kapogiannis et al, 2019 [44] | Y | Y | Y | Y | Y | Y | Y | Y | NA | Y | Low |
| Keri et al, 2014 [50] | Y | N | Y | Y | Y | Y | Y | Y | NA | Y | Low |
| Keri et al, 2017 [51] | Y | Y | Y | Y | Y | Y | Y | Y | NA | Y | Low |
| Khan, 2022 [52] | Y | Y | Y | Y | Y | Y | Y | Y | NA | Y | Low |
| Kim et al, 2021 [53] | Y | Y | Y | Y | Y | Y | Y | Y | NA | Y | Low |
| Kirkpatrick et al, 2009 [28] | Y | Y | Y | Y | Y | Y | Y | Y | NA | Y | Low |
| Kirkpatrick et al, 2010 [29] | Y | Y | Y | Y | Y | Y | Y | Y | NA | Y | Low |
| Kolenic et al, 2018 [54] | Y | Y | Y | Y | Y | Y | Y | Y | NA | Y | Low |
| Korhonen et al, 2023 [38] | Y | Y | Y | Y | Y | Y | Y | Y | NA | Y | Low |
| Kuuskmae et al, 2023 [39] | Y | Y | Y | Y | Y | Y | Y | Y | NA | Y | Low |
| Kuwano et al, 2018 [55] | Y | Y | Y | Y | Y | Y | Y | Y | NA | Y | Low |
| Lan et al, 2021 [56] | Y | Y | Y | Y | Y | Y | Y | Y | Y | Y | Low |
| Lan et al, 2022 [57] | Y | Y | Y | Y | Y | Y | Y | Y | NA | Y | Low |
| Lang et al, 2021 [58] | Y | Y | Y | Y | Y | Y | Y | Y | NA | Y | Low |
| Leo et al, 2006 [60] | Y | Y | Y | Y | Y | Y | Y | Y | NA | Y | Low |
| Li et al, 2013 [61] | Y | Y | Y | Y | Y | Y | Y | Y | NA | Y | Low |

| | | | | | | | | | | | |
|---|---|---|---|---|---|---|---|---|---|---|---|
| Li et al, 2021 [59] | Y | Y | Y | Y | Y | Y | Y | Y | NA | Y | Low |
| Li et al, 2022a [62] | Y | Y | Y | Y | Y | Y | Y | Y | NA | Y | Low |
| Li et al, 2022b [101] | Y | Y | Y | Y | Y | Y | Y | Y | NA | Y | Low |
| Li et al, 2024a [63] | Y | Y | Y | Y | Y | Y | Y | Y | NA | Y | Low |
| Li et al, 2024b [64] | Y | Y | Y | Y | Y | Y | Y | Y | Y | Y | Low |
| Li et al, 2025 [68] | Y | Y | Y | Y | Y | Y | Y | Y | Y | Y | Low |
| Liang et al, 2015 [69] | Y | Y | Y | Y | Y | Y | Y | Y | NA | Y | Low |
| Liang et al, 2019 [70] | Y | Y | Y | Y | Y | N | N | Y | NA | Y | Low |
| Liang et al, 2022 [71] | Y | Y | Y | Y | Y | Y | Y | Y | NA | Y | Low |
| Lin et al, 2021 [72] | Y | Y | Y | Y | Y | Y | N | Y | NA | Y | Low |
| Liu et al, 2015 [73] | Y | Y | Y | Y | Y | Y | N | Y | NA | Y | Low |
| Liu et al, 2022 [74] | Y | Y | Y | Y | Y | Y | N | Y | NA | Y | Low |
| Liu et al, 2023 [75] | Y | Y | Y | Y | Y | N | N | Y | NA | Y | Low |
| Milleit et al, 2019 [76] | Y | Y | Y | Y | Y | Y | Y | Y | NA | Y | Low |
| Misiak et al, 2014 [77] | Y | Y | Y | Y | Y | Y | Y | Y | NA | Y | Low |
| Misiak et al, 2016 [78] | Y | Y | Y | Y | Y | Y | Y | Y | NA | Y | Low |
| Ntouros et al, 2018 [79] | Y | N | Y | Y | Y | Y | Y | Y | NA | Y | Low |
| Onur et al, 2021 [80] | Y | Y | Y | Y | Y | Y | Y | Y | NA | Y | Low |
| Pan et al, 2022 [81] | Y | Y | Y | Y | Y | Y | Y | Y | NA | Y | Low |
| Parksepp et al, 2022 [37] | Y | Y | Y | Y | Y | Y | Y | Y | NA | Y | Low |
| Petrikis et al, 2015a [82] | Y | Y | Y | Y | Y | Y | Y | Y | NA | Y | Low |
| Petrikis et al, 2015b [83] | Y | Y | Y | Y | Y | Y | Y | Y | NA | Y | Low |
| Puangsri and Ninla-Aesong, 2021 [85] | Y | Y | Y | Y | Y | Y | Y | Y | NA | Y | Low |
| Qiao et al, 2024 [435] | Y | Y | Y | Y | Y | Y | Y | Y | NA | Y | Low |
| Radu et al, 2020 [86] | Y | Y | Y | Y | Y | Y | Y | Y | NA | Y | Low |
| Reddy et al, 2003 [87] | Y | Y | Y | Y | Y | Y | Y | Y | NA | Y | Low |
| Ryan et al, 2003 [88] | Y | Y | Y | Y | Y | Y | Y | Y | NA | Y | Low |
| Saddichha et al, 2008a [89] | Y | Y | Y | Y | Y | Y | Y | Y | NA | Y | Low |
| Saddichha et al, 2008b [90] | Y | Y | Y | Y | Y | Y | Y | Y | NA | Y | Low |
| Saddichha et al, 2008c [91] | Y | Y | Y | Y | Y | N | N | Y | NA | Y | Low |

| | | | | | | | | | | |
|---|---|---|---|---|---|---|---|---|---|---|
| Sahpolat et al, 2022 [92] | Y | N | Y | Y | Y | Y | Y | Y | NA | Y | Low |
| Saloojee et al, 2018 [93] | Y | Y | Y | Y | Y | Y | Y | Y | NA | Y | Low |
| Savukoski et al, 2024 [45] | Y | Y | Y | Y | Y | N | N | Y | NA | Y | Low |
| Sayed et al, 2023 [94] | Y | Y | Y | Y | Y | Y | Y | Y | NA | Y | Low |
| Sengupta et al, 2008 [95] | Y | Y | Y | Y | Y | Y | Y | Y | NA | Y | Low |
| Shafiee-Kandjani et al, 2024 [96] | Y | N | Y | Y | Y | Y | Y | Y | NA | Y | Low |
| Shinba, 2017 [97] | Y | Y | Y | Y | Y | Y | Y | Y | NA | Y | Low |
| Singh et al, 2022 [98] | Y | Y | Y | Y | Y | Y | N | Y | NA | Y | Low |
| Song et al, 2014a [99] | Y | N | Y | Y | Y | Y | N | Y | NA | Y | Low |
| Song et al, 2014b [100] | Y | Y | Y | Y | Y | Y | N | Y | NA | Y | Low |
| Steiner et al, 2020 [46] | Y | Y | Y | Y | Y | Y | Y | Y | NA | Y | Low |
| Su et al, 2023 [65] | Y | Y | Y | Y | Y | Y | Y | Y | NA | Y | Low |
| Sugimoto et al, 2018 [103] | Y | Y | Y | Y | Y | Y | Y | Y | NA | Y | Low |
| Sun et al, 2024 [66] | Y | Y | Y | Y | Y | Y | Y | Y | NA | Y | Low |
| Tai et al, 2020 [104] | Y | Y | Y | Y | Y | N | N | Y | NA | Y | Low |
| Tang et al, 2018 [105] | Y | Y | Y | Y | Y | Y | Y | Y | NA | Y | Low |
| Tang et al, 2021 [106] | Y | Y | Y | Y | Y | Y | Y | Y | NA | Y | Low |
| Tao et al, 2020 [107] | Y | N | Y | Y | Y | N | N | Y | NA | Y | Low |
| Targitay Ozturk et al, 2023 [108] | Y | Y | Y | Y | Y | Y | Y | Y | NA | Y | Low |
| Van Nimwegen et al, 2008 [109] | Y | Y | Y | Y | Y | Y | Y | Y | NA | Y | Low |
| Venkatasubramanian et al, 2007 [110] | Y | Y | Y | Y | Y | Y | Y | Y | NA | Y | Low |
| Verma et al, 2009 [111] | Y | Y | Y | Y | Y | Y | Y | Y | NA | Y | Low |
| Wang et al, 2014 [112] | Y | N | Y | Y | Y | Y | Y | Y | NA | Y | Low |
| Wang et al, 2023 [102] | Y | Y | Y | Y | Y | Y | Y | Y | NA | Y | Low |
| Wang et al, 2025 [67] | Y | Y | Y | Y | Y | Y | Y | Y | NA | Y | Low |
| Wani et al, 2015 [113] | Y | N | Y | Y | Y | Y | Y | Y | NA | Y | Low |
| Wu et al, 2013 [114] | Y | Y | Y | Y | Y | Y | Y | Y | NA | Y | Low |
| Xiong et al, 2011 [115] | Y | Y | Y | Y | Y | Y | Y | Y | NA | Y | Low |
| Xiu et al, 2016 [116] | Y | Y | Y | Y | Y | Y | Y | Y | NA | Y | Low |
| Xiu et al, 2018 [117] | Y | Y | Y | Y | Y | Y | Y | Y | NA | Y | Low |

| | | | | | | | | | | | |
|---|---|---|---|---|---|---|---|---|---|---|---|
| Xiu et al, 2023 [118] | Y | Y | Y | Y | Y | N | N | Y | NA | Y | Low |
| Yan et al, 2023 [119] | Y | Y | Y | Y | Y | Y | Y | Y | NA | Y | Low |
| Yang et al, 2016 [120] | Y | Y | Y | Y | Y | Y | Y | Y | NA | Y | Low |
| Yang et al, 2018 [121] | Y | Y | Y | Y | Y | Y | Y | Y | NA | Y | Low |
| Yang et al, 2021 [122] | Y | Y | Y | Y | Y | Y | Y | Y | NA | Y | Low |
| Yazici et al, 2003 [123] | Y | Y | Y | Y | Y | Y | Y | Y | NA | Y | Low |
| Yesilkaya and Bisgin, 2024 [124] | Y | Y | Y | Y | Y | Y | Y | Y | NA | Y | Low |
| Yesilkaya et al, 2024 [125] | Y | N | Y | Y | Y | Y | N | Y | NA | Y | Low |
| Yu et al, 2020 [126] | Y | Y | Y | Y | Y | Y | Y | Y | NA | Y | Low |
| Yuan et al, 2018 [127] | Y | Y | Y | Y | Y | Y | Y | Y | NA | Y | Low |
| Zhang et al, 2015 [128] | Y | Y | Y | Y | Y | Y | Y | Y | NA | Y | Low |
| Zhang et al, 2016a [130] | Y | Y | Y | Y | Y | Y | Y | Y | NA | Y | Low |
| Zhang et al, 2016b [131] | Y | Y | Y | Y | Y | Y | Y | Y | NA | Y | Low |
| Zhang et al, 2020 [129] | Y | Y | Y | Y | Y | Y | Y | Y | NA | Y | Low |
| Zhao et al, 2022 [132] | Y | Y | Y | Y | Y | Y | Y | Y | NA | Y | Low |
| Zhou et al, 2021 [133] | Y | Y | Y | Y | Y | Y | Y | Y | NA | Y | Low |
| Zhu et al, 2018 [134] | Y | Y | Y | Y | Y | Y | Y | Y | NA | Y | Low |
| Zhu et al, 2019 [135] | Y | Y | Y | Y | Y | Y | N | Y | NA | Y | Low |
| Zhu et al, 2020 [136] | Y | Y | Y | Y | Y | Y | Y | Y | NA | Y | Low |
| Zhu et al, 2021 [137] | Y | Y | Y | Y | Y | Y | Y | Y | NA | Y | Low |
| Zou et al, 2018 [138] | Y | Y | Y | Y | Y | Y | Y | Y | NA | Y | Low |

**Supplementary Table 6. Meta-analysis raw data of pro-inflammatory biomarkers in schizophrenia.**

(a) IFN-γ

| Study | Patients (n) | Patients Mean | Patients SD | Controls (n) | Controls Mean | Controls SD |
|---|---|---|---|---|---|---|
| Borovcanin et al, 2012 [6] | 88 | 78.51 | 37.28 | 36 | 29.06 | 10.68 |
| Chen et al, 2021b [14] | 24 | 1.188 | 0.126 | 25 | 0.949 | 0.088 |
| Ding et al, 2014 [23] | 69 | 509.73 | 144.44 | 60 | 438.54 | 113.7 |
| Frydecka et al, 2018 [30] | 39 | 54.5 | 52.6 | 39 | 44.1 | 25.6 |
| Kalmady et al, 2018 [49] | 75 | 1.22 | 6.56 | 102 | 1.38 | 1.58 |
| Milleit et al, 2019 [76] | 18 | 0 | 0 | 25 | 0.2347 | 69.51 |
| Ntouros et al, 2018 [79] | 25 | 39 | 23.12 | 23 | 22 | 16.36 |
| Parksepp et al, 2022 [37] | 54 | 0.36 | 0.14 | 58 | 0.3 | 0.09 |

(b) IL-1β

| Study | Patients (n) | Patients Mean | Patients SD | Controls (n) | Controls Mean | Controls SD |
|---|---|---|---|---|---|---|
| Chen et al, 2021b [14] | 24 | 1.231 | 0.421 | 25 | 1.053 | 0.444 |
| Dai et al, 2020 [19] | 46 | 61.55 | 21.08 | 60 | 24.77 | 8.09 |
| Frydecka et al, 2018 [30] | 39 | 1.3 | 0.3 | 39 | 1.4 | 0.4 |
| Joaquim et al, 2018 [42] | 28 | 0.28 | 0.28 | 30 | 0.39 | 0.58 |
| Ntouros et al, 2018 [79] | 25 | 51 | 39.117 | 23 | 68 | 44.82 |
| Parksepp et al, 2022 [37] | 54 | 1.41 | 0.66 | 58 | 1.63 | 0.91 |
| Song et al, 2014a [99] | 62 | 53.28 | 12.62 | 60 | 23.49 | 15.27 |
| Zhu et al, 2018 [134] | 69 | 1.7 | 0.2 | 61 | 8.3 | 7.5 |

(c) IL-5

| Study | Patients (n) | Patients Mean | Patients SD | Controls (n) | Controls Mean | Controls SD |
|---|---|---|---|---|---|---|
| Chen et al, 2021b [14] | 24 | 1.04 | 0.14 | 25 | 0.979 | 0 |
| Frydecka et al, 2018 [30] | 39 | 2.9 | 3.1 | 39 | 2 | 1 |
| Ntouros et al, 2018 [79] | 25 | 11.5 | 4.53 | 23 | 3.4 | 4.4 |

(d) IL-6

| Study | Patients (n) | Patients Mean | Patients SD | Controls (n) | Controls Mean | Controls SD |
|---|---|---|---|---|---|---|
| Borovcanin et al, 2012 [6] | 88 | 27.86 | 8.07 | 36 | 8.14 | 3.47 |
| Chen et al, 2021b [14] | 24 | 0.696 | 0.122 | 25 | 0.851 | 0.322 |
| Dai et al, 2020 [19] | 46 | 5.08 | 1.14 | 60 | 4.2 | 0.94 |
| Ding et al, 2014 [23] | 69 | 14.75 | 4.52 | 60 | 11.76 | 5.05 |
| Fernandez-Egea et al, 2009a [25] | 50 | 3.63 | 6.91 | 50 | 1.02 | 2.1 |
| Frydecka et al, 2018 [30] | 39 | 9.8 | 4.1 | 39 | 9 | 2.7 |
| Kalmady et al, 2018 [49] | 75 | 3.47 | 3.07 | 102 | 2.33 | 1.33 |
| Milleit et al, 2019 [76] | 18 | 1.904 | 5.323 | 25 | 0.6606 | 1.522 |
| Parksepp et al, 2022 [37] | 54 | 1.27 | 1.14 | 58 | 0.76 | 0.59 |
| Shafiee-Kandjani et al, 2024 [96] | 40 | 42.75 | 24.64 | 40 | 7.01 | 9.32 |
| Song et al, 2014a [99] | 62 | 33.98 | 14.13 | 60 | 15.53 | 7.16 |

(e) IL-8

| Study | Patients (n) | Patients Mean | Patients SD | Controls (n) | Controls Mean | Controls SD |
|---|---|---|---|---|---|---|
| Chen et al, 2021b [14] | 24 | 1.744 | 0.312 | 25 | 1.823 | 0.317 |
| Frydecka et al, 2018 [30] | 39 | 41 | 23.7 | 39 | 46.4 | 29.3 |
| Milleit et al, 2019 [76] | 18 | 1.274 | 2.186 | 25 | 0.796 | 2.022 |
| Ntouros et al, 2018 [79] | 25 | 218 | 35.69 | 23 | 381 | 125.11 |
| Parksepp et al, 2022 [37] | 54 | 6.58 | 4.07 | 58 | 5.98 | 2.9 |

(f) IL-12

| Study | Patients (n) | Patients Mean | Patients SD | Controls (n) | Controls Mean | Controls SD |
|---|---|---|---|---|---|---|
| Chen et al, 2021b [14] | 24 | 2.123 | 0.172 | 25 | 2.158 | 0.089 |
| Frydecka et al, 2018 [30] | 39 | 68.5 | 39.5 | 39 | 59.8 | 34.8 |
| Ntouros et al, 2018 [79] | 25 | 14 | 11.23 | 23 | 12 | 11.17 |
| Shafiee-Kandjani et al, 2024 [96] | 40 | 0.77 | 0.85 | 40 | 1.29 | 1.36 |

(g) IL-17

| Study | Patients (n) | Patients Mean | Patients SD | Controls (n) | Controls Mean | Controls SD |
|---|---|---|---|---|---|---|
| Borovcanin et al, 2012 [6] | 88 | 6.57 | 4.62 | 36 | 30.86 | 13.95 |
| Chen et al, 2021b [14] | 113 | 13.61 | 2.02 | 58 | 12.97 | 1.86 |
| Ding et al, 2014 [23] | 69 | 17.69 | 6.73 | 60 | 15.61 | 5.46 |
| Kalmady et al, 2018 [49] | 75 | 9.96 | 311.82 | 102 | 11.58 | 62 |

(h) TNF-α

| Study | Patients (n) | Patients Mean | Patients SD | Controls (n) | Controls Mean | Controls SD |
|---|---|---|---|---|---|---|
| Chen et al, 2021b [14] | 24 | 0.721 | 0.224 | 25 | 0.717 | 0.195 |
| Frydecka et al, 2018 [30] | 39 | 25.9 | 6.8 | 39 | 26.2 | 14.8 |
| Kalmady et al, 2018 [49] | 75 | 1 | 1.06 | 102 | 1.07 | 0.88 |
| Milleit et al, 2019 [76] | 18 | 2.319 | 11.51 | 25 | 0.8033 | 3.323 |
| Ntouros et al, 2018 [79] | 25 | 9 | 8.69 | 23 | 8 | 8.4 |
| Parksepp et al, 2022 [37] | 38 | 1.98 | 0.84 | 37 | 2.22 | 1.4 |
| Song et al, 2014a [99] | 62 | 50.08 | 12.86 | 60 | 32.12 | 15.23 |
| Zhu et al, 2018 [134] | 69 | 8.2 | 2 | 61 | 15.4 | 7 |
| Zhu et al, 2020 [136] | 119 | 2.21 | 0.33 | 135 | 2.11 | 0.36 |

**Supplementary Table 7. Meta-analysis raw data of anti-inflammatory biomarkers in schizophrenia.**

(a) IL-4

| Study | Patients (n) | Patients Mean | Patients SD | Controls (n) | Controls Mean | Controls SD |
|---|---|---|---|---|---|---|
| Borovcanin et al, 2012 [6] | 88 | 25.1 | 10.73 | 36 | 12.09 | 8 |
| Chen et al, 2021b [14] | 24 | 1.564 | 0.34 | 25 | 1.272 | 0 |
| Frydecka et al, 2018 [30] | 39 | 14.2 | 3.4 | 39 | 13.8 | 3.1 |
| Kalmady et al, 2018 [49] | 75 | 1.2 | 2.19 | 102 | 1.11 | 1.35 |
| Milleit et al, 2019 [76] | 18 | 33.49 | 64.68 | 25 | 7.925 | 22.63 |
| Ntouros et al, 2018 [79] | 25 | 71 | 69.76 | 23 | 68 | 68.7 |
| Parksepp et al, 2022 [37] | 54 | 1.87 | 0.57 | 58 | 1.33 | 0.51 |

(b) IL-10

| Study | Patients (n) | Patients Mean | Patients SD | Controls (n) | Controls Mean | Controls SD |
|---|---|---|---|---|---|---|
| Chen et al, 2021b [14] | 24 | 0.91 | 0.224 | 25 | 0.791 | 0.369 |
| Frydecka et al, 2018 [30] | 39 | 12.9 | 7.2 | 39 | 13.2 | 8.6 |
| Kalmady et al, 2018 [49] | 75 | 1.83 | 3.4 | 102 | 1.59 | 1.07 |
| Ntouros et al, 2018 [79] | 25 | 19 | 4.55 | 23 | 10 | 3.36 |
| Parksepp et al, 2022 [37] | 54 | 0.73 | 0.37 | 58 | 0.71 | 0.42 |

(c) IL-13

| Study | Patients (n) | Patients Mean | Patients SD | Controls (n) | Controls Mean | Controls SD |
|---|---|---|---|---|---|---|
| Chen et al, 2021b [14] | 24 | 1.304 | 0.294 | 25 | 1.141 | 0.303 |
| Frydecka et al, 2018 [30] | 39 | 15.7 | 8.2 | 39 | 16.2 | 11 |
| Milleit et al, 2019 [76] | 18 | 108.3 | 161.1 | 25 | 8.257 | 14.47 |

**Supplementary Table 8. Meta-analysis raw data of adaptive-inflammatory biomarkers in schizophrenia.**

IL-2

| Study | Patients (n) | Patients Mean | Patients SD | Controls (n) | Controls Mean | Controls SD |
|---|---|---|---|---|---|---|
| Chen et al, 2021b [14] | 24 | 0.911 | 0.367 | 25 | 0.581 | 0.187 |
| Frydecka et al, 2018 [30] | 39 | 14.5 | 24.3 | 39 | 13.5 | 16.5 |
| Kalmady et al, 2018 [49] | 75 | 0.5 | 2.18 | 102 | 0.69 | 1.09 |
| Ntouros et al, 2018 [79] | 25 | 99 | 96.12 | 23 | 97 | 93.1 |
| Parksepp et al, 2022 [37] | 54 | 3.15 | 0.87 | 58 | 2.27 | 1.26 |

**Supplementary Table 9. Meta-analysis raw data of other inflammatory biomarkers in schizophrenia.**

(a) C-Reactive Protein (CRP)

| Study | Patients (n) | Patients Mean | Patients SD | Controls (n) | Controls Mean | Controls SD |
|---|---|---|---|---|---|---|
| Bolu et al, 2019 [5] | 48 | 3.74 | 4.27 | 54 | 1.52 | 0.63 |
| De Berardis et al, 2013 [21] | 30 | 2.8 | 0.9 | 30 | 1.5 | 0.6 |
| Fernandez-Egea et al, 2009a [25] | 50 | 0.21 | 0.28 | 50 | 0.2 | 0.18 |
| Yesilkaya et al, 2024 [125] | 69 | 4.08 | 5.38 | 127 | 1.2 | 1.01 |
| Yuan et al, 2018 [127] | 41 | 0.77 | 0.94 | 41 | 0.44 | 0.46 |
| Zhu et al, 2019 [135] | 58 | 1.85 | 1.73 | 31 | 0.77 | 0.48 |

(b) Platelet

| Study | Patients (n) | Patients Mean | Patients SD | Controls (n) | Controls Mean | Controls SD |
|---|---|---|---|---|---|---|
| Cabello-Rangel et al, 2023 [9] | 175 | 242.69 | 56.9 | 52 | 295.8 | 74.7 |
| Huang et al, 2022 [41] | 53 | 242.47 | 52.17 | 59 | 229.64 | 53.4 |
| Kapici et al, 2023 [2] | 63 | 249.06 | 65.835 | 78 | 237.885 | 57.16 |
| Onur et al, 2021 [80] | 60 | 256 | 57.3 | 32 | 261.2 | 57.7 |
| Sahpolat et al, 2022 [92] | 37 | 278.1 | 48.47 | 43 | 271.37 | 58.76 |
| Yesilkaya et al, 2024 [125] | 69 | 268.75 | 68.1 | 127 | 251.2 | 60.1 |
| Yu et al, 2020 [126] | 106 | 252.22 | 6.36 | 120 | 218.26 | 2.72 |

(c) White blood cell (WBC)

| Study | Patients (n) | Patients Mean | Patients SD | Controls (n) | Controls Mean | Controls SD |
|---|---|---|---|---|---|---|
| Garcia-Rizo et al, 2019 [32] | 38 | 7.01 | 2.2 | 49 | 5.97 | 1.4 |
| Huang et al, 2022 [41] | 53 | 6.81 | 2.22 | 59 | 6.33 | 1.24 |
| Kapici et al, 2023 [2] | 63 | 8488 | 2022 | 78 | 8342 | 2052 |
| Onur et al, 2021 [80] | 60 | 8.7 | 1.9 | 32 | 6.8 | 1.5 |
| Sahpolat et al, 2022 [92] | 37 | 8.04 | 1.84 | 43 | 7.39 | 1.95 |
| Yesilkaya et al, 2024 [125] | 69 | 8.51 | 2.55 | 127 | 7.05 | 1.7 |

(d) Lymphocyte

| Study | Patients (n) | Patients Mean | Patients SD | Controls (n) | Controls Mean | Controls SD |
|---|---|---|---|---|---|---|
| Cabello-Rangel et al, 2023 [9] | 175 | 2.11 | 0.54 | 52 | 2.11 | 0.66 |
| Garcia-Rizo et al, 2019 [32] | 38 | 2.31 | 0.8 | 49 | 2.06 | 0.5 |
| Huang et al, 2022 [41] | 53 | 1.95 | 0.69 | 59 | 2.03 | 0.45 |
| Kapici et al, 2023 [2] | 63 | 2219 | 828 | 78 | 2698 | 1042 |
| Onur et al, 2021 [80] | 60 | 2.2 | 0.8 | 32 | 2.1 | 0.5 |
| Sahpolat et al, 2022 [92] | 37 | 2.1 | 0.57 | 43 | 2.4 | 0.64 |
| Yesilkaya et al, 2024 [125] | 69 | 2.3 | 0.83 | 127 | 2.39 | 0.6 |

(e) Monocyte

| Study | Patients (n) | Patients Mean | Patients SD | Controls (n) | Controls Mean | Controls SD |
|---|---|---|---|---|---|---|
| Garcia-Rizo et al, 2019 [32] | 38 | 0.43 | 0.2 | 49 | 0.36 | 0.1 |
| Huang et al, 2022 [41] | 53 | 0.42 | 0.23 | 59 | 0.31 | 0.08 |
| Kapici et al, 2023 [2] | 63 | 567 | 177 | 78 | 539 | 193 |
| Onur et al, 2021 [80] | 60 | 0.7 | 0.2 | 32 | 0.4 | 0.1 |
| Yesilkaya et al, 2024 [125] | 69 | 0.66 | 0.22 | 127 | 0.55 | 0.18 |

(f) Neutrophil

| Study | Patients (n) | Patients Mean | Patients SD | Controls (n) | Controls Mean | Controls SD |
|---|---|---|---|---|---|---|
| Cabello-Rangel et al, 2023 [9] | 175 | 3.78 | 1.29 | 52 | 4.01 | 1.3 |
| Garcia-Rizo et al, 2019 [32] | 38 | 4.24 | 1.9 | 49 | 3.51 | 1.2 |
| Huang et al, 2022 [41] | 53 | 4.47 | 2.13 | 59 | 3.82 | 0.99 |
| Kapici et al, 2023 [2] | 63 | 5476 | 1916 | 78 | 4932 | 1614 |
| Onur et al, 2021 [80] | 60 | 5.8 | 1.8 | 32 | 3.9 | 1.1 |
| Sahpolat et al, 2022 [92] | 37 | 5.27 | 1.62 | 43 | 4.33 | 1.53 |
| Yesilkaya et al, 2024 [125] | 69 | 5.18 | 2.3 | 127 | 3.89 | 1.22 |

(g) Monocyte-lymphocyte ratio (MLR)

| Study | Patients (n) | Patients Mean | Patients SD | Controls (n) | Controls Mean | Controls SD |
|---|---|---|---|---|---|---|
| Huang et al, 2022 [41] | 53 | 0.23 | 0.14 | 59 | 0.16 | 0.06 |
| Onur et al, 2021 [80] | 60 | 0.3 | 0.1 | 32 | 0.2 | 0.05 |
| Yesilkaya et al, 2024 [125] | 69 | 0.31 | 0.2 | 127 | 0.23 | 0.07 |
| Yu et al, 2020 [126] | 106 | 0.21 | 0.01 | 120 | 0.16 | 0 |

(h) Neutrophil-lymphocyte ratio (NLR)

| Study | Patients (n) | Patients Mean | Patients SD | Controls (n) | Controls Mean | Controls SD |
|---|---|---|---|---|---|---|
| Cabello-Rangel et al, 2023 [9] | 175 | 1.91 | 0.87 | 52 | 1.99 | 0.71 |
| Garcia-Rizo et al, 2019 [32] | 38 | 2.04 | 1.3 | 49 | 1.78 | 0.7 |
| Huang et al, 2022 [41] | 53 | 2.57 | 1.46 | 59 | 1.94 | 0.59 |
| Onur et al, 2021 [80] | 60 | 3 | 1.6 | 32 | 1.9 | 0.7 |
| Sahpolat et al, 2022 [92] | 37 | 2.56 | 0.91 | 43 | 1.88 | 0.78 |
| Yesilkaya et al, 2024 [125] | 69 | 2.79 | 3.35 | 127 | 1.85 | 0.95 |
| Yu et al, 2020 [126] | 106 | 2.44 | 0.11 | 120 | 1.78 | 0.05 |

(i) Platelet-lymphocyte ratio (PLR)

| Study | Patients (n) | Patients Mean | Patients SD | Controls (n) | Controls Mean | Controls SD |
|---|---|---|---|---|---|---|
| Huang et al, 2022 [41] | 53 | 134.16 | 41.01 | 59 | 116.49 | 30.96 |
| Onur et al, 2021 [80] | 60 | 130.9 | 60.4 | 32 | 125.5 | 36.1 |
| Sahpolat et al, 2022 [92] | 37 | 0.14 | 0.04 | 43 | 0.12 | 0.04 |
| Yesilkaya et al, 2024 [125] | 69 | 128.44 | 67.94 | 127 | 114.15 | 39.99 |
| Yu et al, 2020 [126] | 106 | 131.21 | 4.26 | 120 | 113.12 | 2.06 |

**Supplementary Table 10. Meta-analysis raw data of BMI in schizophrenia.**

(a) Body mass index (BMI)

| Study | Patients (n) | Patients Mean | Patients SD | Controls (n) | Controls Mean | Controls SD |
|---|---|---|---|---|---|---|
| Amoli et al, 2019 [3] | 15 | 21.48 | 6.81 | 15 | 23.18 | 4.78 |
| Bolu et al, 2019 [5] | 48 | 27 | 3.2 | 54 | 26.2 | 3.6 |
| Chen et al, 2018 [12] | 100 | 20.73 | 2.7 | 118 | 20.96 | 2.25 |
| Cropley et al, 2023 [18] | 24 | 22.6 | 3.3 | 25 | 22.3 | 3.1 |
| Dai et al, 2020 [19] | 46 | 21.8 | 2.8 | 60 | 22 | 3 |
| Dasgupta et al, 2010 [20] | 30 | 20.95 | 3.07 | 25 | 21.01 | 2.72 |
| Garrido-Torres et al, 2022 [33] | 244 | 23.16 | 4.26 | 166 | 26.03 | 4.44 |
| Kalmady et al, 2018 [49] | 75 | 18.97 | 3.66 | 102 | 22.21 | 2.73 |
| Keri et al, 2017 [51] | 42 | 23.2 | 4.1 | 42 | 22.7 | 4.3 |
| Khan, 2022 [52] | 22 | 23.2 | 2.14 | 14 | 25.1 | 2.61 |
| Kolenic et al, 2018 [54] | 120 | 23.32 | 4 | 114 | 22.6 | 2.93 |
| Lang et al, 2021 [58] | 430 | 23.66 | 2.77 | 453 | 23.02 | 2.14 |
| Li et al, 2022a [62] | 83 | 21.85 | 3.81 | 14 | 22.39 | 1.93 |
| Li et al, 2024a [63] | 56 | 22.02 | 3.7 | 108 | 22.01 | 3.69 |
| Li et al, 2024b [64] | 137 | 22.52 | 3.76 | 67 | 24.59 | 5.19 |
| Li et al, 2025 [68] (females) | 89 | 22.13 | 4.21 | 17 | 20.21 | 1.95 |
| Li et al, 2025 [68] (males) | 83 | 21.85 | 3.81 | 14 | 22.39 | 1.93 |
| Liang et al, 2019 [70] | 49 | 21.84 | 2.88 | 71 | 22.76 | 1.67 |
| Liang et al, 2022 [71] | 92 | 22.55 | 3.91 | 59 | 22.47 | 3.36 |
| Liu et al, 2015 [73] | 35 | 22.6 | 11.54 | 35 | 22.21 | 10.18 |
| Misiak et al, 2016 [78] | 135 | 23 | 3.2 | 146 | 22.6 | 3.2 |
| Pan et al, 2022 [81] | 75 | 20.9 | 2.91 | 44 | 23.35 | 3.67 |
| Parksepp et al, 2022 [37] | 54 | 22.6 | 2.8 | 58 | 22.8 | 3 |
| Ryan et al, 2003 [88] | 26 | 24.5 | 3.6 | 26 | 24.6 | 3.7 |
| Saddichha et al, 2008a [89] | 99 | 19.4 | 3 | 51 | 19.5 | 2.3 |
| Saloojee et al, 2018 [93] | 67 | 24.4 | 3.5 | 67 | 24.6 | 5.3 |
| Sayed et al, 2023 [94] | 150 | 29.55 | 4.39 | 120 | 21.68 | 2.32 |
| Sengupta et al, 2008 [95] | 38 | 22.8 | 3.2 | 36 | 23.9 | 3.5 |
| Verma et al, 2009 [111] | 160 | 21.2 | 3.7 | 200 | 23.5 | 4.4 |
| Wang et al, 2023 [102] | 148 | 20.86 | 2.81 | 97 | 21.12 | 2.61 |
| Wu et al, 2013 [114] | 70 | 19.63 | 2.53 | 44 | 20.34 | 2.72 |
| Xiong et al, 2011 [115] | 30 | 23.01 | 1.67 | 28 | 22.45 | 1.23 |
| Xiu et al, 2016 [116] | 256 | 20.1 | 2.8 | 540 | 22 | 4.9 |
| Xiu et al, 2018 [117] | 45 | 20.1 | 2.8 | 40 | 24.5 | 5.6 |
| Xiu et al, 2023 [118] | 43 | 21.3 | 3.1 | 29 | 21.4 | 3.2 |
| Yang et al, 2018 [121] | 79 | 19.51 | 2.64 | 35 | 20.49 | 0.88 |
| Zhang et al, 2016a [130] | 31 | 22.5 | 3.2 | 71 | 22.8 | 1.7 |
| Zhou et al, 2021 [133] (females) | 134 | 23.29 | 3.46 | 55 | 21.3 | 2.3 |
| Zhou et al, 2021 [133] (males) | 123 | 24.3 | 3.68 | 63 | 22.42 | 2.35 |
| Zhu et al, 2020 [136] | 119 | 22.75 | 3.26 | 135 | 23.23 | 3.48 |

(b) Waist-hip circumference ratio (WHR)

| Study | Patients (n) | Patients Mean | Patients SD | Controls (n) | Controls Mean | Controls SD |
|---|---|---|---|---|---|---|
| Chen et al, 2018 [12] | 100 | 0.82 | 0.06 | 118 | 0.82 | 0.07 |
| Chen et al, 2021a [13] | 113 | 0.84 | 0.08 | 58 | 0.82 | 0.07 |
| Keri et al, 2017 [51] | 42 | 0.79 | 0.12 | 42 | 0.79 | 0.13 |
| Li et al, 2025 [68] (females) | 89 | 0.82 | 0.07 | 17 | 0.8 | 0.04 |
| Li et al, 2025 [68] (males) | 83 | 0.86 | 0.06 | 14 | 0.83 | 0.06 |
| Sengupta et al, 2008 [95] | 38 | 0.87 | 0.06 | 36 | 0.82 | 0.06 |
| Wu et al, 2013 [114] | 70 | 0.82 | 0.06 | 44 | 0.79 | 0.06 |

**Supplementary Table 11. Meta-analysis raw data of glucose metabolism in schizophrenia.**

(a) Fasting glucose

| Study | Patients (n) | Patients Mean | Patients SD | Controls (n) | Controls Mean | Controls SD |
|---|---|---|---|---|---|---|
| Chen et al, 2016 [11] | 172 | 5 | 1 | 31 | 4.3 | 0.4 |
| Chen et al, 2018 [5,12] | 100 | 4.75 | 0.74 | 118 | 4.18 | 0.55 |
| Dasgupta et al, 2010 [20] | 30 | 5.33 | 0.78 | 25 | 5.07 | 0.55 |
| Fernandez-Egea et al, 2009a [25] | 50 | 4.55 | 0.66 | 50 | 4.65 | 0.38 |
| Garrido-Torres et al, 2022 [33] | 244 | 86.36 | 21.5 | 166 | 84.49 | 8.54 |
| Kapogiannis et al, 2019 [44] | 24 | 5.37 | 1.05 | 24 | 4.71 | 0.56 |
| Lang et al, 2021 [58] | 430 | 5.13 | 0.85 | 453 | 4.95 | 0.71 |
| Li et al, 2022a [62] | 83 | 4.98 | 1.04 | 14 | 4.49 | 0.28 |
| Li et al, 2025 [68] (females) | 89 | 5.05 | 0.9 | 17 | 4.21 | 0.4 |
| Li et al, 2025 [68] (males) | 83 | 4.98 | 1.04 | 14 | 4.49 | 0.28 |
| Liang et al, 2019 [70] | 49 | 4.66 | 0.61 | 71 | 4.6 | 0.49 |
| Liang et al, 2022 [71] | 92 | 5.11 | 1.09 | 59 | 5.49 | 0.47 |
| Misiak et al, 2014 [77] | 35 | 86.08 | 9.87 | 53 | 86.74 | 10.83 |
| Misiak et al, 2016 [78] | 135 | 86.2 | 9 | 146 | 83.9 | 10.6 |
| Petrikis et al, 2015a [82] | 40 | 87.55 | 11.24 | 40 | 86.67 | 6.35 |
| Radu et al, 2020 [86] | 50 | 97.46 | 10.85 | 50 | 97.24 | 10.32 |
| Ryan et al, 2003 [88] | 26 | 95.8 | 16.9 | 26 | 88.2 | 5.4 |
| Saddichha et al, 2008b [90] | 99 | 82.2 | 10.7 | 51 | 80.8 | 6.3 |
| Saloojee et al, 2018 [93] | 67 | 4.7 | 0.7 | 67 | 4.8 | 0.4 |
| Sayed et al, 2023 [94] | 150 | 84.68 | 11.28 | 120 | 152.34 | 26.22 |
| Sengupta et al, 2008 [95] | 38 | 4.69 | 0.53 | 36 | 4.68 | 0.5 |
| Tao et al, 2020 [107] | 90 | 4.4 | 0.5 | 70 | 4.3 | 0.7 |
| Van Nimwegen et al, 2008 [109] | 7 | 5.1 | 0.5 | 7 | 4.8 | 0.3 |
| Venkatasubramanian et al, 2007 [110] | 44 | 94.8 | 19 | 44 | 92.2 | 9.4 |
| Wang et al, 2023 [102] | 148 | 4.49 | 0.47 | 97 | 4.22 | 0.83 |
| Wani et al, 2015 [113] | 50 | 88.34 | 9.31 | 50 | 87.4 | 6.55 |
| Wu et al, 2013 [114] | 70 | 4.79 | 0.71 | 44 | 4.77 | 0.46 |
| Xiu et al, 2023 [118] | 43 | 4.9 | 1.2 | 29 | 4.3 | 0.5 |
| Yuan et al, 2018 [127] | 41 | 4.37 | 0.52 | 41 | 4.4 | 0.97 |
| Zhang et al, 2016a [130] | 31 | 4.67 | 0.47 | 71 | 4.61 | 0.48 |
| Zhang et al, 2020 [129] | 39 | 4.92 | 0.67 | 31 | 4.33 | 0.37 |
| Zhou et al, 2021 [133] (females) | 134 | 5.33 | 0.87 | 55 | 4.97 | 0.45 |
| Zhou et al, 2021 [133] (males) | 123 | 5.28 | 0.83 | 63 | 5.19 | 0.55 |

(b) 2-hour glucose

| Study | Patients (n) | Patients Mean | Patients SD | Controls (n) | Controls Mean | Controls SD |
|---|---|---|---|---|---|---|
| Chen et al, 2016 [11] | 172 | 6.7 | 2.4 | 31 | 4.7 | 1.2 |
| Chen et al, 2018 [12] | 100 | 6.5 | 1.79 | 118 | 4.98 | 1.31 |
| Fernandez-Egea et al, 2009a [25] | 50 | 6.1 | 1.93 | 50 | 4.49 | 1.06 |
| Li et al, 2022a [62] | 83 | 6.48 | 2.51 | 14 | 5.21 | 0.88 |
| Li et al, 2025 [68] (females) | 89 | 7.07 | 2.27 | 17 | 4.31 | 1.22 |
| Li et al, 2025 [68] (males) | 83 | 6.48 | 2.51 | 14 | 5.21 | 0.88 |
| Saddichha et al, 2008b [90] | 99 | 95.6 | 12.3 | 51 | 90.9 | 7.9 |
| Wani et al, 2015 [113] | 50 | 130.26 | 12.53 | 50 | 129.98 | 4.74 |

(c) Insulin

| Study | Patient s (n) | Patient s Mean | Patient s SD | Control s (n) | Control s Mean | Control s SD |
|---|---|---|---|---|---|---|
| Chen et al, 2016 [11] | 172 | 5.1 | 2.4 | 31 | 4.4 | 1.2 |
| Chen et al, 2018 [5,12] | 100 | 8.57 | 4.88 | 118 | 6.57 | 3.77 |
| Fernandez-Egea et al, 2009a [25] | 50 | 10.3 | 7.3 | 50 | 9.6 | 3.8 |
| Kapogiannis et al, 2019 [44] | 24 | 5.82 | 7.25 | 24 | 2.51 | 2.06 |
| Lang et al, 2021 [58] | 430 | 91.6 | 46.95 | 453 | 60.06 | 31.81 |
| Li et al, 2025 [68] (females) | 89 | 5.47 | 2.87 | 17 | 4.21 | 1.2 |
| Li et al, 2025 [68] (males) | 83 | 5.57 | 3.12 | 14 | 4.63 | 1.21 |
| Liang et al, 2019 [70] | 49 | 11.37 | 10.76 | 71 | 7.53 | 3.75 |
| Parksepp et al, 2022 [37] | 54 | 12.9 | 14.8 | 58 | 15.6 | 14.3 |
| Ryan et al, 2003 [88] | 26 | 9.8 | 3.9 | 26 | 7.7 | 3.7 |
| Sengupta et al, 2008 [95] | 38 | 16.7 | 6.5 | 36 | 18 | 10.9 |
| Tao et al, 2020 [107] | 90 | 11.1 | 3.3 | 70 | 6.1 | 1.1 |
| Van Nimwegen et al, 2008 [109] | 7 | 49 | 18 | 7 | 29 | 12 |
| Venkatasubramanian et al, 2007 [110] | 44 | 9.5 | 6.4 | 44 | 7.1 | 3.6 |
| Wu et al, 2013 [114] | 70 | 8.49 | 4.92 | 44 | 5.96 | 2.15 |
| Xiu et al, 2023 [118] | 43 | 5.7 | 3.4 | 29 | 4.4 | 1.7 |
| Zhang et al, 2016a [130] | 31 | 10.32 | 7.27 | 71 | 7.52 | 3.75 |
| Zhang et al, 2020 [129] | 39 | 5.6 | 3.11 | 31 | 4.4 | 1.2 |
| Zhou et al, 2021 [133] (females) | 134 | 13.13 | 6.15 | 55 | 8.25 | 3.23 |
| Zhou et al, 2021 [133] (males) | 123 | 15.03 | 6.94 | 63 | 7.89 | 3.05 |

(d) Homeostatic Model Assessment for Insulin Resistance (HOMA-IR)

| Study | Patients (n) | Patients Mean | Patients SD | Controls (n) | Controls Mean | Controls SD |
|---|---|---|---|---|---|---|
| Chen et al, 2016 [11] | 172 | 1.2 | 0.7 | 31 | 0.8 | 0.3 |
| Chen et al, 2018 [5,12] | 100 | 1.86 | 1.06 | 118 | 1.25 | 0.78 |
| Dasgupta et al, 2010 [20] | 30 | 1.34 | 0.5 | 25 | 1.12 | 0.18 |
| Fernandez-Egea et al, 2009a [25] | 50 | 1.48 | 1.08 | 50 | 1.4 | 0.56 |
| Kapogiannis et al, 2019 [44] | 24 | 1.46 | 1.78 | 24 | 0.52 | 0.42 |
| Lang et al, 2021 [58] | 430 | 3.03 | 1.75 | 453 | 2.16 | 1.1 |
| Li et al, 2025 [68] (females) | 89 | 1.18 | 0.63 | 17 | 0.79 | 0.26 |
| Li et al, 2025 [68] (males) | 83 | 1.25 | 0.81 | 14 | 0.93 | 0.26 |
| Ryan et al, 2003 [88] | 26 | 2.3 | 1 | 26 | 1.7 | 0.7 |
| Sengupta et al, 2008 [95] | 38 | 3.65 | 1.6 | 36 | 4 | 2.6 |
| Tao et al, 2020 [107] | 90 | 2.2 | 0.7 | 70 | 1.2 | 0.3 |
| Venkatasubramanian et al, 2007 [110] | 44 | 1.2 | 0.8 | 44 | 0.9 | 0.4 |
| Wu et al, 2013 [114] | 70 | 1.83 | 1.16 | 44 | 1.28 | 0.52 |
| Xiu et al, 2023 [118] | 43 | 1.1 | 0.7 | 29 | 0.85 | 0.3 |
| Zhang et al, 2020 [129] | 39 | 1.21 | 0.71 | 31 | 0.85 | 0.26 |
| Zhou et al, 2021 [133] (females) | 134 | 3.17 | 1.78 | 55 | 1.81 | 0.69 |
| Zhou et al, 2021 [133] (males) | 123 | 3.51 | 1.71 | 63 | 1.81 | 0.67 |

(e) Glycated haemoglobin (HbA1c)

| Study | Patients (n) | Patients Mean | Patients SD | Controls (n) | Controls Mean | Controls SD |
|---|---|---|---|---|---|---|
| Fernandez-Egea et al, 2009a [25] | 41 | 4.4 | 0.38 | 41 | 4.4 | 0.26 |
| Lang et al, 2021 [58] | 430 | 5.57 | 0.68 | 453 | 5.3 | 0.53 |
| Petrikis et al, 2015a [82] | 40 | 5.14 | 0.43 | 40 | 5.25 | 0.42 |
| Sayed et al, 2023 [94] | 150 | 9.13 | 2.29 | 120 | 5.91 | 0.95 |
| Sengupta et al, 2008 [95] | 38 | 0.05 | 0.005 | 36 | 0.05 | 0.004 |
| Zhou et al, 2021 [133] (females) | 134 | 5.57 | 0.73 | 55 | 5.42 | 0.55 |
| Zhou et al, 2021 [133] (males) | 123 | 5.65 | 0.66 | 63 | 5.39 | 0.41 |

**Supplementary Table 12. Meta-analysis raw data of lipid metabolism in schizophrenia.**

(a) Cholestrol

| Study | Patients (n) | Patients Mean | Patients SD | Controls (n) | Controls Mean | Controls SD |
|---|---|---|---|---|---|---|
| Chen et al, 2016 [11] | 172 | 4.4 | 0.9 | 31 | 4.2 | 1 |
| Chen et al, 2018 [5,12] | 100 | 4.01 | 0.83 | 118 | 4.12 | 0.84 |
| Dasgupta et al, 2010 [20] | 30 | 157.6 | 42.88 | 25 | 169.2 | 17.08 |
| Dongxia et al, 2024 [24] | 123 | 4.22 | 1.08 | 38 | 3.96 | 1.12 |
| Kirkpatrick et al, 2010 [29] | 76 | 168.1 | 34.8 | 76 | 175.4 | 32.2 |
| Lang et al, 2021 [58] | 430 | 4.27 | 1 | 453 | 4.28 | 0.93 |
| Li et al, 2022a [62] | 83 | 4.43 | 0.9 | 14 | 4.3 | 0.97 |
| Li et al, 2025 [68] (females) | 89 | 4.83 | 0.93 | 17 | 4.15 | 1 |
| Li et al, 2025 [68] (males) | 83 | 4.43 | 0.9 | 14 | 4.3 | 0.97 |
| Liang et al, 2019 [70] | 49 | 4.16 | 0.72 | 71 | 4.18 | 0.66 |
| Liang et al, 2022 [71] | 92 | 4.33 | 1.02 | 59 | 4.72 | 0.76 |
| Misiak et al, 2014 [77] | 35 | 173 | 30.19 | 53 | 170.81 | 30.89 |
| Misiak et al, 2016 [78] | 135 | 169 | 34.5 | 146 | 173.5 | 34.3 |
| Petrikis et al, 2015a [82] | 40 | 188.21 | 46.92 | 40 | 196.96 | 43.57 |
| Ryan et al, 2003 [88] | 26 | 4.02 | 0.78 | 26 | 4.57 | 0.81 |
| Saloojee et al, 2018 [93] | 67 | 3.7 | 0.8 | 67 | 3.8 | 0.7 |
| Sayed et al, 2023 [94] | 150 | 234.72 | 41.25 | 120 | 127.13 | 11.88 |
| Sengupta et al, 2008 [95] | 38 | 4.04 | 0.77 | 36 | 4.17 | 0.9 |
| Venkatasubramanian et al, 2007 [110] | 44 | 168.6 | 32.8 | 44 | 172.2 | 32.4 |
| Verma et al, 2009 [111] | 160 | 4.7 | 1 | 200 | 5.1 | 0.9 |
| Wang et al, 2023 [102] | 148 | 3.67 | 0.73 | 97 | 3.73 | 0.68 |
| Wu et al, 2013 [114] | 70 | 4.16 | 0.77 | 44 | 4.54 | 0.76 |
| Xiu et al, 2023 [118] | 43 | 4.7 | 0.9 | 29 | 4.4 | 1.2 |
| Zhang et al, 2016a [130] | 31 | 4.01 | 0.76 | 71 | 4.18 | 0.66 |
| Zhang et al, 2020 [129] | 39 | 4.65 | 0.85 | 31 | 4.22 | 0.97 |
| Zhou et al, 2021 [133] (females) | 134 | 4.26 | 1.05 | 55 | 4.32 | 0.99 |
| Zhou et al, 2021 [133] (males) | 123 | 4.44 | 1.08 | 63 | 4.35 | 1.01 |

(b) Triglycerides

| Study | Patients (n) | Patients Mean | Patients SD | Controls (n) | Controls Mean | Controls SD |
|---|---|---|---|---|---|---|
| Chen et al, 2016 [11] | 172 | 1.2 | 0.7 | 31 | 1.1 | 0.7 |
| Chen et al, 2018 [5,12] | 100 | 0.94 | 0.53 | 118 | 0.99 | 1.138 |
| Dasgupta et al, 2010 [20] | 30 | 145.96 | 71.83 | 25 | 138.64 | 20.99 |
| Dongxia et al, 2024 [24] | 123 | 1.75 | 0.13 | 38 | 1.51 | 0.12 |
| Garrido-Torres et al, 2022 [33] | 244 | 85.27 | 38.76 | 166 | 86.6 | 40.94 |
| Kirkpatrick et al, 2010 [29] | 76 | 84.9 | 38.6 | 76 | 87.5 | 52.2 |
| Kolenic et al, 2018 [54] | 120 | 1.32 | 0.56 | 114 | 1.1 | 0.45 |
| Lang et al, 2021 [58] | 430 | 1.58 | 0.94 | 453 | 1.25 | 0.47 |
| Li et al, 2022a [62] | 83 | 1.12 | 0.51 | 14 | 1.06 | 0.62 |
| Li et al, 2025 [68] (females) | 89 | 1.03 | 0.57 | 17 | 1.08 | 0.76 |
| Li et al, 2025 [68] (males) | 83 | 1.12 | 0.51 | 14 | 1.06 | 0.62 |
| Liang et al, 2019 [70] | 49 | 1.18 | 0.57 | 71 | 0.92 | 0.42 |
| Liang et al, 2022 [71] | 92 | 1.43 | 1.2 | 59 | 1.12 | 0.46 |
| Misiak et al, 2014 [77] | 35 | 110.09 | 53.31 | 53 | 95.11 | 45.42 |
| Misiak et al, 2016 [78] | 135 | 116.7 | 63.4 | 146 | 96.8 | 49.1 |
| Petrikis et al, 2015a [82] | 40 | 93.29 | 51.87 | 40 | 85.54 | 40.88 |
| Radu et al, 2020 [86] | 50 | 140.9 | 48.89 | 50 | 141.5 | 43.43 |
| Ryan et al, 2003 [88] | 26 | 0.99 | 0.43 | 26 | 0.92 | 0.3 |
| Saddichha et al, 2008c [91] | 99 | 116.3 | 16.9 | 51 | 101.3 | 46.4 |
| Saloojee et al, 2018 [93] | 67 | 0.7 | 0.3 | 67 | 0.7 | 0.4 |
| Sayed et al, 2023 [94] | 150 | 260.66 | 49.77 | 120 | 132.33 | 19.03 |
| Sengupta et al, 2008 [95] | 38 | 1.1 | 0.65 | 36 | 1 | 0.72 |
| Venkatasubramanian et al, 2007 [110] | 44 | 116 | 57.3 | 44 | 105.5 | 56 |
| Wang et al, 2023 [102] | 148 | 0.87 | 0.41 | 97 | 0.97 | 0.5 |
| Wu et al, 2013 [114] | 70 | 1.12 | 0.75 | 44 | 0.99 | 0.41 |
| Xiu et al, 2023 [118] | 43 | 1.1 | 0.5 | 29 | 0.9 | 0.3 |
| Yuan et al, 2018 [127] | 41 | 0.97 | 0.67 | 41 | 0.83 | 0.4 |
| Zhang et al, 2016a [130] | 31 | 1.05 | 0.65 | 71 | 0.92 | 0.42 |
| Zhang et al, 2020 [129] | 39 | 1.07 | 0.52 | 31 | 1.07 | 0.69 |
| Zhou et al, 2021 [133] (females) | 134 | 1.49 | 1.01 | 55 | 1.1 | 0.43 |
| Zhou et al, 2021 [133] (males) | 123 | 1.98 | 1.23 | 63 | 1.21 | 0.53 |

(c) High-density lipoprotein (HDL)

| Study | Patients (n) | Patients Mean | Patients SD | Controls (n) | Controls Mean | Controls SD |
|---|---|---|---|---|---|---|
| Chen et al, 2016 [11] | 172 | 1.4 | 0.3 | 31 | 1.4 | 0.2 |
| Chen et al, 2018 [5,12] | 100 | 1.31 | 0.32 | 118 | 1.5 | 0.3 |
| Dasgupta et al, 2010 [20] | 30 | 39.26 | 7.51 | 25 | 40.72 | 4.1 |
| Dongxia et al, 2024 [24] | 123 | 1.41 | 0.43 | 38 | 1.47 | 0.63 |
| Garrido-Torres et al, 2022 [33] | 244 | 53.55 | 15.76 | 166 | 57.71 | 15.5 |
| Kirkpatrick et al, 2010 [29] | 76 | 51.5 | 17.3 | 76 | 52 | 12.6 |
| Kolenic et al, 2018 [54] | 120 | 1.34 | 0.37 | 114 | 1.55 | 0.38 |
| Lang et al, 2021 [58] (females) | 222 | 1.4 | 0.33 | 256 | 1.57 | 0.33 |
| Lang et al, 2021 [58] (males) | 208 | 1.27 | 0.42 | 197 | 1.41 | 0.4 |
| Li et al, 2022a [62] | 83 | 1.42 | 0.35 | 14 | 1.49 | 0.36 |
| Li et al, 2025 [68] (females) | 89 | 1.58 | 0.35 | 17 | 1.4 | 0.29 |
| Li et al, 2025 [68] (males) | 83 | 1.42 | 0.35 | 14 | 1.5 | 0.36 |
| Liang et al, 2019 [70] | 49 | 2.18 | 0.94 | 71 | 1.41 | 0.3 |
| Liang et al, 2022 [71] | 92 | 1.38 | 0.39 | 59 | 1.27 | 0.29 |
| Misiak et al, 2014 [77] | 35 | 55.1 | 18.56 | 53 | 66.07 | 17.46 |
| Misiak et al, 2016 [78] | 135 | 52.4 | 14.6 | 146 | 65.4 | 18.9 |
| Petrikis et al, 2015a [82] | 40 | 51.3 | 10.75 | 40 | 61.68 | 16.53 |
| Radu et al, 2020 [86] | 50 | 37.04 | 10.22 | 50 | 36.72 | 9.01 |
| Ryan et al, 2003 [88] | 26 | 1.2 | 0.44 | 26 | 1.25 | 0.25 |
| Saddichha et al, 2008c [91] | 99 | 35.6 | 10 | 51 | 36.4 | 8.9 |
| Saloojee et al, 2018 [93] | 67 | 1.2 | 0.3 | 67 | 1.2 | 0.3 |
| Sayed et al, 2023 [94] | 150 | 34.68 | 11.02 | 120 | 93.71 | 16.52 |
| Sengupta et al, 2008 [95] | 38 | 1.06 | 0.26 | 36 | 1.19 | 0.36 |
| Verma et al, 2009 [111] | 160 | 1.5 | 0.4 | 200 | 1.5 | 0.3 |
| Wang et al, 2023 [102] | 148 | 1.32 | 0.3 | 97 | 1.31 | 0.36 |
| Wu et al, 2013 [114] | 70 | 1.29 | 0.26 | 44 | 1.58 | 0.31 |
| Xiu et al, 2023 [118] | 43 | 1.6 | 0.3 | 29 | 1.4 | 0.3 |
| Yuan et al, 2018 [127] | 41 | 1.32 | 0.29 | 41 | 1.3 | 0.25 |
| Zhang et al, 2016a [130] | 31 | 1.19 | 0.33 | 71 | 1.41 | 0.3 |
| Zhang et al, 2020 [129] | 39 | 1.48 | 0.35 | 31 | 1.44 | 0.32 |
| Zhou et al, 2021 [133] (females) | 134 | 1.32 | 0.34 | 55 | 1.72 | 0.49 |
| Zhou et al, 2021 [133] (males) | 123 | 1.19 | 0.31 | 63 | 1.49 | 0.36 |

(d) Low-density lipoprotein (LDL)

| Study | Patients (n) | Patients Mean | Patients SD | Controls (n) | Controls Mean | Controls SD |
|---|---|---|---|---|---|---|
| Chen et al, 2016 [11] | 172 | 2.6 | 0.7 | 31 | 2.7 | 0.8 |
| Chen et al, 2018 [5,12] | 100 | 2.31 | 0.7 | 118 | 2.16 | 0.5 |
| Dasgupta et al, 2010 [20] | 30 | 89.14 | 37.01 | 25 | 101.55 | 18.43 |
| Dongxia et al, 2024 [24] | 123 | 3.1 | 0.95 | 38 | 3.12 | 0.62 |
| Kirkpatrick et al, 2010 [29] | 76 | 99.6 | 31.4 | 76 | 105.8 | 28.8 |
| Kolenic et al, 2018 [54] | 120 | 2.61 | 0.73 | 114 | 2.24 | 0.56 |
| Lang et al, 2021 [58] | 430 | 2.63 | 0.81 | 453 | 2.59 | 0.72 |
| Li et al, 2022a [62] | 83 | 2.51 | 0.82 | 14 | 2.47 | 0.35 |
| Li et al, 2025 [68] (females) | 89 | 2.87 | 0.76 | 17 | 2.51 | 0.96 |
| Li et al, 2025 [68] (males) | 83 | 2.51 | 0.82 | 14 | 2.47 | 0.35 |
| Liang et al, 2019 [70] | 49 | 1.91 | 0.85 | 71 | 1.58 | 0.64 |
| Liang et al, 2022 [71] | 92 | 2.47 | 0.74 | 59 | 2.64 | 0.6 |
| Misiak et al, 2014 [77] | 35 | 96.21 | 29.77 | 53 | 85.68 | 24.65 |
| Misiak et al, 2016 [78] | 135 | 92.8 | 28.3 | 146 | 90.4 | 29.2 |
| Radu et al, 2020 [86] | 50 | 120.2 | 21.81 | 50 | 122.9 | 22.58 |
| Ryan et al, 2003 [88] | 26 | 2.39 | 0.84 | 26 | 2.91 | 0.69 |
| Saloojee et al, 2018 [93] | 67 | 2.3 | 1.2 | 67 | 2.2 | 0.7 |
| Sayed et al, 2023 [94] | 150 | 89.47 | 14.5 | 120 | 64.58 | 9.05 |
| Sengupta et al, 2008 [95] | 38 | 2.48 | 0.63 | 36 | 2.52 | 0.72 |
| Verma et al, 2009 [111] | 160 | 2.7 | 0.9 | 200 | 3.1 | 0.8 |
| Wang et al, 2023 [102] | 148 | 2.12 | 0.6 | 97 | 2.04 | 0.5 |
| Wu et al, 2013 [114] | 70 | 2.5 | 0.67 | 44 | 2.62 | 0.63 |
| Xiu et al, 2023 [118] | 43 | 2.6 | 0.7 | 29 | 2.4 | 0.9 |
| Yuan et al, 2018 [127] | 41 | 2.22 | 0.64 | 41 | 2.01 | 0.63 |
| Zhang et al, 2016a [130] | 31 | 2.46 | 0.72 | 71 | 2.61 | 0.71 |
| Zhang et al, 2020 [129] | 39 | 2.66 | 0.74 | 31 | 2.49 | 0.74 |
| Zhou et al, 2021 [133] (females) | 134 | 2.64 | 0.82 | 55 | 2.62 | 0.98 |
| Zhou et al, 2021 [133] (males) | 123 | 2.74 | 0.8 | 63 | 2.55 | 0.83 |

**Supplementary Table 13. Meta-analysis raw data of liver function in schizophrenia.**

(a) Albumin

| Study | Patients (n) | Patients Mean | Patients SD | Controls (n) | Controls Mean | Controls SD |
|---|---|---|---|---|---|---|
| Huang et al, 2022 [41] | 53 | 43 | 6.06 | 59 | 46.1 | 2.66 |
| Reddy et al, 2003 [87] | 31 | 4.58 | 0.82 | 40 | 4.97 | 0.58 |
| Yesilkaya et al, 2024 [125] | 69 | 4.21 | 0.2 | 127 | 15.5 | 16.1 |

(b) Bilirubin

| Study | Patients (n) | Patients Mean | Patients SD | Controls (n) | Controls Mean | Controls SD |
|---|---|---|---|---|---|---|
| Huang et al, 2022 [41] | 53 | 12.72 | 7.04 | 59 | 10.52 | 4.58 |
| Reddy et al, 2003 [87] | 31 | 0.52 | 0.3 | 40 | 0.75 | 0.49 |
| Wang et al, 2023 [102] | 148 | 10.4 | 4.97 | 97 | 8.6 | 4.07 |

**Supplementary Table 14. Meta-analysis raw data of cardiovascular function in schizophrenia.**

(a) Systolic blood pressure (SBP)

| Study | Patients (n) | Patients Mean | Patients SD | Controls (n) | Controls Mean | Controls SD |
|---|---|---|---|---|---|---|
| Fernandez-Egea et al, 2009b [26] | 41 | 120.1 | 11.7 | 41 | 118.8 | 11.9 |
| Garrido-Torres et al, 2022 [33] | 244 | 119.08 | 15.11 | 166 | 106.14 | 12.08 |
| Lang et al, 2021 [58] | 430 | 119.75 | 13.23 | 453 | 114.03 | 11.53 |
| Misiak et al, 2016 [78] | 135 | 119.7 | 14.1 | 146 | 117.9 | 12.5 |
| Radu et al, 2020 [86] | 50 | 133.5 | 9.5 | 50 | 134.1 | 7.6 |
| Saddichha et al, 2008c [91] | 99 | 117.9 | 10.3 | 51 | 117 | 5.9 |
| Saloojee et al, 2018 [93] | 67 | 112.9 | 11.2 | 67 | 113.7 | 13.5 |
| Sayed et al, 2023 [94] | 150 | 150.92 | 11.7 | 120 | 125.57 | 7.07 |
| Tai et al, 2020 [104] | 42 | 113.4 | 13.41 | 42 | 109.17 | 8.45 |
| Zhou et al, 2021 [133] (females) | 134 | 121.63 | 13.37 | 55 | 110.44 | 12.31 |
| Zhou et al, 2021 [133] (males) | 123 | 125.74 | 13.72 | 63 | 110.33 | 10.89 |

(b) Diastolic blood pressure (DBP)

| Study | Patients (n) | Patients Mean | Patients SD | Controls (n) | Controls Mean | Controls SD |
|---|---|---|---|---|---|---|
| Fernandez-Egea et al, 2009b [26] | 41 | 72.2 | 9.5 | 41 | 77 | 8.5 |
| Garrido-Torres et al, 2022 [33] | 244 | 70.65 | 11.61 | 166 | 62.24 | 8.77 |
| Lang et al, 2021 [58] | 430 | 77.64 | 8.86 | 453 | 74.97 | 7.11 |
| Misiak et al, 2016 [78] | 135 | 73.1 | 10.5 | 146 | 74.6 | 10.3 |
| Radu et al, 2020 [86] | 50 | 81.4 | 6.2 | 50 | 82.2 | 5.8 |
| Saddichha et al, 2008c [91] | 99 | 77.7 | 7.5 | 51 | 77.7 | 5.5 |
| Saloojee et al, 2018 [93] | 67 | 72.1 | 8.2 | 67 | 70.7 | 12.2 |
| Tai et al, 2020 [104] | 42 | 62.31 | 11.01 | 42 | 61.02 | 7.35 |
| Zhou et al, 2021 [133] (females) | 134 | 77.63 | 8.95 | 55 | 72.09 | 8.4 |
| Zhou et al, 2021 [133] (males) | 123 | 79.62 | 10.16 | 63 | 72.11 | 7.28 |

**Supplementary Table 15. Meta-analysis raw data of pro-inflammatory biomarkers in major depressive disorder.**

(a) IFN-γ

| Study | Patients (n) | Patients Mean | Patients SD | Controls (n) | Controls Mean | Controls SD |
|---|---|---|---|---|---|---|
| Gao et al, 2024 [31] | 23 | 15.86 | 4.08 | 23 | 10.36 | 2.39 |
| Kakeda et al, 2018 [47] | 40 | 9.636 | 15 | 47 | 7.459 | 13.4 |
| Lan et al, 2022 [57] (females) | 72 | 1.3 | 0.3 | 25 | 1 | 0.24 |
| Lan et al, 2022 [57] (males) | 55 | 1.21 | 0.28 | 35 | 1 | 0.3 |
| Sugimoto et al, 2018 [103] | 35 | 10.455 | 15.861 | 35 | 8.969 | 15.225 |

(b) IL-1β

| Study | Patients (n) | Patients Mean | Patients SD | Controls (n) | Controls Mean | Controls SD |
|---|---|---|---|---|---|---|
| Gao et al, 2024 [31] | 23 | 7.55 | 2.49 | 23 | 6.39 | 0.94 |
| Kakeda et al, 2018 [47] | 40 | 0.041 | 0.048 | 47 | 0.053 | 0.087 |
| Kim et al, 2021 [53] (females) | 27 | 0.5555 | 0.3033 | 26 | 0.3455 | 0.2777 |
| Lan et al, 2022 [57] (females) | 72 | 0.34 | 0.41 | 25 | 0.11 | 0.38 |
| Lan et al, 2022 [57] (males) | 55 | 0.33 | 0.35 | 35 | 0.12 | 0.46 |
| Leo et al, 2006 [60] | 46 | 1.58 | 1.23 | 46 | 0.67 | 0.44 |
| Liu et al, 2022 [74] | 66 | 195.02 | 39.58 | 43 | 192.69 | 40.98 |
| Sugimoto et al, 2018 [103] | 35 | 0.038 | 0.044 | 35 | 0.058 | 0.098 |
| Yang et al, 2021 [122] | 34 | 3.8 | 0.11 | 34 | 3.93 | 0.16 |
| Zhao et al, 2022 [132] | 24 | 858.3 | 432.7 | 26 | 359.52 | 160.63 |
| Zou et al, 2018 [138] | 75 | 0.87 | 0.27 | 102 | 0.38 | 0.21 |

(c) IL-6

| Study | Patients (n) | Patients Mean | Patients SD | Controls (n) | Controls Mean | Controls SD |
|---|---|---|---|---|---|---|
| Chen et al, 2022a [15] | 29 | 6.65 | 4.62 | 25 | 7.89 | 3.84 |
| Kakeda et al, 2018 [47] | 40 | 0.85 | 1.808 | 47 | 0.37 | 0.327 |
| Keri et al, 2014 [50] | 50 | 1.2 | 0.7 | 30 | 0.8 | 0.5 |
| Kim et al, 2021 [53] | 27 | 1.064 | 0.5719 | 26 | 0.7237 | 0.3694 |
| Lan et al, 2022 [57] (females) | 72 | 0.34 | 0.56 | 25 | -0.26 | 0.68 |
| Lan et al, 2022 [57] (males) | 55 | 0.08 | 0.59 | 35 | -0.34 | 0.72 |
| Leo et al, 2006 [60] | 46 | 2.59 | 1.45 | 46 | 1.26 | 0.63 |
| Liu et al, 2022 [74] | 66 | 118.04 | 26.37 | 43 | 122.74 | 23.4 |
| Sugimoto et al, 2018 [103] | 35 | 0.616 | 0.812 | 35 | 0.41 | 0.364 |
| Tang et al, 2021 [106] | 139 | 3.38 | 2.12 | 76 | 3.52 | 1.17 |
| Zhao et al, 2022 [132] | 24 | 901.03 | 617.92 | 26 | 707.31 | 584.07 |
| Zou et al, 2018 [138] | 75 | 1.61 | 1.6 | 102 | 1.5 | 1.29 |

(d) IL-8

| Study | Patients (n) | Patients Mean | Patients SD | Controls (n) | Controls Mean | Controls SD |
|---|---|---|---|---|---|---|
| Gao et al, 2024 [31] | 23 | 451.24 | 383.24 | 23 | 26 | 21.29 |
| Lan et al, 2022 [57] (females) | 72 | 0.61 | 0.37 | 25 | 0.46 | 0.37 |
| Lan et al, 2022 [57] (males) | 55 | 0.51 | 0.3 | 35 | 0.55 | 0.4 |
| Zou et al, 2018 [138] | 75 | 8.94 | 3.99 | 102 | 18.67 | 12.12 |

(e) TNF-α

| Study | Patients (n) | Patients Mean | Patients SD | Controls (n) | Controls Mean | Controls SD |
|---|---|---|---|---|---|---|
| Chen et al, 2022a [15] | 29 | 48.45 | 29.29 | 25 | 41.35 | 15.86 |
| Gao et al, 2024 [31] | 23 | 4.5 | 2.43 | 23 | 2.88 | 0.87 |
| Kakeda et al, 2018 [47] | 40 | 1.592 | 0.576 | 47 | 2.041 | 5.39 |
| Kim et al, 2021 [53] | 50 | 9.32 | 5.3 | 50 | 9.87 | 6.38 |
| Lan et al, 2022 [57] (females) | 72 | 1.21 | 0.33 | 25 | 0.71 | 0.21 |
| Lan et al, 2022 [57] (males) | 55 | 0.83 | 0.21 | 35 | 0.74 | 0.22 |
| Leo et al, 2006 [60] | 46 | 3.37 | 1.87 | 46 | 1.44 | 0.83 |
| Li et al, 2013 [61] | 64 | 2.2 | 0.35 | 64 | 1.98 | 0.34 |
| Liu et al, 2022 [74] | 66 | 471.54 | 123.17 | 43 | 473.89 | 106.05 |
| Sugimoto et al, 2018 [103] | 35 | 1.612 | 0.598 | 35 | 2.336 | 6.239 |
| Tang et al, 2021 [106] | 139 | 2.35 | 1.67 | 76 | 1.69 | 0.68 |
| Zhao et al, 2022 [132] | 24 | 514.12 | 194.41 | 26 | 414.87 | 288.36 |
| Zou et al, 2018 [138] | 75 | 3.79 | 0.71 | 102 | 2.69 | 1.46 |

**Supplementary Table 16. Meta-analysis raw data of anti-inflammatory biomarkers in major depressive disorder.**

IL-10

| Study | Patients (n) | Patients Mean | Patients SD | Controls (n) | Controls Mean | Controls SD |
|---|---|---|---|---|---|---|
| Lan et al, 2022 [57] (females) | 72 | 1.5 | 0.44 | 25 | 1.1 | 0.37 |
| Lan et al, 2022 [57] (males) | 55 | 1.47 | 0.39 | 35 | 1.08 | 0.49 |
| Liu et al, 2022 [74] | 66 | 146.02 | 35.79 | 43 | 148.67 | 34.17 |
| Tang et al, 2021 [106] | 139 | 2.04 | 1.3 | 76 | 1.05 | 0.92 |
| Zou et al, 2018 [138] | 75 | 0.55 | 0.38 | 102 | 0.26 | 0.23 |

**Supplementary Table 17. Meta-analysis raw data of other inflammatory biomarkers in major depressive disorder.**

C-Reactive Protein (CRP)

| Study | Patients (n) | Patients Mean | Patients SD | Controls (n) | Controls Mean | Controls SD |
|---|---|---|---|---|---|---|
| Chang et al, 2017 [10] | 72 | 323.1 | 431.72 | 96 | 177.4 | 215.02 |
| Chen et al, 2022a [15] | 29 | 0.88 | 1.07 | 25 | 1.11 | 1.07 |
| Keri et al, 2014 [50] | 50 | 1.6 | 1.4 | 30 | 0.9 | 1.1 |
| Kim et al, 2021 [53] (females) | 27 | 790.8 | 632.4 | 26 | 415.2 | 361.5 |
| Kuwano et al, 2018 [55] | 15 | 5.18 | 1.01 | 19 | 5.66 | 1.36 |
| Liu et al, 2022 [74] | 66 | 116.8 | 27.35 | 43 | 103.14 | 24.92 |

**Supplementary Table 18. Meta-analysis raw data of BMI in major depressive disorder.**

Body mass index (BMI)

| Study | Patients (n) | Patients Mean | Patients SD | Controls (n) | Controls Mean | Controls SD |
|---|---|---|---|---|---|---|
| Bai et al, 2021 [4] | 53 | 23.41 | 2.37 | 86 | 23.09 | 2.14 |
| Chang et al, 2017 [10] | 72 | 22.25 | 4.11 | 96 | 22.78 | 3.68 |
| de Menezes Galvao et al, 2021 [22] | 30 | 25.65 | 5.42 | 32 | 23.98 | 4.78 |
| Gao et al, 2024 [31] | 23 | 22.63 | 3.83 | 23 | 23.95 | 3.06 |
| Herran et al, 2000 [40] | 19 | 25 | 4.9 | 19 | 26.4 | 6.1 |
| Keri et al, 2014 [50] | 50 | 23.2 | 12.7 | 30 | 24 | 13.7 |
| Kim et al, 2021 [53] | 50 | 22.68 | 4.04 | 50 | 21.83 | 4.77 |
| Lan et al, 2022 [57] (females) | 72 | 20.2 | 3.37 | 25 | 22.06 | 3.08 |
| Lan et al, 2022 [57] (males) | 55 | 21.9 | 2.99 | 35 | 22.73 | 4.27 |
| Leo et al, 2006 [60] | 46 | 27.7 | 1.99 | 46 | 28.29 | 2.06 |
| Li et al, 2013 [61] | 64 | 21.3 | 2.2 | 64 | 21.5 | 2.9 |
| Liang et al, 2015 [69] | 156 | 22.6 | 3.4 | 49 | 22.8 | 2.3 |
| Liu et al, 2015 [73] | 35 | 22.09 | 12.54 | 35 | 22.21 | 10.18 |
| Liu et al, 2022 [74] | 66 | 21.46 | 4.61 | 43 | 21.81 | 2.15 |
| Puangsri and Ninla-Aesong, 2021 [85] | 137 | 23.01 | 0.52 | 56 | 22.75 | 0.52 |
| Targitay Ozturk et al, 2023 [108] | 58 | 27.24 | 6.05 | 58 | 26.2 | 5.54 |
| Wang et al, 2025 [67] | 237 | 23.08 | 4.14 | 89 | 24.41 | 6.29 |
| Xiong et al, 2011 [115] | 30 | 22.63 | 1.29 | 28 | 22.45 | 1.23 |
| Yang et al, 2021 [122] | 34 | 21.6 | 3 | 34 | 23.9 | 4.6 |
| Zhang et al, 2016b [131] | 50 | 22.7 | 1.4 | 50 | 23.3 | 1 |
| Zou et al, 2018 [138] | 75 | 21.89 | 2.89 | 102 | 23.37 | 3.12 |

**Supplementary Table 19. Meta-analysis raw data of BMI in bipolar disorder.**

Body mass index (BMI)

| Study | Patients (n) | Patients Mean | Patients SD | Controls (n) | Controls Mean | Controls SD |
|---|---|---|---|---|---|---|
| Chang et al, 2017 [10] | 88 | 22.83 | 4.42 | 96 | 22.78 | 3.68 |
| Chen et al, 2022b [16] | 191 | 23.06 | 4.32 | 112 | 23.12 | 3.94 |
| Li et al, 2024a [63] | 130 | 21.92 | 3.46 | 108 | 22.01 | 3.69 |
| Liu et al, 2023 [75] | 66 | 21.37 | 2.65 | 66 | 21.29 | 2.37 |

**Supplementary Table 20. Meta-analysis after excluding studies with unknown number of episodes.**

| Marker | Category | No. of Studies | Patients (n) | Controls (n) | Statistic (SMD) | P-value | Effect Size | Heterogeneity ($I^2$) |
|---|---|---|---|---|---|---|---|---|
| | | | | **SCZ** | | | | |
| IFN-γ* | Immune | 7 | 317 | 266 | 0.81 (95% CI: 0.28 to 1.35) | 0.003 | Large | High (89%) |
| TNF-α | Immune | 8 | 410 | 426 | 0.03 (95% CI: -0.5 to 0.55) | 0.992 | Very small | High (92.31%) |
| IL-6* | Immune | 10 | 490 | 453 | 0.88 (95% CI: 0.29 to 1.47) | 0.004 | Large | High (94.36%) |
| IL-17 | Immune | 3 | 270 | 154 | -0.73 (95% CI: -2.80 to 1.35) | 0.493 | Medium | High (98.85%) |
| IL-4* | Immune | 6 | 248 | 206 | 0.71 (95% CI: 0.27 to 1.15) | 0.002 | Medium | High (78.87%) |
| IL-10 | Immune | 4 | 142 | 145 | 0.62 (95% CI: -0.38 to 1.61) | 0.223 | Medium | High (93.69%) |
| IL-2 | Immune | 4 | 142 | 145 | 0.49 (95% CI: -0.03 to 1.01) | 0.066 | Small | High (78.23%) |
| BMI | Metabolic function | 36 | 3490 | 3289 | -0.04 (95% CI: -0.21 to 0.13) | 0.67 | Very small | High (90.43%) |
| Fasting glucose | Metabolic function | 30 | 2862 | 2189 | 0.17 (95% CI: -0.10 to 0.43) | 0.214 | Very small | High (94.69%) |
| Insulin* | Metabolic function | 17 | 1652 | 1268 | 0.66 (95% CI: 0.44 to 0.87) | < 0.001 | Medium | High (85.08%) |
| HOMA-IR* | Metabolic function | 14 | 1541 | 1086 | 0.66 (95% CI: 0.44 to 0.87) | < 0.001 | Medium | High (81.56%) |
| Cholesterol | Metabolic function | 24 | 2566 | 1994 | 0.08 (95% CI: -0.21 to 0.36) | 0.60 | Very small | High (94.88%) |
| Triglycerides* | Metabolic function | 28 | 2960 | 2216 | 0.37 (95% CI: 0.13 to 0.60) | 0.003 | Small | High (93.63%) |
| | | | | **MDD** | | | | |
| IFN-γ* | Immune | 3 | 202 | 142 | 0.50 (95% CI: 0.06 to 0.94) | 0.027 | Medium | High (74.13%) |
| TNF-α* | Immune | 7 | 407 | 320 | 0.50 (95% CI: 0.04to 0.95) | 0.033 | Medium | High (85.51%) |
| IL-1β | Immune | 7 | 348 | 265 | 0.12 (95% CI: -0.35 to 0.58) | 0.629 | Medium | High (87.24%) |
| IL-6* | Immune | 8 | 393 | 286 | 0.46 (95% CI: 0.11 to 0.82) | 0.011 | Small | High (79.94%) |
| IL-10 | Immune | 3 | 193 | 103 | 0.58 (95% CI: -0.08 to 1.23) | 0.087 | Medium | High (85.6%) |
| CRP | Immune | 5 | 232 | 213 | 0.25 (95% CI: -0.10 to 0.59) | 0.161 | Small | Moderate (65.14%) |
| BMI | Metabolic function | 15 | 919 | 678 | -0.07 (95% CI: -0.21 to 0.08) | 0.386 | Very small | Moderate (49.3%) |

*p<0.05 FDR corrected.
